# Is exposure to chemical pollutants associated with sleep outcomes? A systematic review

**DOI:** 10.1101/2022.11.02.22281802

**Authors:** Danielle A. Wallace, Jayden Pace Gallagher, Shenita R. Peterson, Seyni Ndiaye-Gueye, Kathleen Fox, Susan Redline, Dayna A. Johnson

## Abstract

**Background:** Sleep disruption is a public health concern and environmental exposures that influence the biological processes underpinning sleep may contribute to impaired sleep health. However, the contributions of environmental chemical pollutants to sleep health have not been systematically investigated.

**Objectives:** This systematic literature review sought to identify, evaluate, summarize, and synthesize the existing evidence between environmental chemical pollutant exposure and dimensions of sleep health in humans. We sought to evaluate potential mechanisms underlying exposure-outcome relationships and recommend areas for future research.

**Methods:** Databases were searched for peer-reviewed published research on chemical environmental pollutants and sleep health and disorders with no date restriction up to the last search date of April 2021 (registered protocol PROSPERO # CRD42021256918), resulting in 9,918 records. Two reviewers independently screened identified records against inclusion and exclusion criteria, extracted study information, and performed risk of bias assessments.

**Results:** We identified 204 studies of exposure to air pollution, exposures related to the Gulf War and other conflicts, endocrine disruptors, metals, pesticides, and solvents with dimensions of sleep health (architecture, duration, quality, timing) and sleep disorders (sleeping pill use, insomnia, sleep-disordered breathing) for inclusion in this review. Metals were the most frequently evaluated pollutants, while sleep maintenance/insomnia and quality were the most reported sleep outcomes. Synthesis of the evidence suggests mechanisms related to cholinergic signaling, neurotransmission, and inflammation as the most shared pathways.

**Discussion:** Evidence indicates that exposure to particulate matter, secondhand smoke, dioxins and dioxin-like compounds, lead, mercury, pesticides, solvents, and exposures related to the Gulf War are associated with worse sleep health and disorders. Chemical pollutants are likely key environmental determinants of sleep health and disorders; thus, there is a need to improve measurement of pollutants and sleep, study rigor, and determine the populations most vulnerable to the effects of exposure.

## INTRODUCTION

Sleep is as vital to life as air or water and is essential for metabolic, immunologic, developmental, and cognitive functions[1]. Healthy sleep is a multidimensional construct defined by sleep duration, efficiency, timing, alertness, and quality[2]. However, disrupted sleep is prevalent and of public health concern[3–5]; for example, two thirds of high school students[6] and a third of adults[7] in the United States experience disrupted sleep. In the U.S., approximately 31-40% of people younger than 18 years of age do not attain the recommended sleep duration for their age group[8], and 11% of adults report sleeping 5 hours or less per night[8]. Obstructive sleep apnea (OSA), a form of sleep-disordered breathing (SDB) characterized by breathing cessations that disrupts sleep, is estimated to affect a billion people worldwide[9]. Chronic insomnia – difficulty falling asleep and maintaining sleep – is another common sleep disorder affecting approximately 6% of the population[10]. The pervasiveness of these outcomes is alarming because disrupted sleep acts as an endocrine disruptor, preventing or altering the secretion of growth hormone, prolactin, cortisol, and other hormones[11–14], and is associated with inflammation[15–18] and cardiovascular[19–21], and cardiometabolic diseases [22–29].

Sleep is a dynamic neurophysiological process characterized by cycles of non-rapid eye movement (NREM) and rapid eye movement (REM) states. Sleep is regulated by a 2-part process composed of a 1) homeostatic sleep drive (Process S) and 2) and a circadian rhythm in wakefulness (Process C)[30]. These processes are governed by neurological circuits, neurotransmitters, and hormones that drive or inhibit arousal and sleep, with interaction from the circadian system to influence the timing, latency, and quality of sleep.

Pathways that promote arousal include monoaminergic, cholinergic, glutamatergic, histaminergic, and GABAergic neurotransmission from the midbrain, basal forebrain, and other regions[31–33]; during sleep promotion, these pathways are inhibited and somnogens such as adenosine, cytokines, and melatonin aid to induce sleep. These processes and sleep-wake behavior can be altered by untreated OSA, insomnia, individual behaviors and health status, socioeconomic status[34], occupation[35, 36], interpersonal relationships, social and cultural factors, societal factors[37–39], and environmental factors[38]. To improve sleep at a population-level, further understanding of environmental factors is warranted.

Environmental exposures are particularly important for sleep health, and chemical pollutants are common environmental exposures with potential for chronic effects on the respiratory system, cardiovascular system, central nervous system, and neuroendocrine system. Prior systematic reviews of environmental exposures and sleep have reported summarized associations with these physical components, such as reviews on sleep and noise pollution[40, 41], light pollution[42], greenspace[43], climate change[44], and other aspects of the built environment[45]. Other environmental exposures, such as air pollution[45–49], second-hand smoke exposure[50, 51], micronutrients[52], and occupational exposures[35], have also been reviewed as contributors to sleep health.

While understanding of the social and environmental factors that affect sleep is growing, less is known regarding the effects of chemical pollutants such as pesticides, heavy metals, solvents, and endocrine-disrupting chemicals (EDCs) on sleep health. Chemical pollutants have been shown to affect neurotransmitter systems and other sleep-relevant processes such as inflammation[53–55]. For example, animal and in vitro studies have shown that lead exposure alters GABA release[56], and a study in monkeys reported that treatment with lead during infancy caused insomnia[57]. Experimental and epidemiological evidence links exposure to air pollution and inflammation[58, 59], an established risk factor for sleep disturbances[17] and OSA[60], and pulmonary impairment due to air pollution may contribute to SDB[61]. Altered cholinergic signaling has also been implicated in upper airway collapsibility and OSA[62–64], and this may be a pathway by which environmental exposures that act on the cholinergic system, such as pesticides, influence SDB. For example, a rat study showed that exposure to chlorpyrifos, a commonly used pesticide, during early development resulted in a higher sleep apnea index and lower diaphragm acetylcholinesterase (AChE) activity later in life, compared to rats not developmentally exposed to chlorpyrifos[65]. Therefore, exposure to chemical pollutants may result in disturbed sleep by acting upon the biological pathways that regulate sleep-wake behavior, with developmental windows of vulnerability; however, the connections between environmental chemical exposures and sleep disturbance are not well-established.

Importantly, environmental pollution does not affect all people equally, with systematic differences in pollution exposure by race/ethnicity and class[66]; disturbed sleep and sleep disorders are also more common among historically marginalized racial groups and individuals of lower socioeconomic status. Due to environmental racism, marginalized communities face a disproportionately higher burden of adverse environmental exposures[67], with environmental disparities in housing[68] and ambient pollution[69], which may contribute to sleep health disparities[70–72]. Individuals living in these communities also have a greater burden of disease associated with these adverse exposures, such as higher rates of asthma[73] and OSA morbidity and mortality[74, 75]. Early exposure during childhood influences health trajectories later in life, and research in environmental determinants of sleep health can benefit from a life course approach with particular focus on environmental exposures during vulnerable developmental windows and critical periods[76]. Thus, identifying the environmental factors that lead to disrupted sleep in both childhood and adulthood can inform public health efforts to improve sleep health and address disparities in sleep and related morbidity [75,77–79].

In summary, chemical pollutants may contribute to sleep disruption by influencing the underlying biology of sleep-wake behavior, with possible age-specific effects and disparities in exposure. Prior studies have assessed links between chemical pollutants and sleep outcomes, but this literature has not yet been evaluated in a comprehensive, systematic way; doing so would provide a starting point from which to assess the interactions between chemical environmental exposures and sleep health. Therefore, to address these gaps in knowledge and to synthesize the current epidemiological evidence, we conducted a systematic review of the relationships between chemical pollutants and sleep health and disorders. Furthermore, we discuss potential mechanisms linking pollutants to sleep outcomes and areas for future research.

## METHODS

### Data Sources and Literature Search Strategy

The Preferred Reporting Items for Systematic Reviews and Meta-Analyses (PRISMA) statement was used as a guideline for conducting this review. We registered our protocol with the international prospective register of systematic reviews, PROSPERO (#CRD42021256918, first submitted May 27, 2021). As outlined by the PROSPERO protocol, a comprehensive search was created by the research team and executed by information specialists (SP and KF) in 6 bibliographic databases: Agricultural & Environmental Science Collection (ProQuest), PubMed, Embase (Elsevier), Environment Complete (Ebsco), Web of Science Core Collection [Science Citation Index Expanded, Social Sciences Citation Index, Arts & Humanities Citation Index, Emerging Sources Citation Index, Conference Proceedings Citation Index, Book Citation Index, Current Chemical Reactions, Index Chemicus], and Scopus (Elsevier) in the Spring of 2021. The search was crafted of various keywords and controlled vocabulary concerning the concept of ‘environmental pollutants’ and the sleep outcomes listed in the inclusion criteria. Searches were limited to articles and reviews written in English. We did not restrict by date. Animal studies were excluded from the PubMed and Embase searches. Search terms, search dates, and additional details are provided in **Supplemental File 1**. The database search retrieved 9,918 citations. Studies were imported into EndNote X9 for data management and deduplication. Additional duplicates were removed by the web-based evidence synthesis tool Covidence, which was used for screening and data extraction.

### Inclusion criteria

Inclusion criteria included: research in humans, chemical pollutant as an exposure (such as metals, pesticides, flame retardants, etc.), sleep health or disorder (such as sleep quality or SDB), sleep measurement (polysomnography (PSG), actigraphy, etc.), and quantitative data. Studies of in vitro or cultured tissue, completely modeled data, or non-human data were excluded from the analysis. Of the environmental exposure literature, studies of noise pollution, light pollution, radiation, smell, green space, nutrition, allergens, and/or mold were excluded from the analysis. A recent systematic review by Liu et al[46] summarized the air pollution and sleep literature (n=22 articles) up until the last search date of October 2019; therefore, we excluded those studies of air pollution and sleep already reviewed by Liu et al[46] and include discussion of our findings with the previously reviewed literature. Metals are common environmental pollutants[80], and while we excluded most studies of micronutrients and sleep, we included those of manganese, copper, and selenium, because they have relevance to pesticide use and as environmental contaminants. Studies of Parkinson’s disease, Alzheimer’s disease, and Wilson’s disease were identified and excluded. Symptoms of sleep disorders such as snoring, restless legs syndrome, and REM sleep behavior disorder were also identified and excluded due to being outside the scope of this review. Lastly, studies with only qualitative data, not available in English, not peer-reviewed, or epidemiologic studies without a control or comparator group, such as varying levels of exposure, were also excluded. Studies with multiple measures of sleep that met inclusion criteria but also evaluated snoring, for example, were extracted with information on valid sleep outcome(s) only. Inclusion categories were further reviewed during mutual group discussions (DW, JPG, DAJ).

### Screening

The resulting papers were screened by two independent reviewers (DW and JP) against inclusion and exclusion criteria using Covidence. The number of articles screened during the two steps can be found in the Preferred Reporting Items for Systematic Reviews and Meta-Analyses (PRISMA) flowchart in **Figure 1**. The search resulted in 4,464 unique citations identified for screening after de-duplication. After the full-text screening, bibliographies of 20 review articles (**Supplemental File 2**) were searched for additional citations in February 2022, resulting in an additional 58 articles (2 duplicates) added to Covidence for title/abstract and full-text screening. Of the initial 4,464 papers and 58 papers identified from reviews, 3,984 articles were considered irrelevant and excluded during the title/abstract screening process. Full text review of the remaining 538 papers resulted in 204 studies included for data extraction, 15 of which were identified from the review articles. Of the included articles, 74% (n=151) were epidemiological studies and 26% (n=53) were case reports or case series Consensus was arbitrated by the first author during title/abstract and full-text screening, and, in the instance of discordant responses, consensus was determined by mutual discussion (DW and JPG).

**Figure 1.**
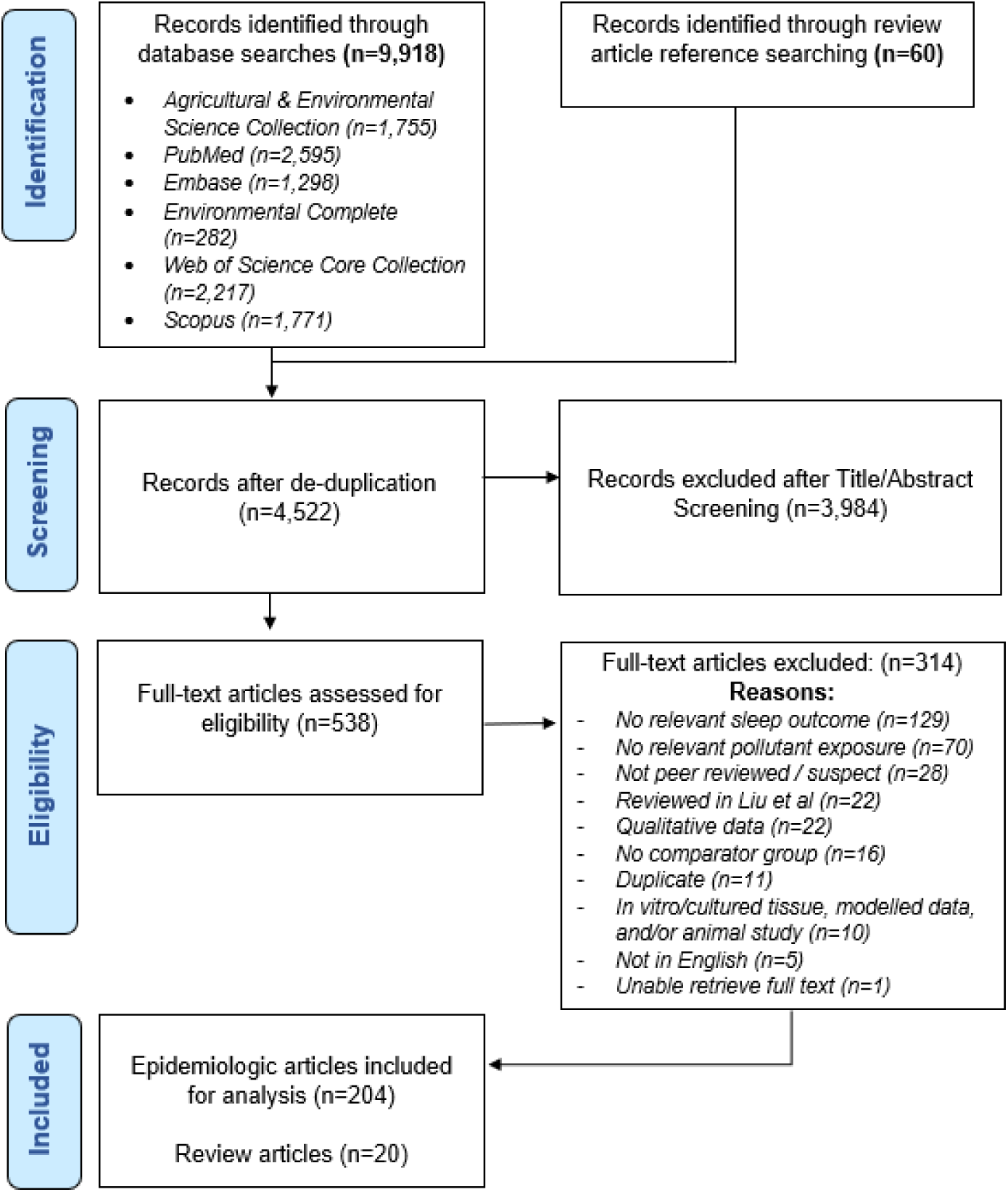
PRISMA flow chart detailing screening and inclusion of studies in this review.

### Data Extraction

Data on year, study location, study design, exposure and exposure measurement, outcome and outcome measurement, and risk of bias (RoB, details below) for the 204 included studies was independently extracted by two reviewers (DW and JPG) using Covidence and Google Forms (abstraction forms available in **Supplemental File 3**). PSG studies inclusive of sleep stages were considered measures of sleep architecture, which refers to sleep stage composition (stages N1, N2, N3 and REM) and the cycles of REM and NREM. Outcomes of sleeplessness, inability to sleep, sleep onset latency, problems with sleep maintenance (such as wake after sleep onset or sleep fragmentation), and early morning awakening were considered measures of sleep maintenance/insomnia. Studies where the sleep outcome was described as a sleep problem or disorder (not insomnia or SDB), sleep disturbance, or sleep efficiency were included in the sleep quality group. Outcomes related to sleep timing or sleep midpoint were included in the sleep timing group.

However, we recognize that extracted exposure or outcome categories are not exclusive, and that an exposure or an outcome listed in one group may also have characteristics that overlap with another group (for example, antimony is a heavy metal that is also used as a flame retardant). Studies where exposure and outcome of interest were measured concurrently were considered cross-sectional. Results were merged, and in the instance of discordant responses, consensus determined by mutual discussion. For studies on smoking and/or secondhand smoke (SHS) exposure, only results pertaining to ambient or secondhand smoke exposure, and not active smoking or in utero exposure, were included. For all studies, only relevant information on included exposures and sleep outcomes were extracted. Studies involving participants younger than 18 years of age were considered pediatric populations. Estimates were rounded to the nearest 2 decimals, and, in some instances, unadjusted estimates were calculated from the provided data where indicated. Due to heterogeneity, meta-analysis and comparison of effect estimates was not performed.

### Risk of Bias (RoB) Assessment

Included epidemiologic studies were rated for risk of bias (RoB) according to human-related research questions adapted from both the NIEHS and National Toxicology Program’s Office of Health Assessment and Translation tool (https://ntp.niehs.nih.gov/ntp/ohat/pubs/riskofbiastool_508.pdf) as well as the Cochrane ROBINS-I tool (template tool available in **Supplemental File 4**). RoB was independently assessed by two reviewers (DW and JPG) using Covidence and Google Forms; when responses differed between reviewers, consensus was reached after discussion. For each study, reviewers evaluated whether there was a “Definitely low”, “Probably low”, “Probably high”, “Definitely high”, or “No information” RoB for the following components: selection bias, confounding, exposure assessment, attrition/exclusion bias, outcome assessment, reporting bias, and other bias. The “other” bias group included bias due to improper statistical methods or factors not captured by other bias groups that could influence the validity of the study. The overall RoB for epidemiologic studies was assessed first by intervention domains and then weighted across the domains considered most important for a final RoB determination; these domains included: selection bias, confounding, exposure assessment, and outcome assessment. For case reports and case series, RoB was graded according to a tool proposed by Murad et al[81]; this tool rates the quality of case studies according to 4 areas: selection, ascertainment, causality, and reporting (template tool available in **Supplemental File 4**). Overall judgement of case study quality was ascertained as recommended[81], with ratings of “poor”, “fair”, or “good”, and greater weight given to areas perceived most important for the study. Results were visualized using R version 4.1. Data extraction and quality assessment responses for epidemiological studies are provided in **Supplemental File 5** and for case studies in **Supplemental File 6**.

## RESULTS

The included epidemiologic studies (n=151) evaluated exposure to air pollution (19%), conflict-related exposures (10.5%), EDCs/other (8%), metals (34%), pesticides (8%), solvents (20.5%), and exposures with sleep outcomes related to sleep architecture (2.6%), dreams/nightmares (6%), duration (26.5%), quality (47.7%), timing (2.6%), sleeping pill use (4%), sleep maintenance/insomnia (37.1%), and SDB (23.8%), as shown in **Table 1**. The majority of case studies (n=53) reported exposure to metals (64%) and sleep outcomes such as insomnia (50.8%) and quality (29.2%) (**Table 2**); there were no case studies of air pollution, conflict-related exposures, or sleep timing. The majority of included studies were conducted in adults, with only 17.6% of epidemiologic and case studies including pediatric populations. Of the 22 epidemiologic and 14 case study articles with pediatric populations included in this review, the most common exposures were to metals (47%) and/or air pollution (28%), particularly secondhand smoke. Most epidemiologic studies relied on device-based measures of the exposure (61%) and self-reported measures of sleep (77%)(**Table 3**) using questionnaires such as the Pittsburgh Sleep Quality Index (PSQI, N=16)[82]. Among device-based sleep measures, PSG was most common (N=17), and only 5 studies assessed sleep via actigraphy with commercially-available wrist-worn devices; 2 additional studies relied upon static charge sensitive beds.

**Table 1.**
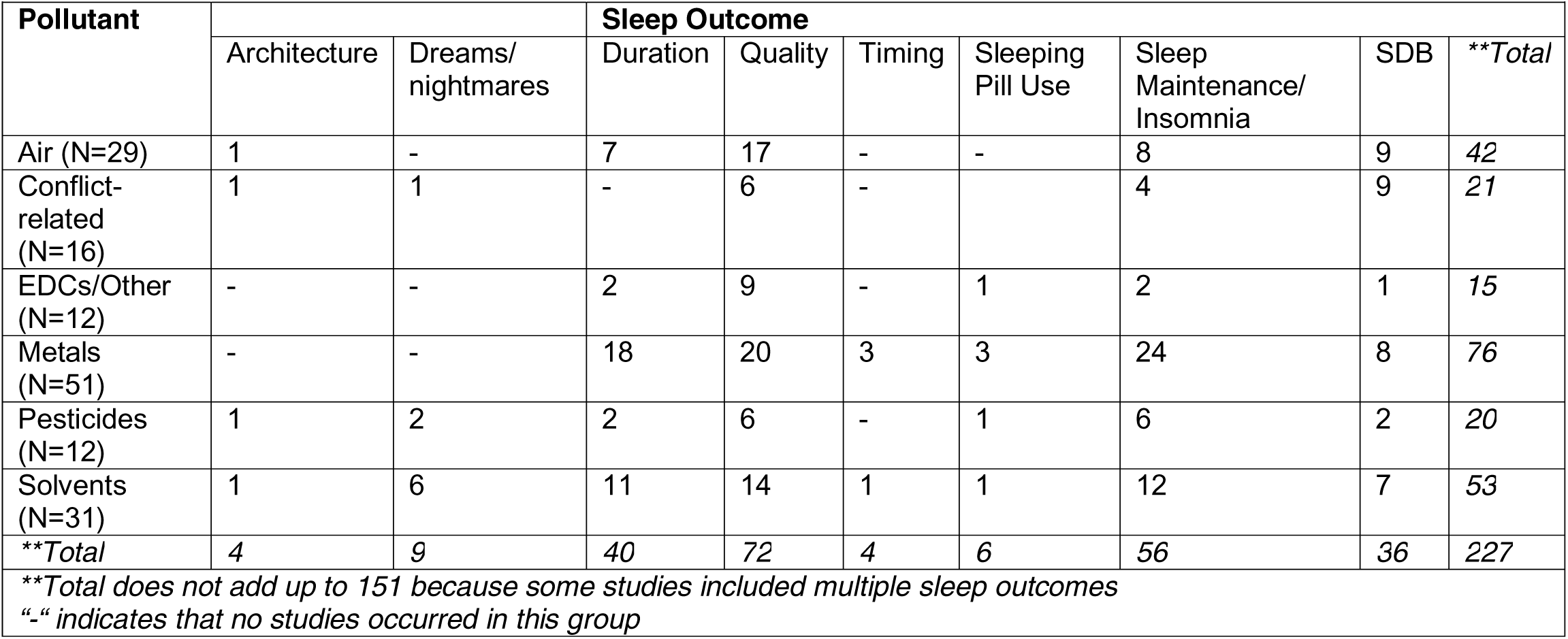
Breakdown of included epidemiologic studies (n=151) by pollutant exposure category and sleep outcome.

| Pollutant | Sleep Outcome |  |  |  |  |  |  |  |  |
| --- | --- | --- | --- | --- | --- | --- | --- | --- | --- |
|  | Architecture | Dreams/<br>nightmares | Duration | Quality | Timing | Sleeping<br>Pill Use | Sleep<br>Maintenance/<br>Insomnia | SDB | **\*\*Total** |
| Air (N=29) | 1 | - | 7 | 17 | - | - | 8 | 9 | 42 |
| Conflict-related<br>(N=16) | 1 | 1 | - | 6 | - |  | 4 | 9 | 21 |
| EDCs/Other<br>(N=12) | - | - | 2 | 9 | - | 1 | 2 | 1 | 15 |
| Metals<br>(N=51) | - | - | 18 | 20 | 3 | 3 | 24 | 8 | 76 |
| Pesticides<br>(N=12) | 1 | 2 | 2 | 6 | - | 1 | 6 | 2 | 20 |
| Solvents<br>(N=31) | 1 | 6 | 11 | 14 | 1 | 1 | 12 | 7 | 53 |
| **\*\*Total** | 4 | 9 | 40 | 72 | 4 | 6 | 56 | 36 | 227 |
| **\*\*Total does not add up to 151 because some studies included multiple sleep outcomes**<br>**“-“ indicates that no studies occurred in this group** |  |  |  |  |  |  |  |  |  |

**Table 2.**
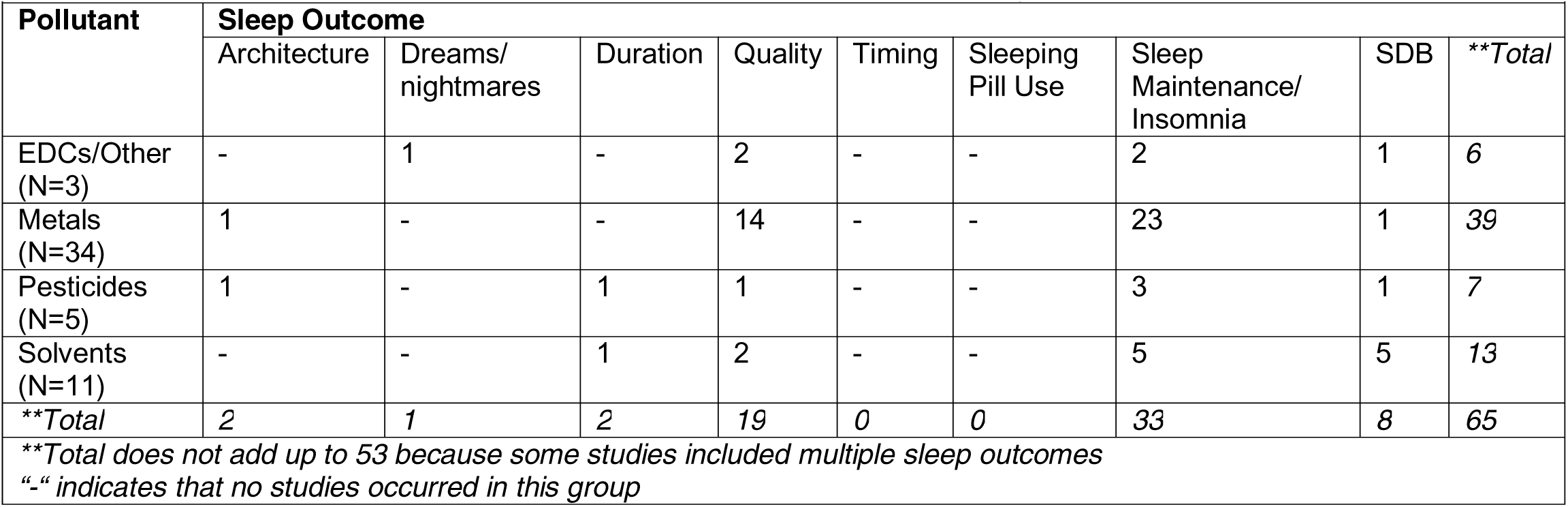
Breakdown of included case studies (n=53) by pollutant exposure category and sleep outcome

| Pollutant | Sleep Outcome |  |  |  |  |  |  |  |  |
| --- | --- | --- | --- | --- | --- | --- | --- | --- | --- |
|  | Architecture | Dreams/<br>nightmares | Duration | Quality | Timing | Sleeping<br>Pill Use | Sleep<br>Maintenance/<br>Insomnia | SDB | **Total |
| EDCs/Other<br>(N=3) | - | 1 | - | 2 | - | - | 2 | 1 | 6 |
| Metals<br>(N=34) | 1 | - | - | 14 | - | - | 23 | 1 | 39 |
| Pesticides<br>(N=5) | 1 | - | 1 | 1 | - | - | 3 | 1 | 7 |
| Solvents<br>(N=11) | - | - | 1 | 2 | - | - | 5 | 5 | 13 |
| **Total | 2 | 1 | 2 | 19 | 0 | 0 | 33 | 8 | 65 |
| **Total does not add up to 53 because some studies included multiple sleep outcomes<br>“-“ indicates that no studies occurred in this group |  |  |  |  |  |  |  |  |  |

**Table 3.** Frequency table of self-reported and device-measured exposure and outcome of included epidemiologic studies*

| Pollutant Exposure | Sleep Outcome |  |  |
| --- | --- | --- | --- |
|  | Device-measured | Self-reported | Total |
| Device-measured | 18 | 74 | 92 |
| Self-reported | 17 | 42 | 59 |
| Total | 35 | 116 | 151 |
| *if exposure or outcome measured with both device and self-report, grouped under device-measured. |  |  |  |

### Air Pollution

Pollutants in the air pollution category comprised: particulate matter (PM_1_, PM_2.5_, PM_10_), nitrogen oxides (NO_x_), ozone (O_3_), sulfur oxides (SO_x_), secondhand smoke (SHS) or environmental tobacco smoke (ETS), and volcanic ash (**Table 4**). Of the 29 included epidemiologic studies of air pollution, most occurred in urban environments in Asia and Europe, relied on device-based measures of exposure (65.5%), and evaluated self-reported measures (79%) of sleep quality or insomnia. Most air pollution studies of adult populations reported an association between ambient air pollution and disrupted sleep. Findings from the Henan Rural Cohort Study supported associations between PM and nitrogen dioxide (NO_2_) with longer sleep latency[83] and worse sleep quality[84, 85]; PM and NO_2_ levels were also linked to incident sleep disorders in a prospective cohort study in China[86]. However, smaller studies in Europe and the U.S. did not replicate these findings[87, 88]. The few pediatric studies of ambient air pollution investigated SDB, with heterogenous findings[89–91]. A study of UK Biobank data reported an association between NO_x_ and 7% increased odds of sleep disorders (which included insomnia and sleep apnea) when modelled as a singular pollutant, but a protective effect between NO_2_ and sleep disorders when PM_2.5_, PM_10_, and NO_x_ were included as covariates in the model[92]. An ecological study in Europe reported an association between NO2 exposure and SDB in children[90]. AQI, a measure of air quality and pollution in relation to national standards, was also found to be associated with insomnia in an ecological study[93] and admission for sleep disorders[94]. There were no associations between SO_2_ or CO and sleep, and no studies of ambient air pollution and sleep architecture or timing in adult or pediatric populations. The overall quality of the included studies was mixed (**Figure 2**).

**Figure 2.**
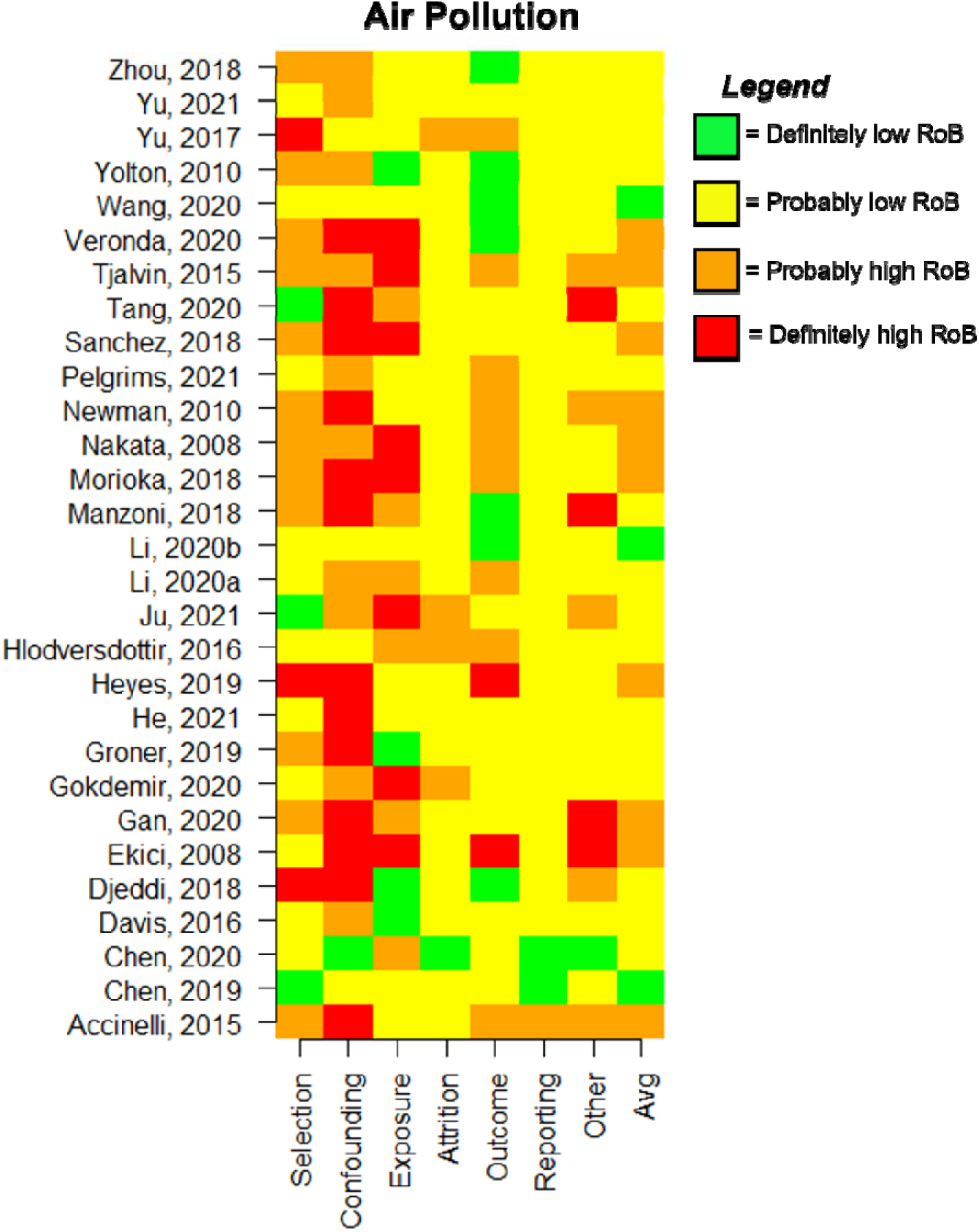
Chart depicting RoB ratings across evaluation domains for included epidemiologic studies on air pollution exposure.

**Table 4.**
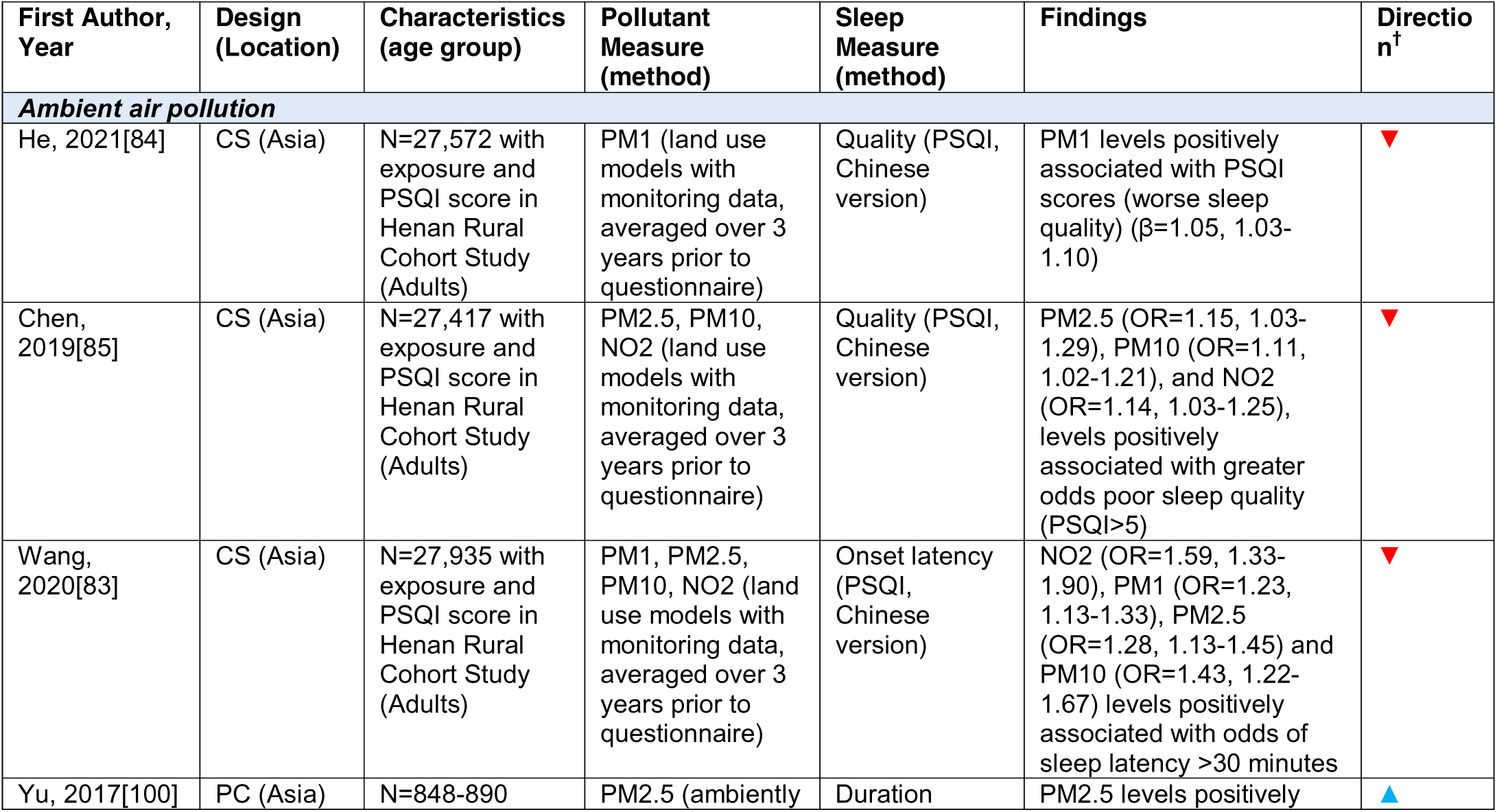
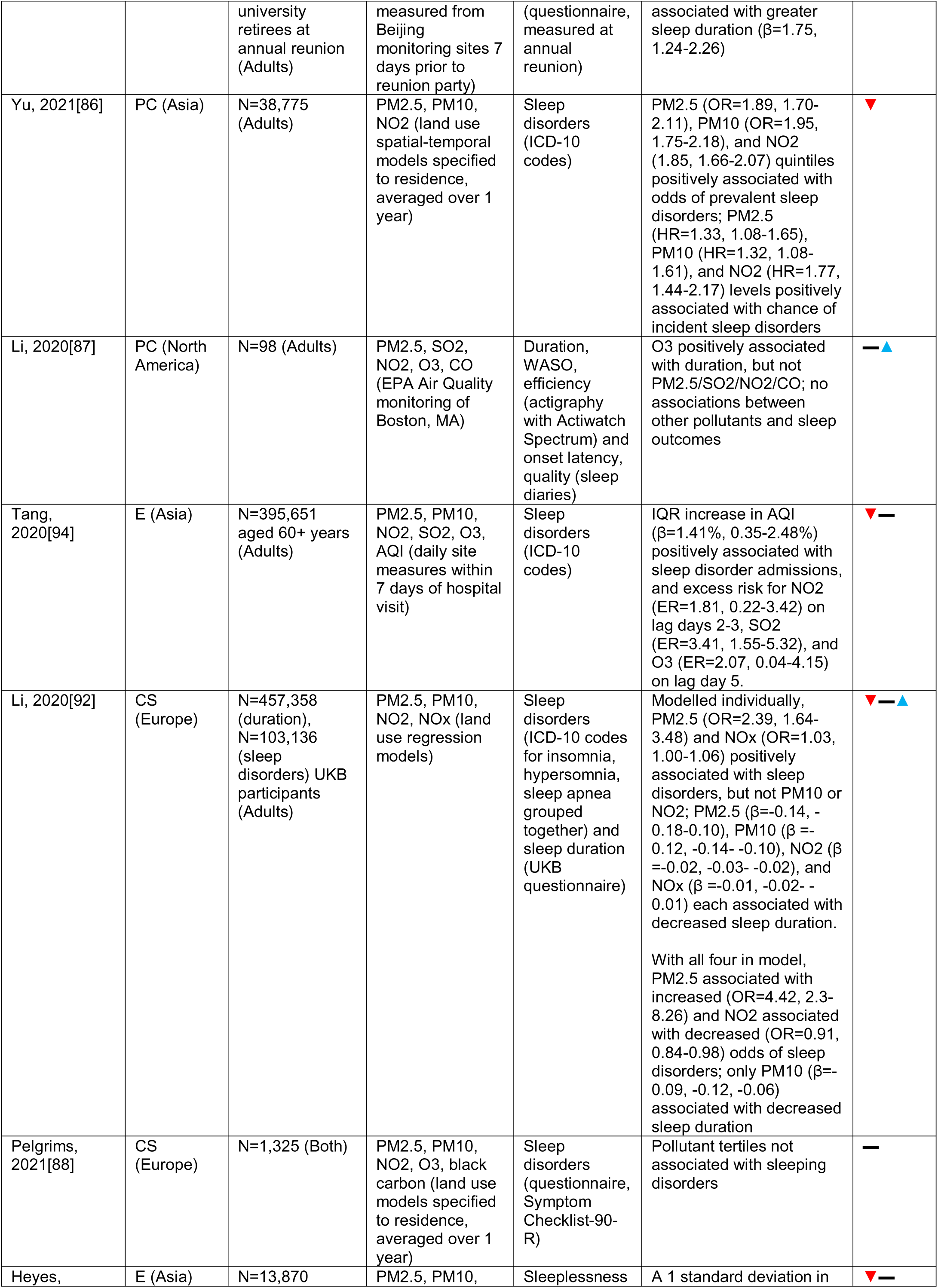
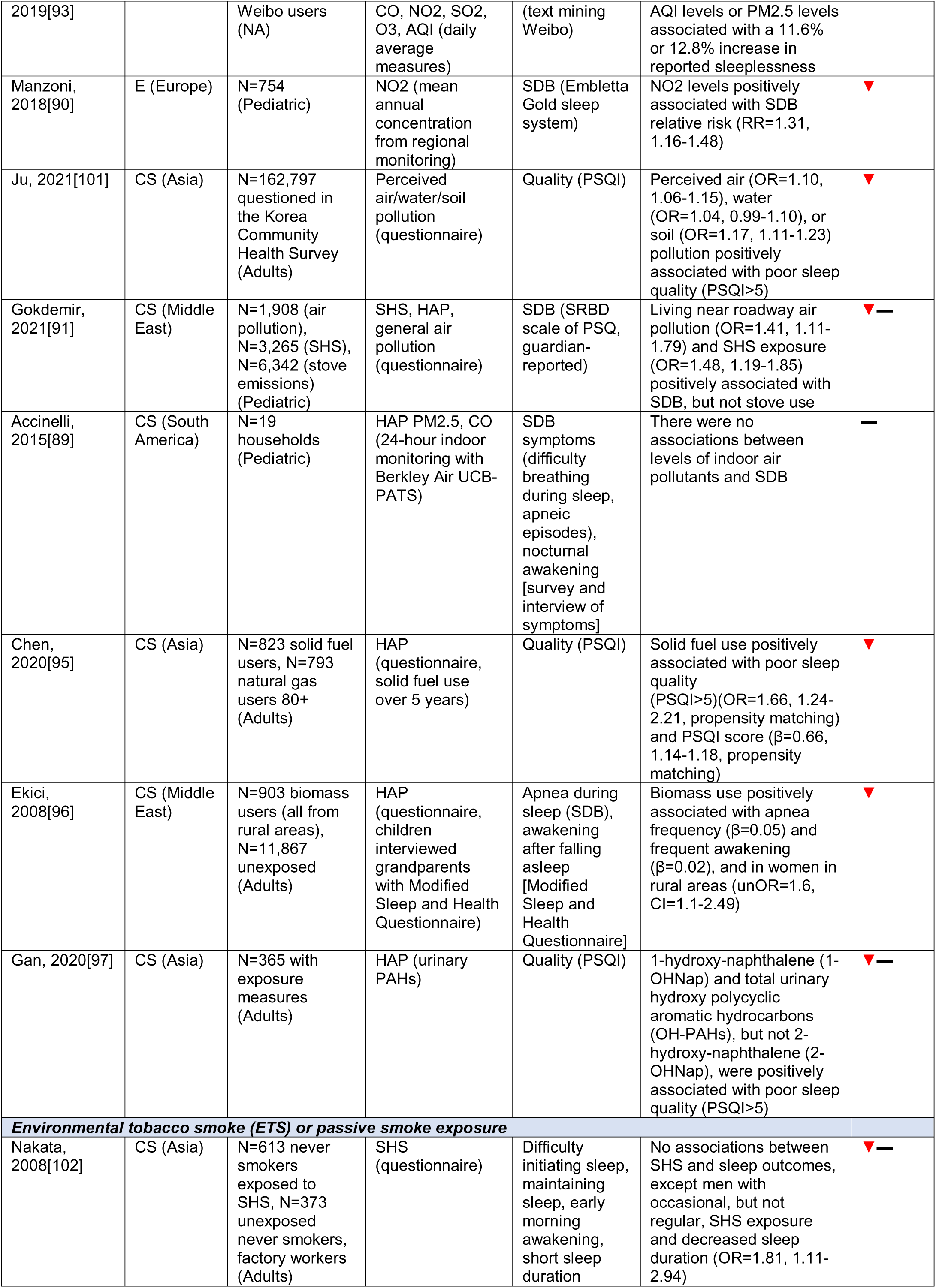
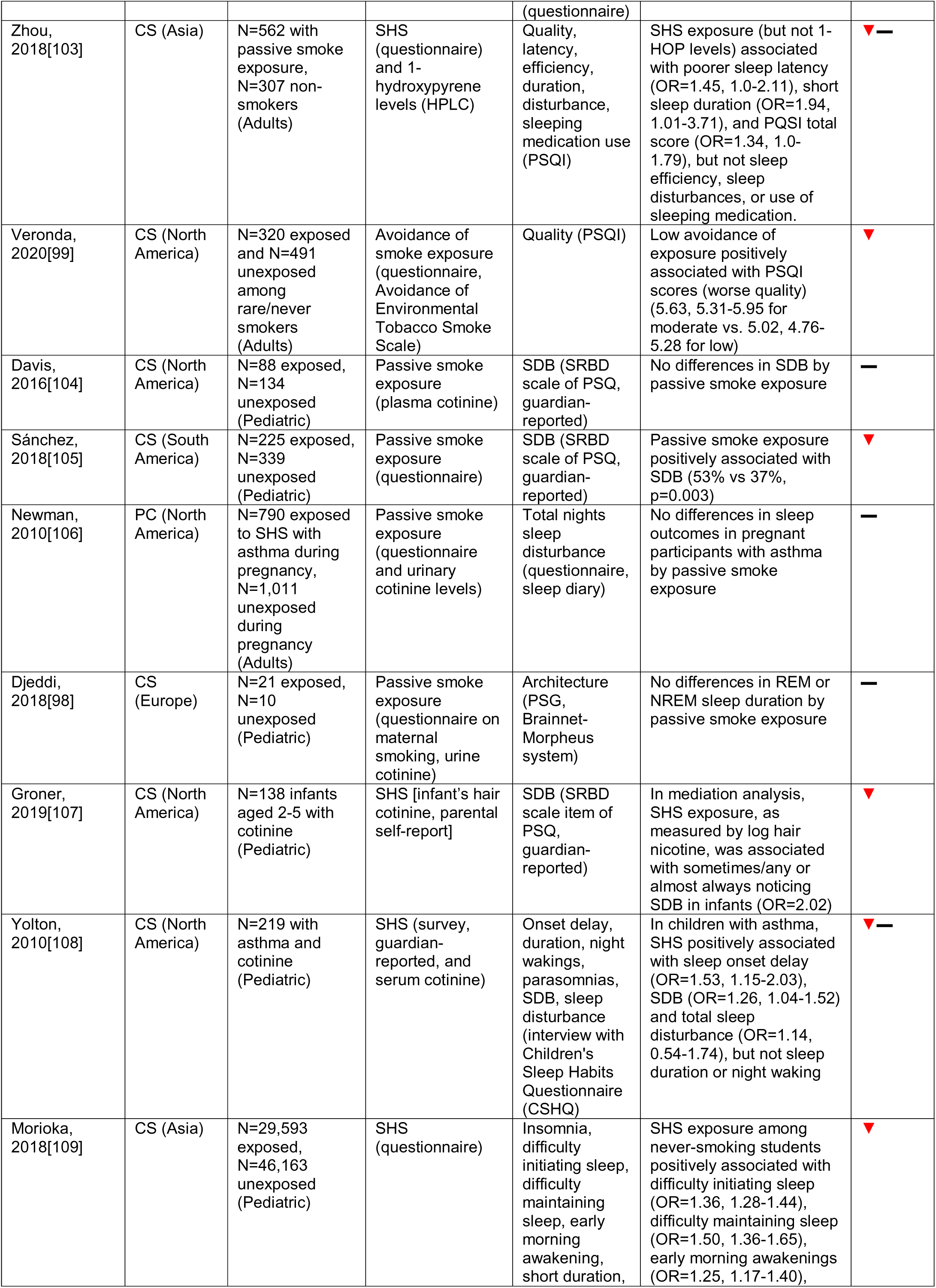
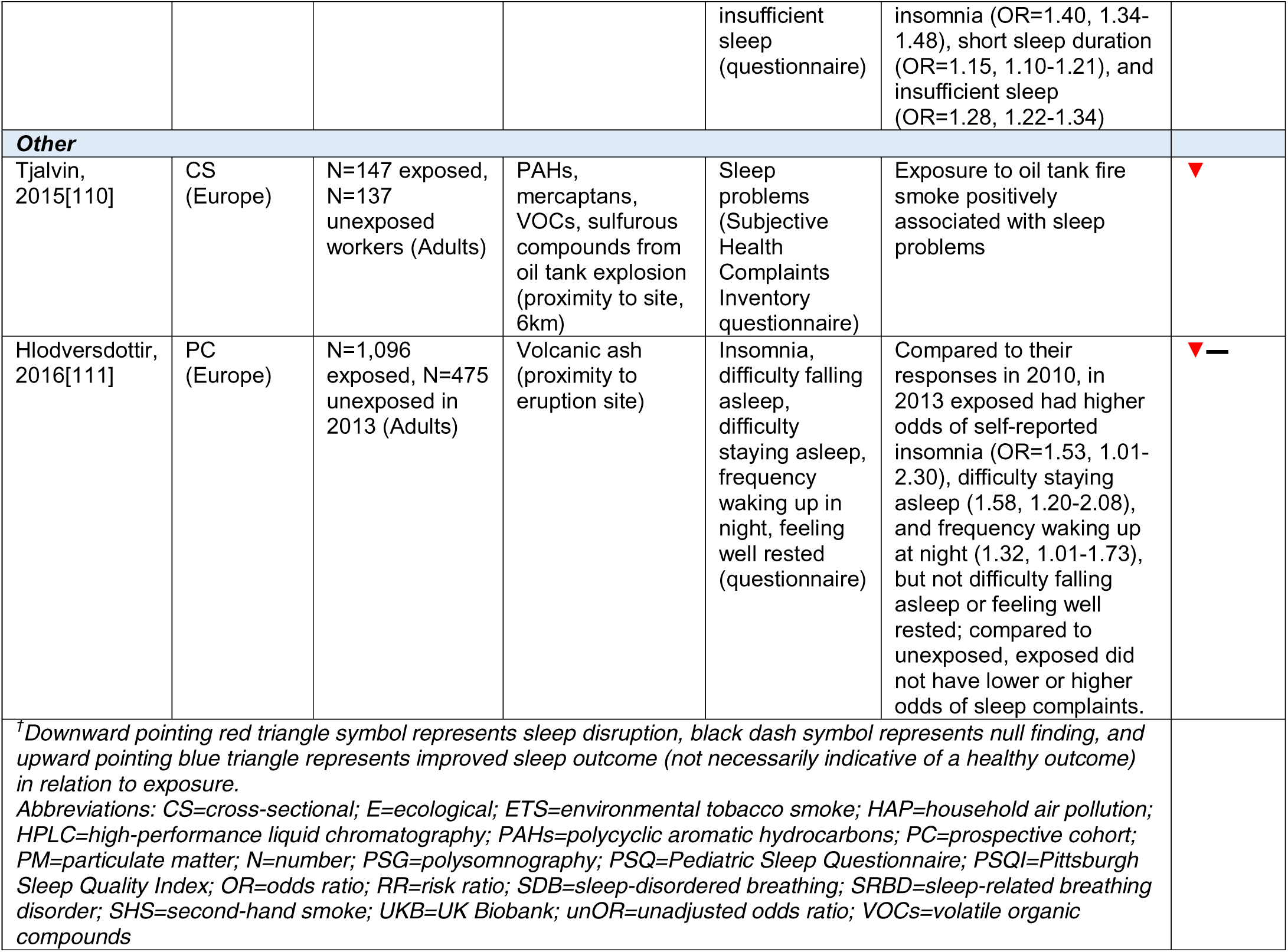
Summary table of epidemiologic studies of air pollutants/SHS and sleep outcomes.

Of the 5 studies which investigated indoor air pollution or household air pollution (HAP), the only study with device-based indoor air measurement reported null findings with SDB in children[89]; 2 studies reported associations between questionnaire-assessed solid fuel use and poor sleep quality[95] and biomass use and SDB and sleep disruption[96]; however, a larger study reported null findings between stove use and pediatric SDB[91]. One study with measures of urinary polycyclic aromatic hydrocarbon metabolites found an association between PAH exposure and poor sleep quality[97].

The majority of the 11 reviewed studies with ETS or SHS exposure reported positive associations between exposure and sleep disruption. Of these studies, only one included device-based measured sleep (PSG)[98] and found no association, and most assessed tobacco exposure through self-report. Of the 5 studies with measures of cotinine (a metabolite of nicotine), 2 reported associations with sleep impairment, while other results were null. ETS/SHS exposure measurement was heterogeneous; for example, some studies did not ask specifically about tobacco smoke exposure, but rather measured proximity to smokers or avoidance behavior[99]. Most pediatric studies demonstrated an association between ETS or SHS and self-reported SDB, insomnia, and poorer quality sleep.

### Conflict-related Exposures

There were 16 included epidemiologic studies with exposures related to the Gulf War and Gulf War Illness (GWI), the World Trade Center (WTC) toxic dust exposures of first responders, and other conflict-related exposures (**Table 5**). All investigations of GWI relied on self-reported measures, whereas WTC studies relied on self-reported exposures (known occupational history, response time to WTC site, duration of work at site) and mostly PSG or home sleep apnea test (HSAT) measures of SDB. Some of the included studies of GWI focused on the categorization and case definition of GWI, which included sleep symptoms[112–114]. Of the WTC studies, a majority (5 of 7 studies) did not support an association between exposure duration or dose and SDB[115–119]; however, the largest study of responders (n=11,701 total) reported an association between arrival time and incident OSA, with earlier arrival time to the WTC site linked to higher odds of incident OSA[120]. Follow-up studies reported similar findings, with earlier response time and exposure to the dust cloud associated with greater log AHI[118] and severe, but not mild or moderate, OSA[121]. Among the two studies of sarin exposure, an OP compound, an older study by Duffy et al from 1979[122] reported differences in sleep architecture (as measured by EEG), such as increased REM sleep, among male workers occupationally exposed to sarin. Among civilians, sarin exposure was also linked to a higher frequency of insomnia, but not bad dreams, 3 years following a sarin attack, compared to unexposed[123]. The only study of sulfur mustard exposure thoroughly evaluated sleep health using both validated questionnaires (PSQI and STOP-Bang) and device-measured sleep (PSG), as well as serum melatonin, reporting detrimental effects of occupational sulfur mustard exposure on all of these outcomes[124]. Study quality was mixed (**Figure 3**), with overall supporting evidence between Gulf War exposures, sarin, and sulfur mustard with impaired sleep quality, insomnia, SDB, and altered sleep architecture.

**Figure 3.**
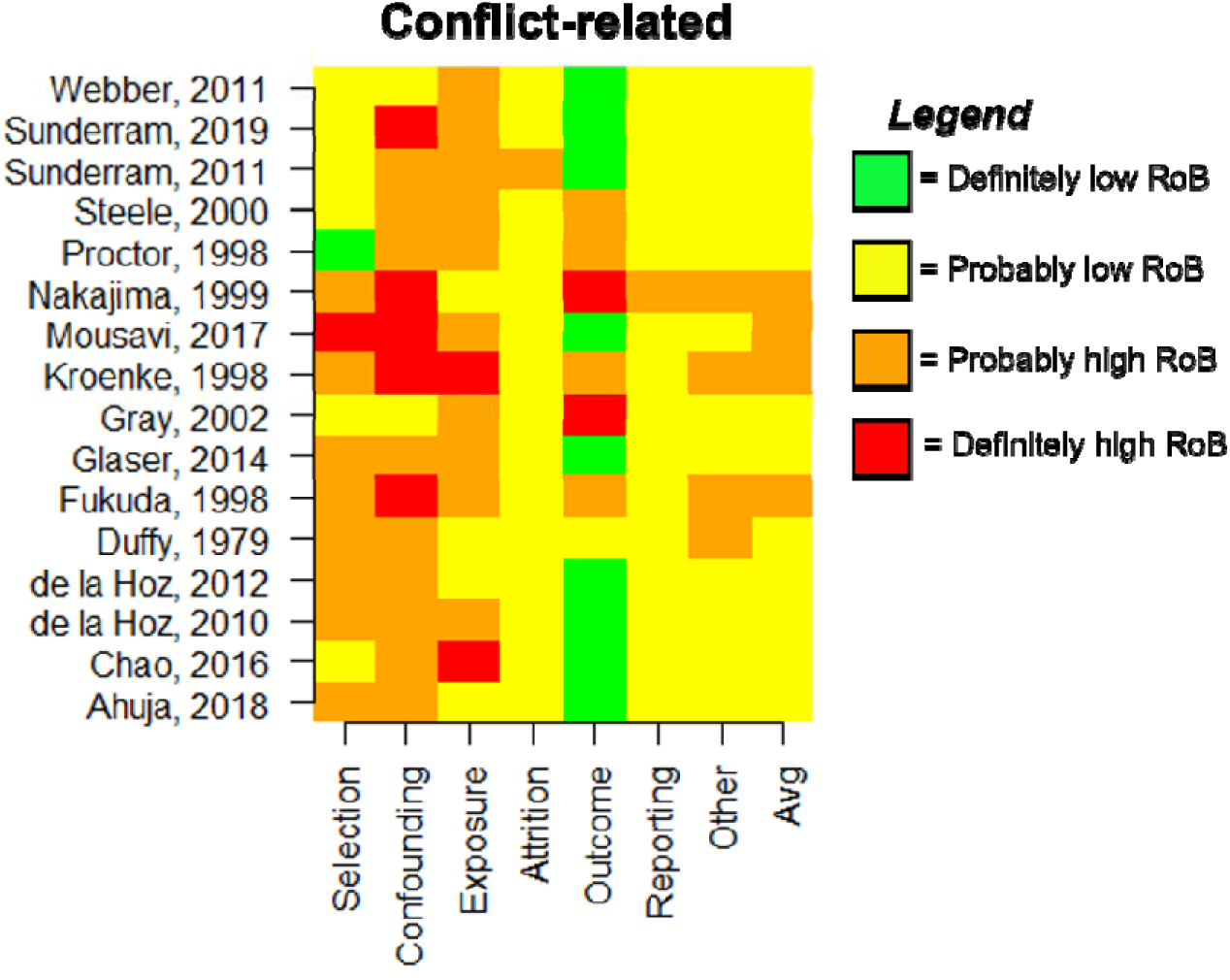
Chart depicting RoB ratings across evaluation domains for included epidemiologic studies on conflict-related chemical pollutant exposures.

**Table 5.** Summary table of epidemiologic studies of conflict-related exposures and sleep outcomes.

| First Author, Year | Design (Location) | Characteristics (age group) | Pollutant Measure (method) | Sleep Measure (method) | Findings | Direction<sup>†</sup> |
| --- | --- | --- | --- | --- | --- | --- |
| **World Trade Center-related 9/11 exposures** |  |  |  |  |  |  |
| Sunderram, 2019[115] | PC (North America) | N=601 patients from WTC responder clinics (Adults) | WTC dust (self-reported exposure based on duration and occupational history and classified as high, intermediate, or low) | OSA (HSAT, ARES Unicorder) | WTC responders with very high or intermediate exposure did not have higher prevalence of OSA compared to those with low exposure | — |
| Glaser, 2014[121] | CS (North America) | N=636 WTC FDNY responders, all males (Adults) | WTC dust (self-reported occupational history and arrival at WTC site within first 2 weeks; comparing responders who arrived on 9/11 versus responders who arrived over the next two weeks) | OSA, AHI (PSG) | Responders with earliest exposure to WTC (arrived on 9/11) had higher odds of severe (but not mild or moderate) OSA (OR=1.91, 1.15-3.17) compared to those who arrived within 2 weeks following | ▼ — |
| de la Hoz, 2010[116] | PC (North America) | N=100 WTC responders (Adults) | WTC dust (self-reported occupational history and arrival at WTC site within 48 hours) | OSA, AHI (PSG, Embla) | WTC exposure duration was not significantly associated with AHI | — |
| de la Hoz, 2012[117] | CS (North America) | N=272 exposed WTC responders, N=384 unexposed (Adults) | WTC dust (self-reported occupational history and treatment sought at WTC clinic) | OSA, AHI, REM-related OSA, UARS (PSG, Embla) | WTC exposure was not associated with OSA or any of the other PSG diagnoses (SDB, REM-related OSA, UARS) | — |
| Ahuja, 2018[118] | CS (North America) | N=143 exposed WTC responders (Adults) | WTC dust (self-report of degree of exposure to dust cloud) | SDB/OSA, AHI (PSG in lab or HSAT) | WTC exposure was not associated with OSA diagnosis, but exposure to dust cloud was positively associated with log AHI $\geq 4\%$ | ▼ — |
| Webber, 2011[120] | PC (North America) | N=11,701 total exposed WTC responders; N=1,879 arrival group 1, N=7,165 arrival group 2, N=1,477 arrival group 3, N=1,180 arrival group 4 (Adults) | WTC dust (self-report of occupational exposure and FDNY-WTC Exposure Intensity Index) | OSA (modified Berlin questionnaire for OSA risk and self-reported physician-diagnosed OSA) | Responders in arrival groups 1 (OR=2.24, 1.47-3.41), 2 (OR=2.06, 1.42-2.99), and 3 (OR=1.86, 1.19-2.92) had higher odds of incident OSA (but not prevalent OSA) at follow up compared to those in group 4. Responders with $\geq 4$ months work duration at the WTC site had higher odds of incident OSA at follow-up (OR=1.26, 1.05-1.52) | ▼ |
| Sunderram, 2011[119] | CS (North America) | N=50 exposed male WTC responders, N=50 unexposed (Adults) | WTC dust (occupational history) | AHI measures, sleep quality (PSG and questionnaire for sleep quality) | Responders had worse sleep quality (p=0.046) compared to non-responders (p=0.12) but lower AHI (22.7 vs 43.4, p=0.0004) and fewer apneas (28.7 vs 43.7, p=0.023) | ▼ — |
| **Exposures related to serving in the Gulf War** |  |  |  |  |  |  |
| Kroenke, 1998[113] | CS (North America) | N=18,495 Gulf War veterans (Adults) | Gulf War exposures (questionnaire and occupational history) | Sleep disturbance (questionnaire asking about sleep problems) | Among Gulf War veterans, prevalence of sleep problems ~3% before the war and ~33.5-17% during and after the war; approximately 33% of Gulf War veterans reported sleep problems. Prevalence of self-reported sleep problems was 33% in participants exposed to diesel fuel, tent heater fumes, and/or pyridostigmine, 34% in participants exposed to oil-fire smoke, solvents, CARC paint, and/or pesticides, 39% of participants exposed to mustard gas, and 40% of participants exposed to nerve gas | ▼ |
| Chao, 2016[125] | CS (North America) | N=98 Gulf War veterans; N=77 | Gulf War exposures (Kansas Gulf War | Sleep quality (PSQI), | Compared to Gulf War veterans without GWI, | ▼ — |
|  |  | cases and N=21 controls by CDC case definition, N=47 cases and 26 controls by Kansas definition (Adults) | Military History and Health Questionnaire for GWI status) | SDB/OSA (OSA STOP score), insomnia (insomnia severity index), sleep disturbance | those with GWI had higher STOP scores, and predicted nerve agent exposure was sig associated with STOP scores (but other covariates were null)(Spearman's: 0.73, p<0.001); GWI veterans had greater insomnia and worse sleep PSQI, but these were sig associated with PTSD (not nerve agent) |  |
| Fukuda, 1998[112] | CS (North America) | N=1,163 Gulf War veterans and N=2,538 non-deployed personnel (Adults) | Gulf War exposures (deployment record, occupational history) | Difficulty sleeping (questionnaire) | Air force Gulf war veterans had higher prevalence of sleep disturbance than nondeployed (self-calculated unOR=2.6, 2.19-3.10) | ▼ |
| Gray, 2002[126] | CS (North America) | N=3,831 Gulf War deployed Seabees, N=8,037 Seabees non-deployed or deployed elsewhere (Adults) | Gulf War exposures (deployment history, occupational history) | Trouble sleeping (questionnaire) | Seabees deployed in the Gulf War had higher prevalence of trouble sleeping post deployment compared to deployed elsewhere (OR=3.02, 2.72-3.35) and nondeployed Seabees (OR=3.08, 2.71-3.50) | ▼ |
| Proctor, 1998[127] | CS (North America) | N=252 exposed veterans, N=48 unexposed (Adults) | Gulf War exposures (Health Symptom checklist questionnaire, deployment) | Inability to fall asleep (Expanded Health Symptom Checklist) | Higher prevalence of insomnia among Gulf-deployed than Germany deployed veterans (OR=3.4 and 3.6, no CI provided but stated it excludes 1) | ▼ |
| Steele, 2000[114] | CS (North America) | N=1,435 exposed veterans, N=409 unexposed (Adults) | Gulf War exposures (deployment history, occupational history) | Problems falling or staying asleep (interview) | Gulf War veterans had higher prevalence of problems falling or staying asleep compared to non-PGW veterans (OR=2.98, 2.18-4.08) | ▼ |
| **Other** |  |  |  |  |  |  |
| Duffy, 1979[122] | CS (North America) | N=77 exposed workers, N=38 unexposed (Adults) | Sarin (occupational history of at least 1 exposure) | Architecture (EEG recorded via Ampex FR 1300 tape deck) | Exposure associated with greater time in REM sleep, both as absolute duration and as a percentage of total sleep | ▲ |
| Nakajima, 1999[123] | PC (Asia) | N=318 exposed (N=167 with 3-year data), N=919 unexposed (N=669 with 3-year data) (Adults) | Sarin (self-reported exposure and vicinity to attack) | Insomnia, bad dreams (questionnaire) | Exposure was not associated with cholinesterase activity between those who reported bad dreams or insomnia; when measured 3 years after the incident, sarin-exposed participants (n=167) had slightly higher frequency of self-reported insomnia, but not bad dreams, compared to non-exposed (n=669)(OR=2.48, CI:1.07-5.78) | ▼ — |
| Mousavi, 2017[124] | CS (Middle East) | N=30 exposed, N=10 unexposed males (Adults) | Sulfur Mustard (occupational history, documented exposure) | Sleep quality (PSQI), AHI, ODI, O<sub>2</sub> saturation, RDI (PSG, ALICE 5 Health dyne, and STOP-Bang questionnaire), AHI (PSG), melatonin levels (serum melatonin with ELISA) | Exposure associated with greater mean PSQI score (11.76 vs. 2.78, p<0.001), STOP-Bang score (5.03 vs. 1.18, p=0.003), AHI (20.06 vs. 3.42, p<0.001), lower O<sub>2</sub> saturation (78 vs 95, p<0.0001), higher ODI (37.19 vs. 4.44, p=0.003), and lower serum melatonin (29 vs 78, p=0.005) | ▼ |
| <sup>†</sup>Downward pointing red triangle symbol represents sleep disruption, black dash symbol represents null finding, and upward pointing blue triangle represents improved sleep outcome (not necessarily indicative of a healthy outcome) in relation to exposure.<br>Abbreviations: AAS=atomic absorption spectrometry; AHI=apnea-hypopnea index; CS=cross-sectional; E=ecological; EEG=electroencephalography; ELISA=enzyme-linked immunoassay; FDNY=New York City Fire Department; GWI=Gulf War Illness; HPLC=high-performance liquid chromatography; HSAT=Home Sleep Apnea Test; ICP-MS=inductively coupled mass spectrometry; N=number; NHANES=National Health and Nutrition Examination Survey; PC=prospective cohort; PGW=Persian Gulf War; PSG=polysomnography; PSQI=Pittsburgh Sleep Quality Index; ODI=Oxygen Desaturation Index; OR=odds ratio; SDB=sleep-disordered breathing; UARS=Upper Airway Resistance Syndrome; WTC=World Trade Center |  |  |  |  |  |  |

### Endocrine Disrupting Chemicals (EDCs) / Other

This category included 12 epidemiologic studies and 3 case studies of EDCs and other chemicals that weren’t well captured by other categories. Pollutants included: bisphenol A (BPA), dioxins, per- and polyfluoroalkyl substances (PFAS), phthalates, polybrominated diphenyl ethers (PBDEs), and polychlorinated biphenyls (PCBs). Among epidemiologic studies, the most common sources of exposure to this class were unknown or occupational, and most relied on questionnaire data for outcome measurement (**Table 6**). Among pollutants represented by more than one study, BPA was reported to be protective of short sleep (continuous or dichotomized) in an NHANES analysis, but this relationship did not remain across BPA quartiles[128]; in another study, higher BPA levels were reported in patients with OSA compared to adults without OSA[129]. In a prospective birth cohort, female, but not male, children with the highest tertile of cord-blood measured PBDE had greater problems sleeping compared to those in the lowest tertile at four years old[130]. PCBs and dioxins were also linked to problems sleeping[131, 132], although in one case, exposure was due to a fire, and stress may be a confounder in the association[132]. Of the two studies of phthalates exposure, an analysis in the Midlife Women’s Health Study did not report an association with insomnia or sleep disturbance except in former smokers; an analysis of NHANES participants ages 16-17 did report higher odds for short (<8 hours) weekday sleep duration among those in the highest quartile of phthalate exposure, but did not consider school start times or the influence of puberty and chronotype on the outcome[133]. An analysis of 140 people exposed to PCBs from contaminated rice bran reported worse sleep quality and higher prevalence of difficulty initiating sleep among those in the highest quartile of exposure[134]. Case studies of residents living in methamphetamine-contaminated homes [135] and homes with off-gassing from polyurethane foam insulation[136] highlighted reports of sleep disturbance and insomnia. The overall quality of studies varied (**Figure 4, Supplemental Figure 1, Table 7**), with no clear consistent associations, possibly compounded by the heterogeneity of the pollutants. However, persistent EDCs such as dioxins, PCBs, and PBDEs showed the strongest evidence for sleep disruption.

**Figure 4.**
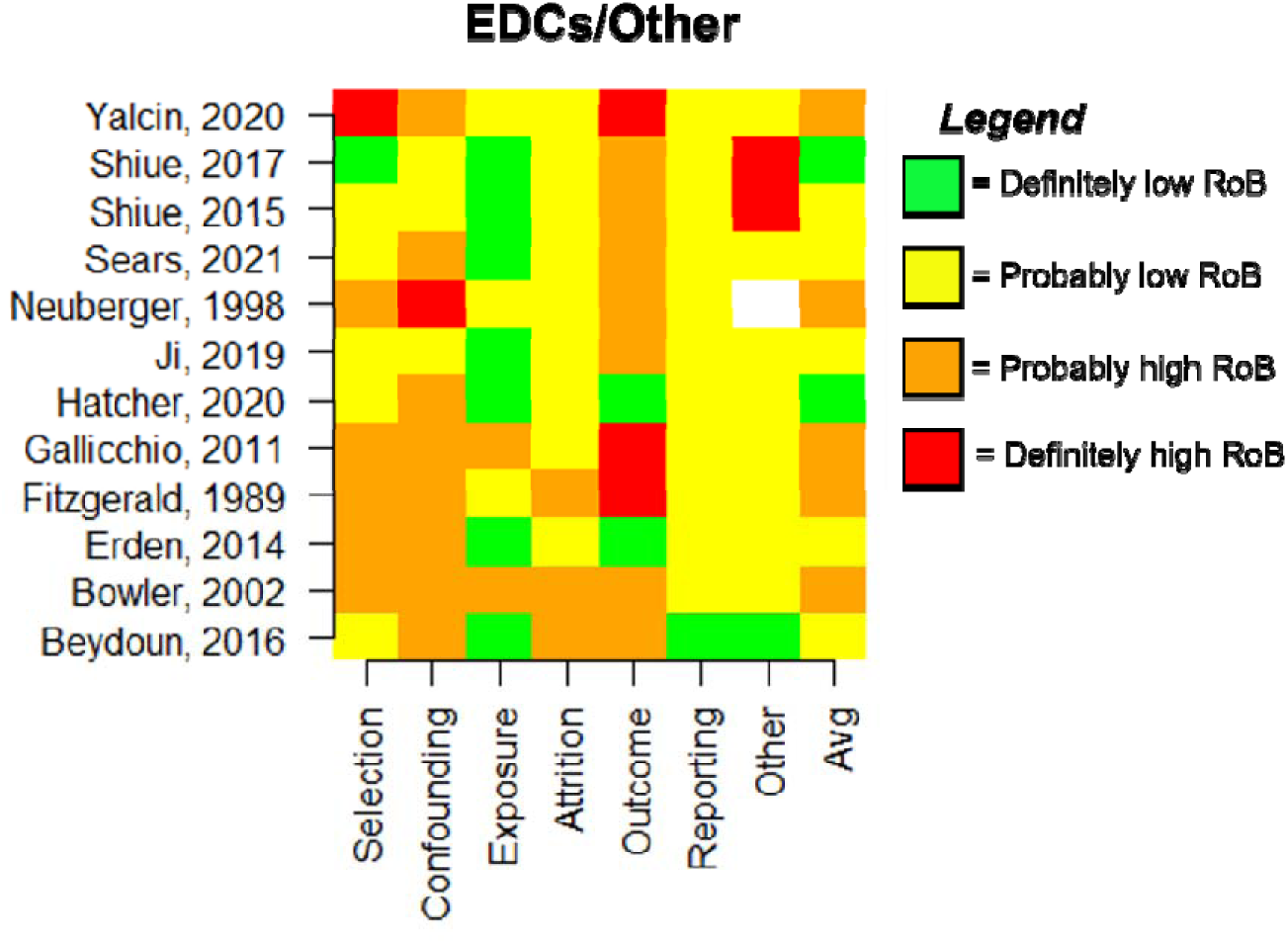
Chart depicting RoB ratings across evaluation domains for included epidemiologic studies on EDCs and pollutants not captured by other categories.

**Table 6.**
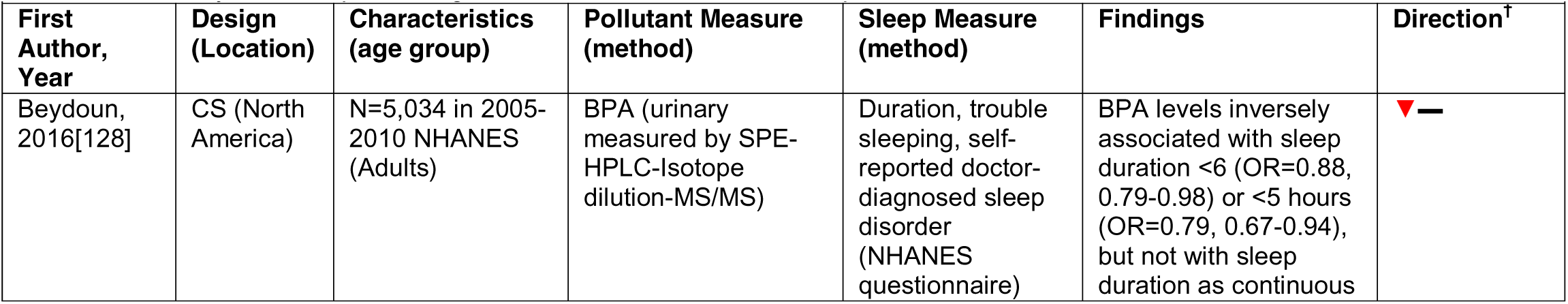
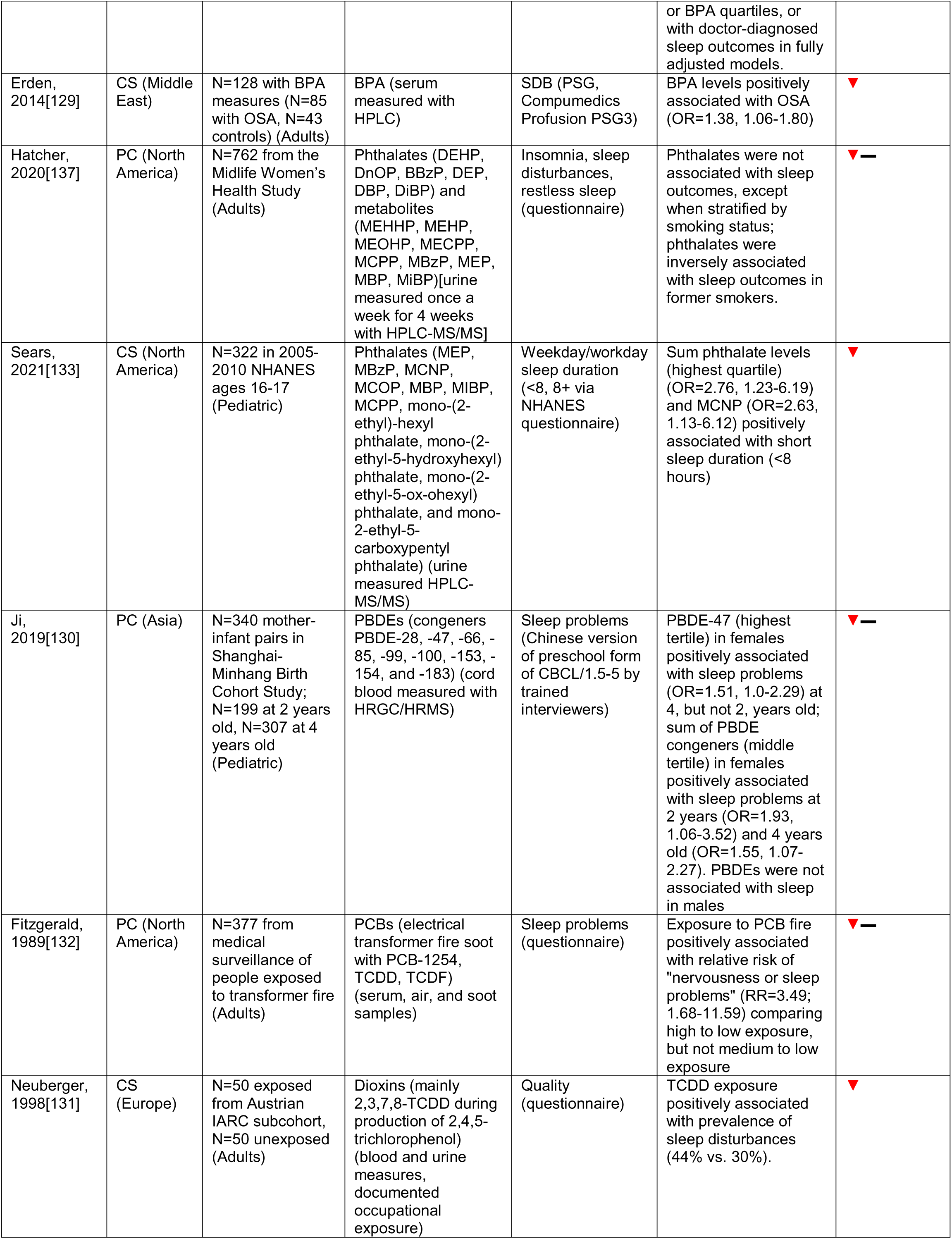
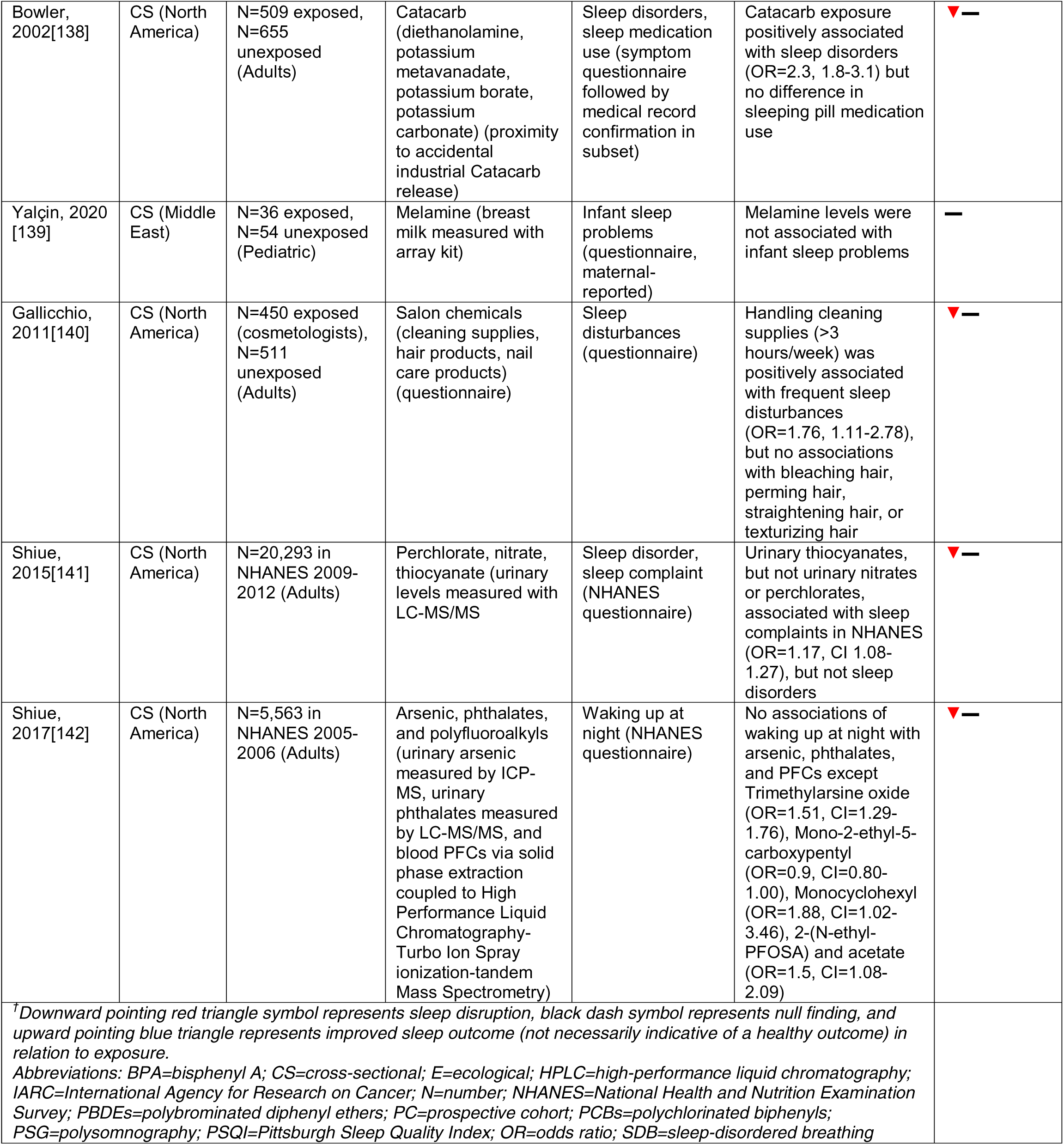
Summary table of epidemiologic studies of EDCs/other and sleep outcomes.

**Table 7.** Summary table of case studies / case series of EDCs/other and sleep outcomes.

|  |  |  |  |  |  |
| --- | --- | --- | --- | --- | --- |
| First Author, Year | Location | Characteristics (age group) | Pollutant Measure (method) | Sleep Measure (method)* | Findings |
| Kondo, 2018[134] | Asia | N=140 ate contaminated rice bran (Adults) | PCBs (such as 2,3,4,7,8,-PeCDF) (blood measured every few years) | Insomnia (Insomnia Severity Index, self-report), | PeCDF not associated with SDB, but PeCDF (fourth quartile) associated with PSQI ≥ 8 (OR=4.84, 1.10-21.25) (but |
| | | | | quality (PSQI), SDB (pulse oximetry) | not PSQI $\geq 6$) and higher prevalence of difficulty initiating sleep; overall evidence is good |
| Huang, 2014[136] | North America | N=13 across 10 households, N=5 females aged 33-57, N=8 males aged 35-82 (Adult) | Spray polyurethane foam for housing insulation (known application and some with indoor air sampling for VOCs, measured with HPLC) | Insomnia (self-report) | VOCs present in all households, insomnia experienced by 12/13 people, sampling taken before and after foam removal confirmed source; overall evidence is fair |
| Wright, 2020[135] | Oceania / Australia | N=65 people across 25 case studies, N=37 adults, N=28 children, inadvertently exposed after moving into contaminated residence (Both) | Methamphetamine residue from drug manufacture and/or use (surface wiping tests and hair analysis) | Quality, difficulty sleeping (self-report) | N=23/37 adults and N=21/28 children experienced sleep impairment after exposure; symptoms resolved after changing residence; overall evidence is good |

*\*Note: method of self-report for sleep outcomes were not always explicitly stated in case studies, symptoms assumed reported to physician unless stated otherwise.* Abbreviations: HPLC=high throughput liquid chromatography; N=number; PCBs=polychlorinated biphenyls; PSQI=Pittsburgh Sleep Quality Index; SDB=sleep-disordered breathing; VOCs=volatile organic compounds

### Metals

There were 51 epidemiologic studies and 34 case studies of exposure to metals included in the results. Most focused on exposure to lead or mercury, but a few also investigated exposure to aluminum, antimony, arsenic, cadmium, coal ash, copper, fluoride, lead, manganese, mercury, selenium, and thallium (**Table 8**). The most common sources were workplace or occupational, particularly welding and/or metallurgy work, as well as dental work and amalgams. Among metals represented by more than one epidemiologic study, working with aluminum was linked to insomnia[143, 144]. Studies of mercury and insomnia or sleep quality were inconsistent; however, positive associations were more often reported with higher occupational exposure to mercury[145, 146], whereas most studies of dental amalgams and dentistry workers reported null associations[147–150]. In both adult and pediatric populations, exposure to lead was most consistently associated with insomnia[151–153] and shorter sleep duration[154–156], whereas associations with sleep quality were mixed. Thallium exposure was also linked to sleep disruption[157–159], although some of these findings may be confounded by smoking. The evidence between metals exposure and SDB were inconsistent. Associations between arsenic and sleep were null[160, 161], and the majority of studies did not support an association between metals exposure and sleep duration in adult populations (**Figure 5)**. Findings from the pediatric literature was also inconsistent, with most studies of sleep quality reporting null associations and mixed associations with sleep timing, with positive findings in relation to self-reported sleep[156, 162] and null findings with actigraphy measured sleep[163, 164]. In contrast to the epidemiologic literature, most case studies in adults and children reported an association with insomnia and sleep quality and were of fair or good quality (**Table 9, Supplemental Figure 1**). A case study with multiple biological measures of mercury exposure and environmental sampling reported sleep disturbances among 70% of 18 adults living in a town polluted by mercury due to gold mining[165]. Other case studies reported symptoms of sleep disturbance among male welders following exposure to lead[166, 167], manganese[168, 169], and other metals from welding fumes. Overall, findings supported a link between exposure to mercury or lead and insomnia.

**Figure 5.**
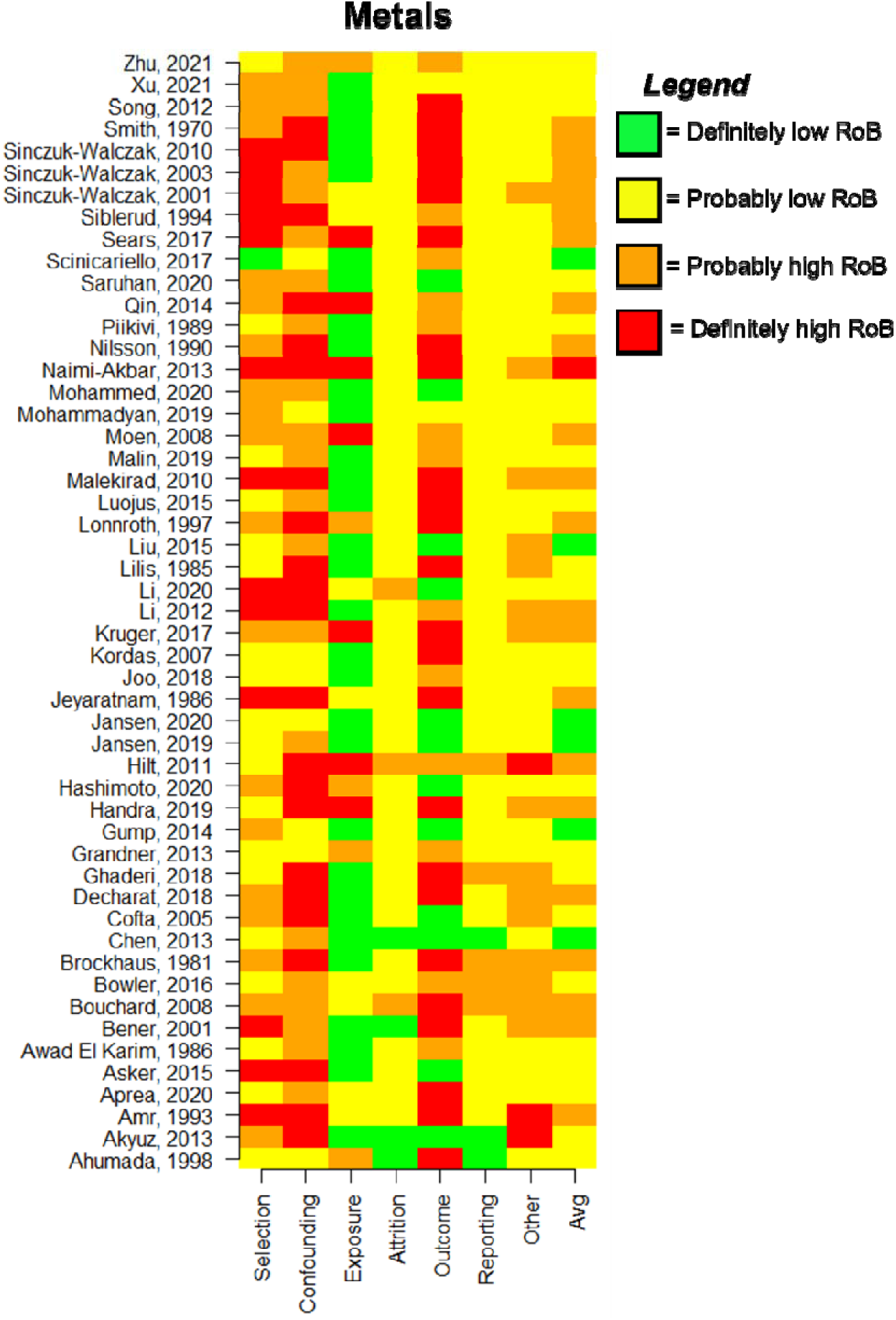
Chart depicting RoB ratings across evaluation domains for included epidemiologic studies on metals.

**Table 8.**
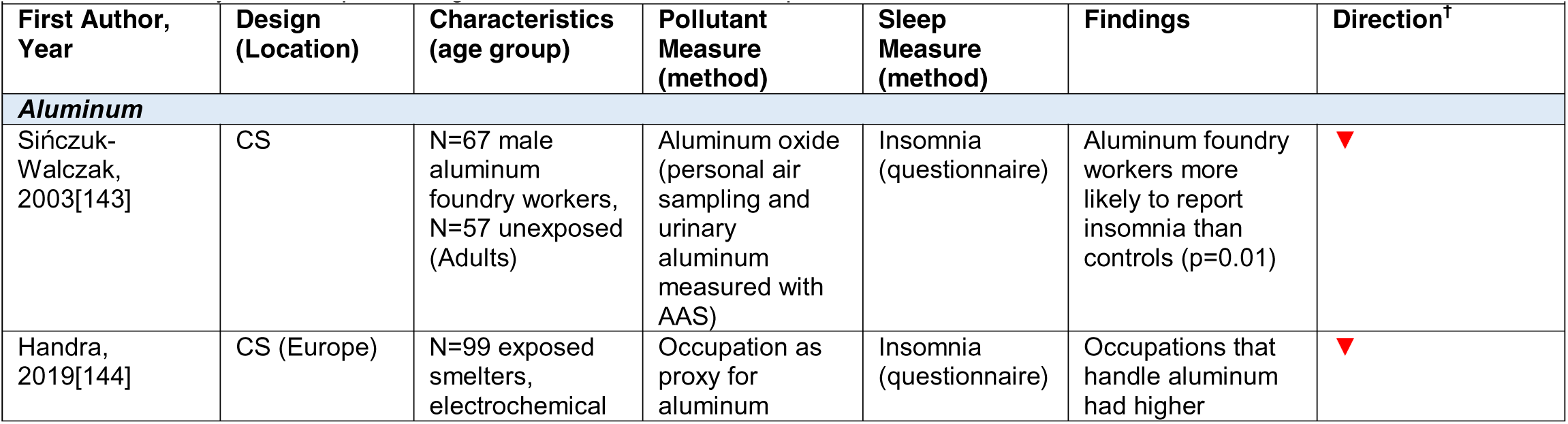
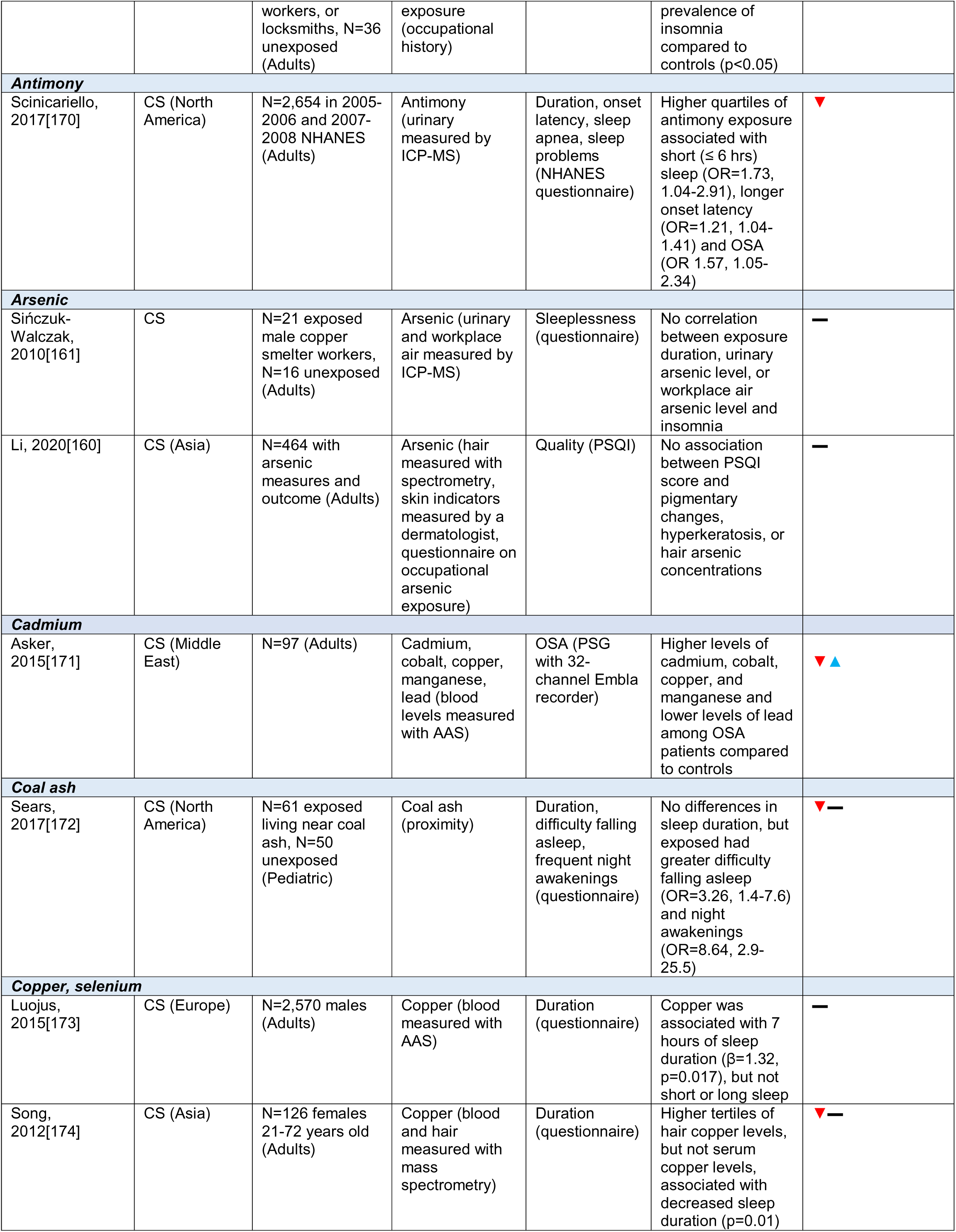
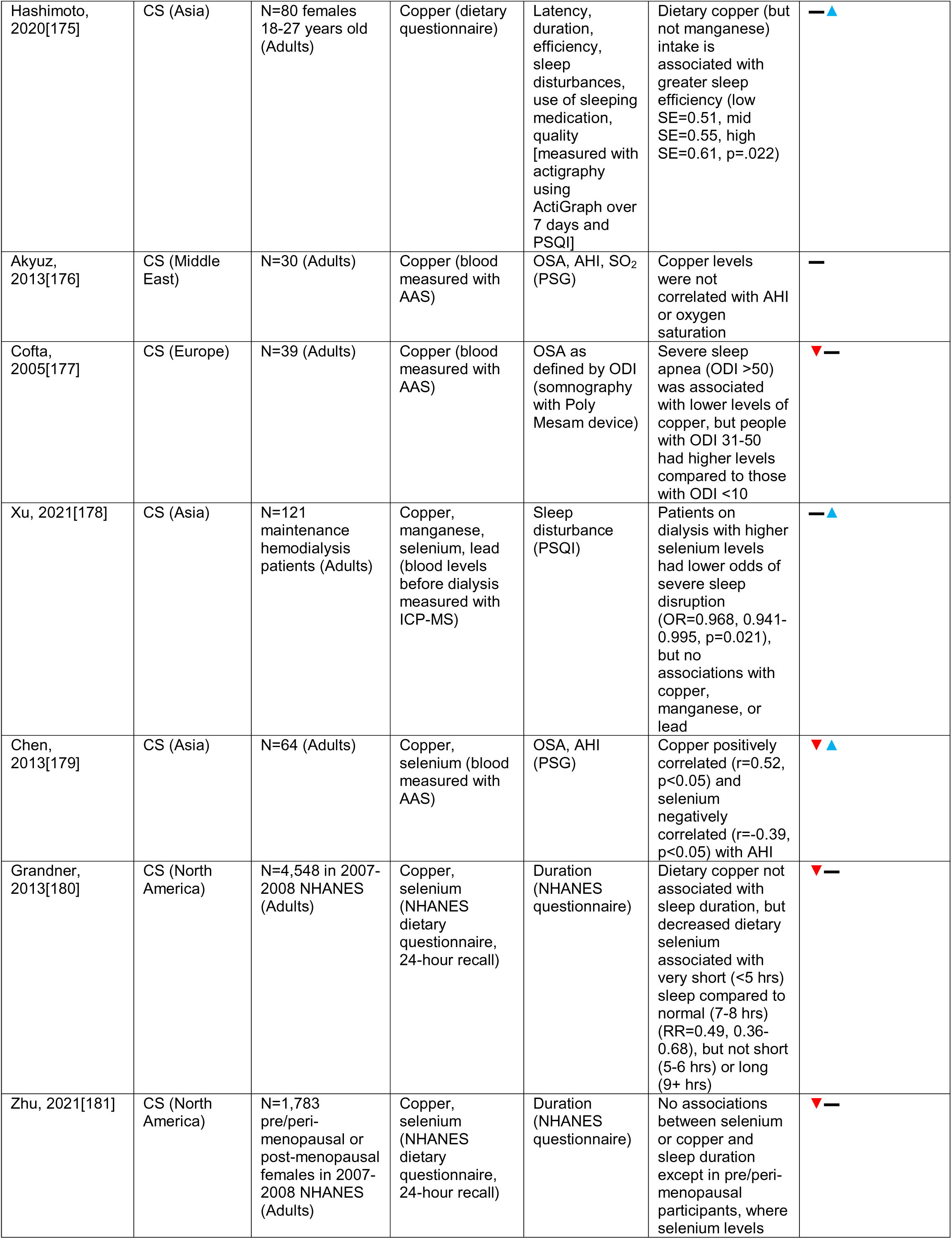
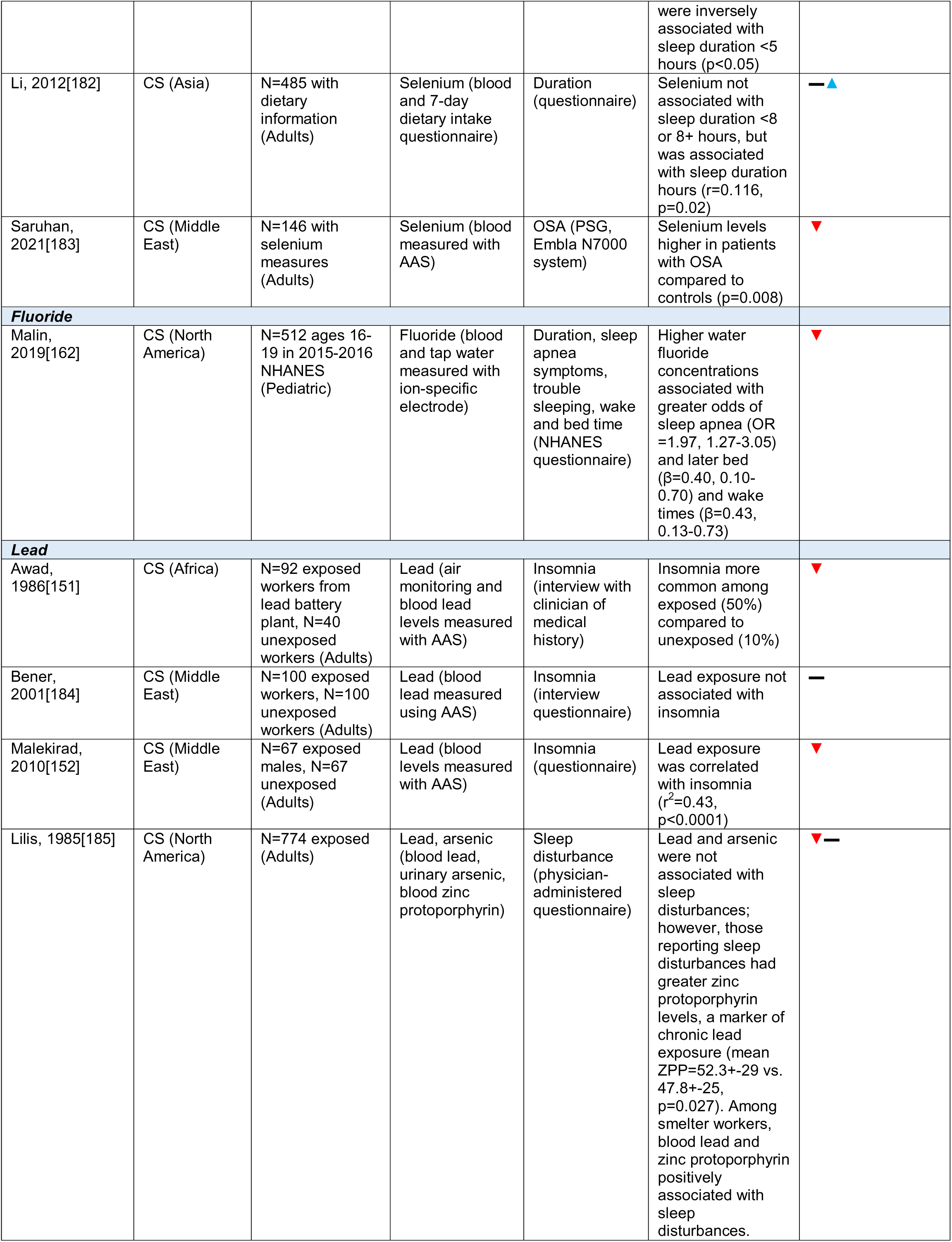
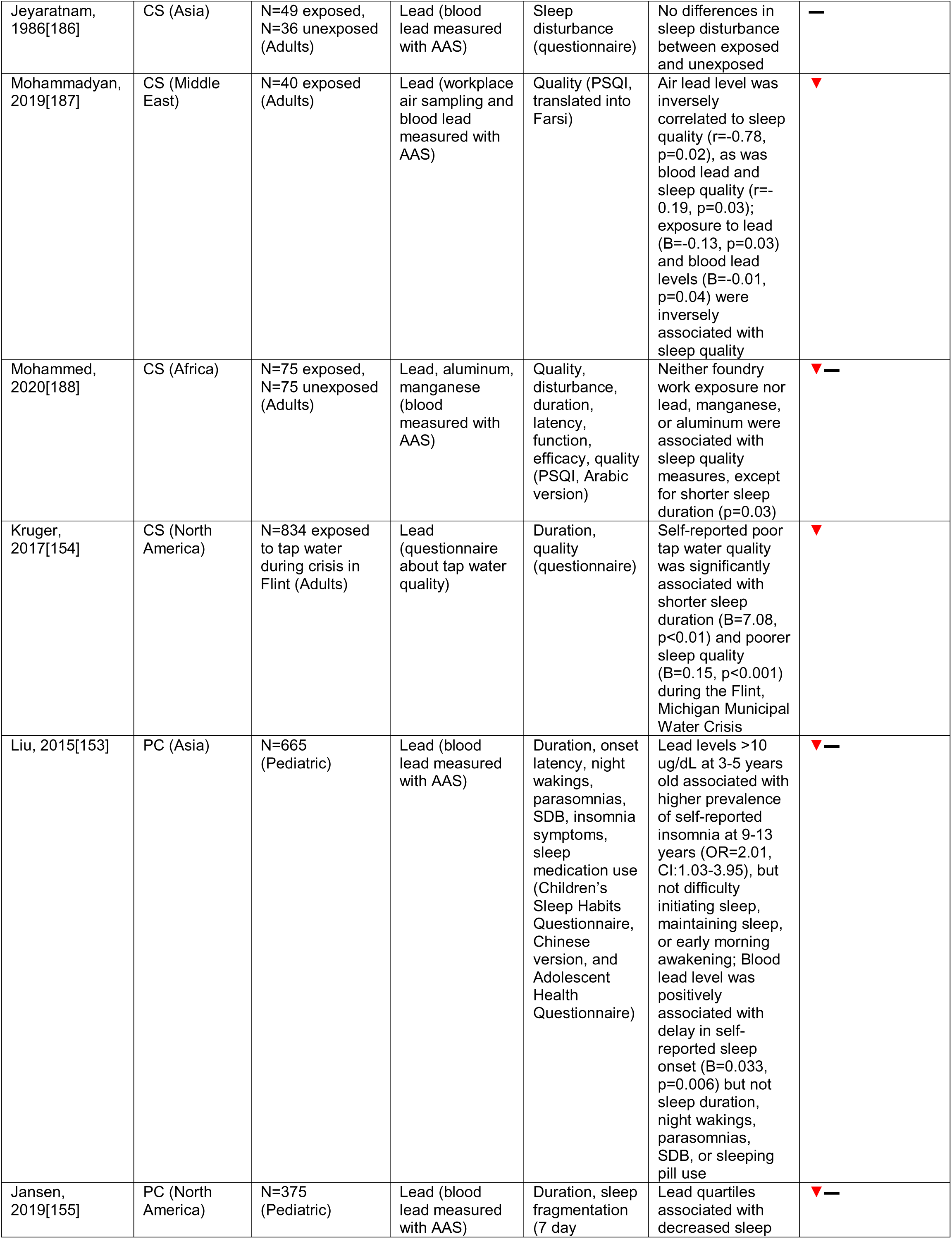
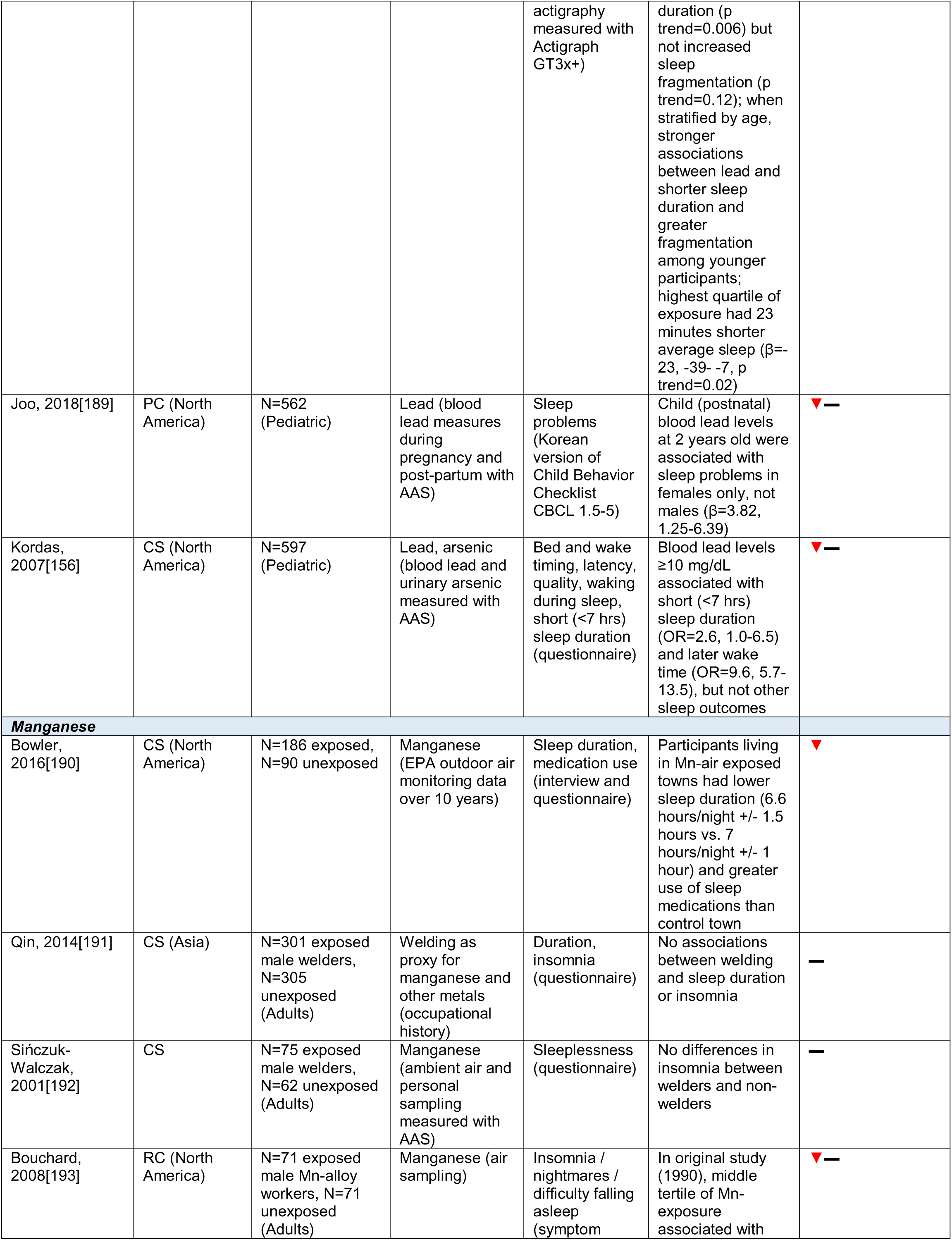
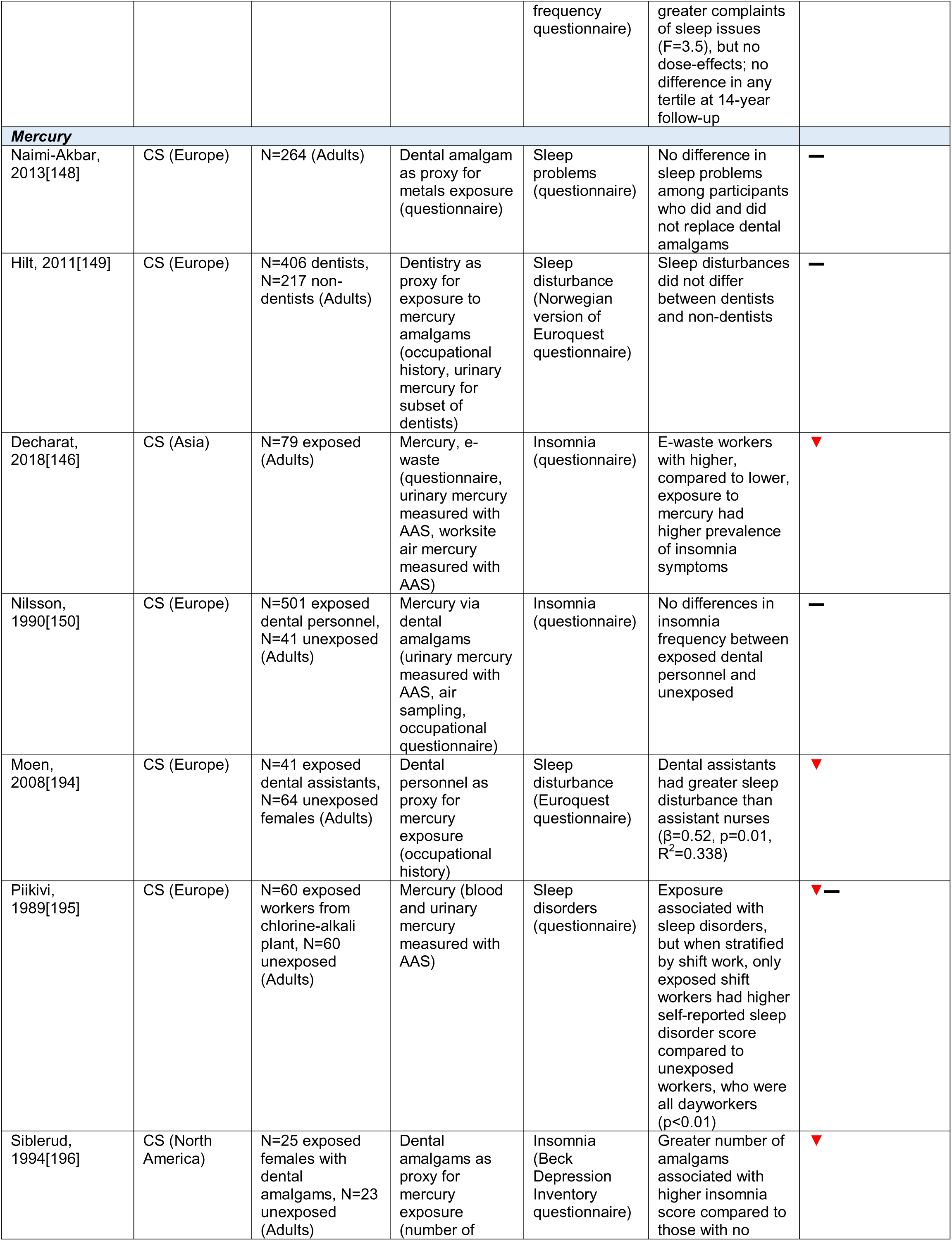
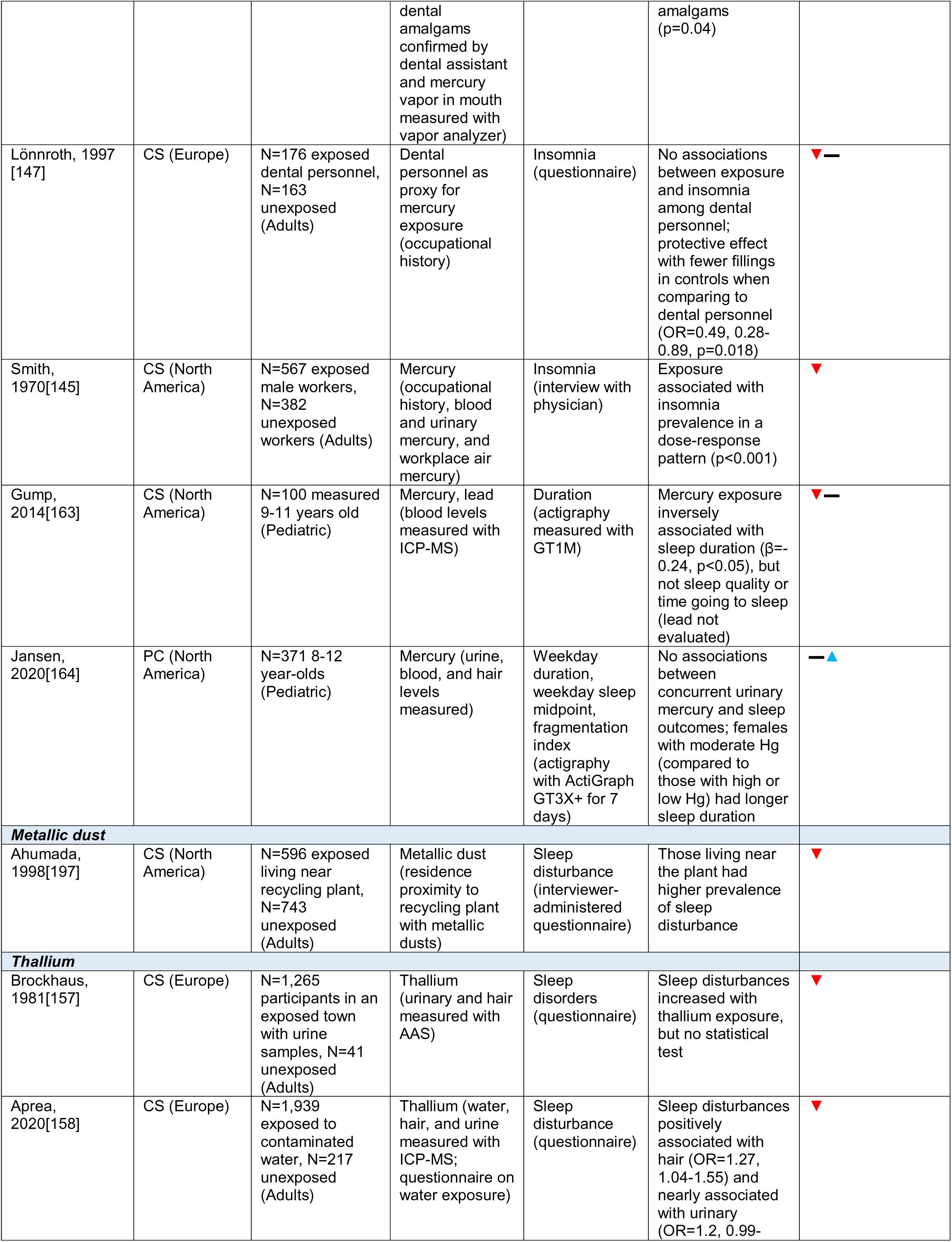
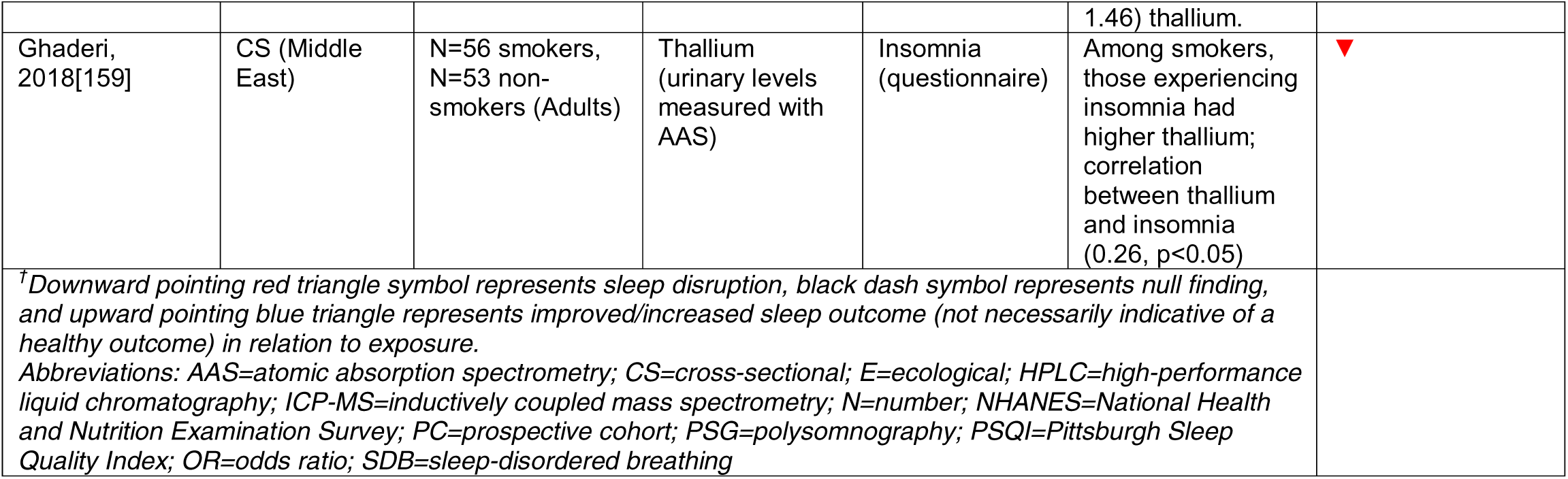
Summary table of epidemiologic studies of metals and sleep outcomes

**Table 9.**
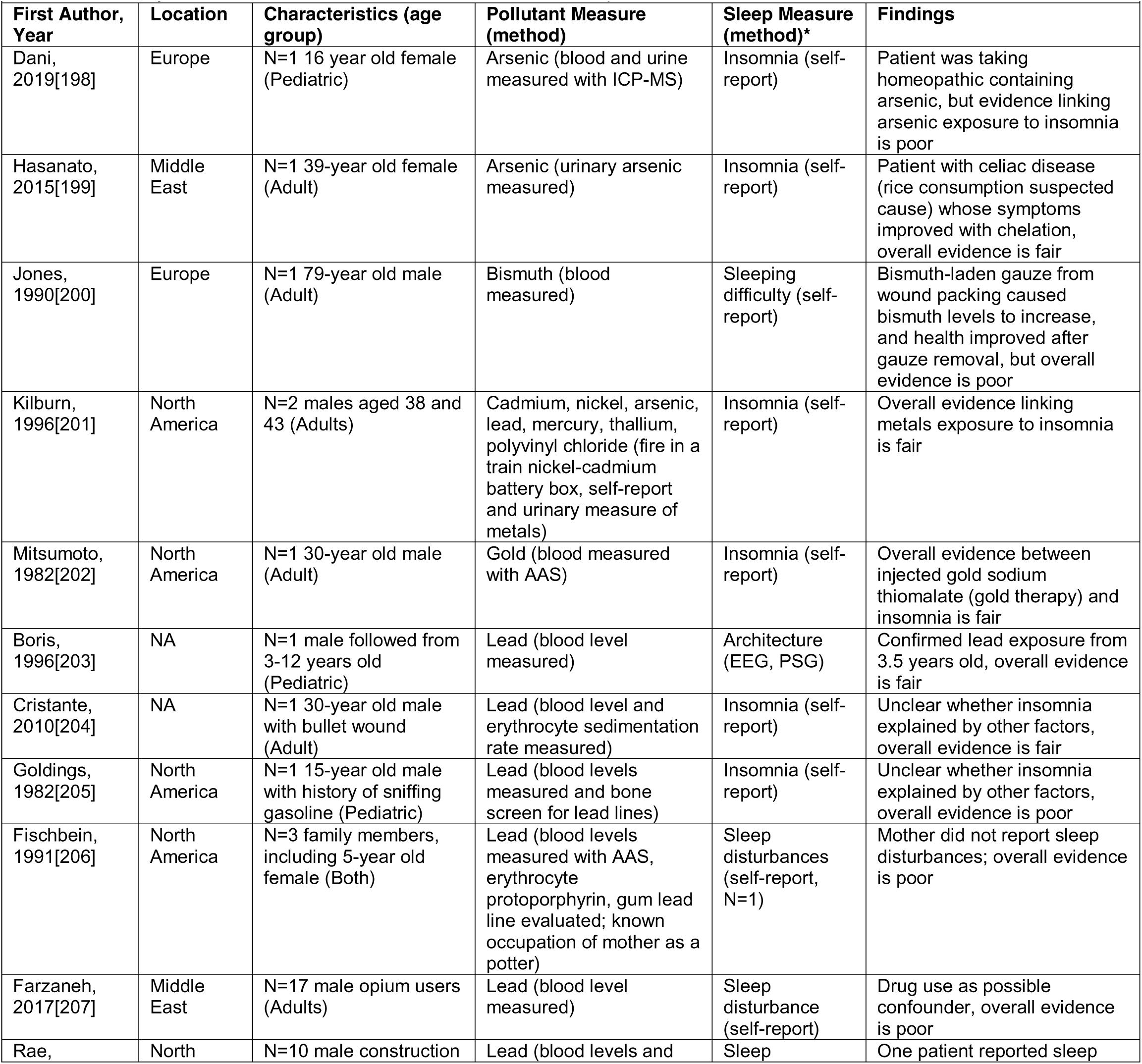
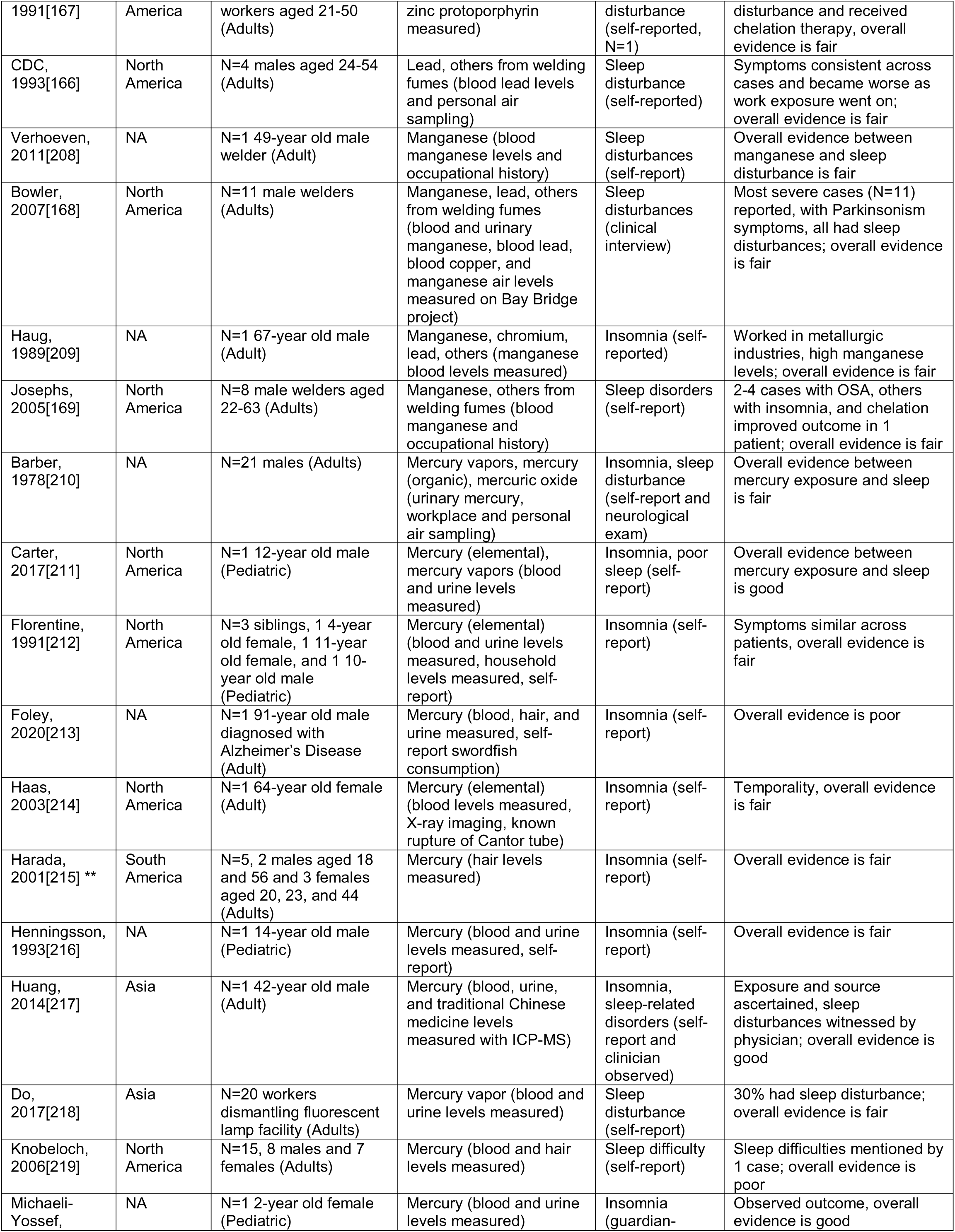
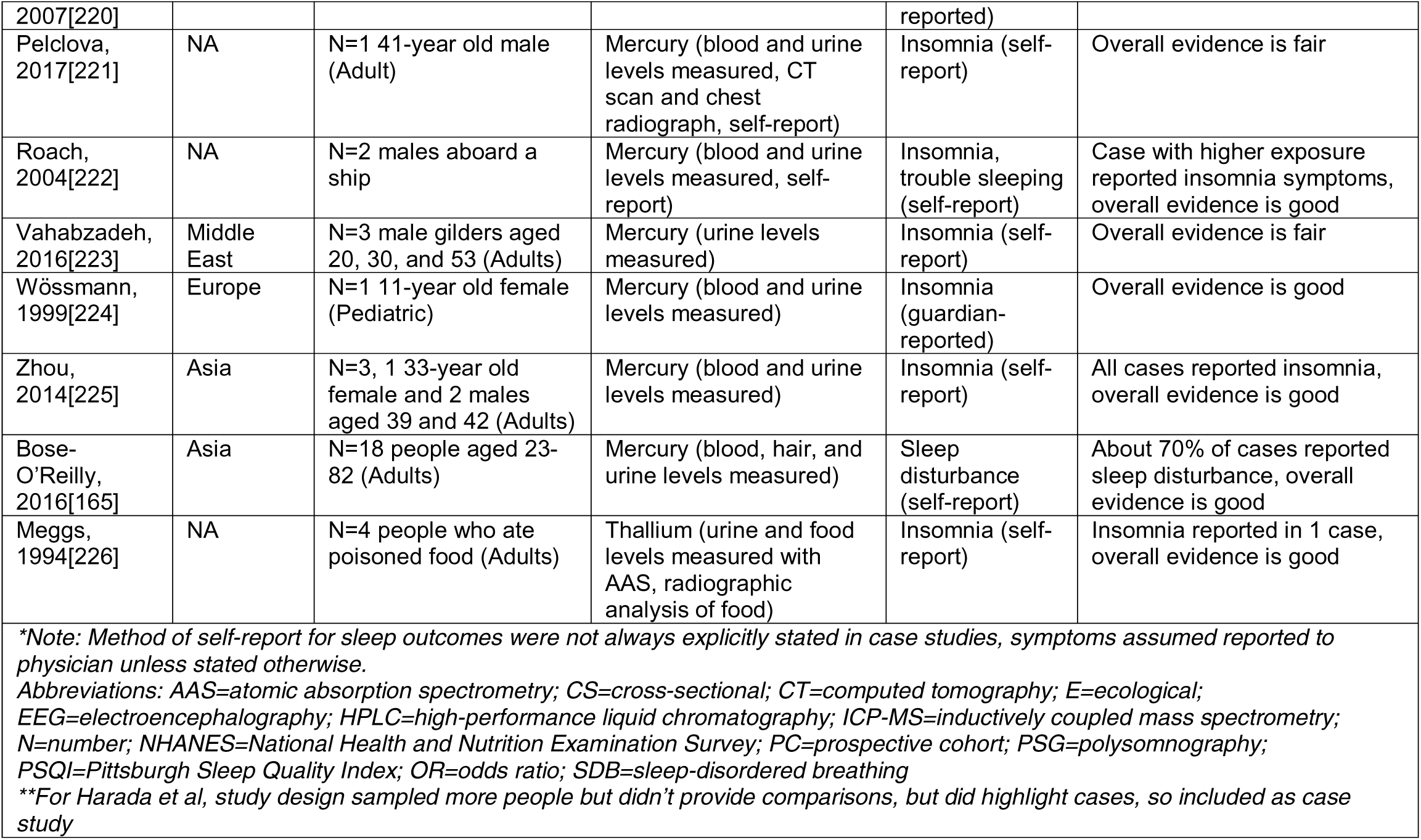
Summary table of case studies / case series of metals and sleep outcomes.

### Pesticides

Of the 12 epidemiologic and 5 case studies of pesticide exposure, most included pesticides in the organophosphate (OP), organochlorine, and/or carbamate class (**Table 10**) from a rural/farmland source in the Middle East, Africa, or North America and had a small sample size. Some of the included studies relied on farm work as a proxy for pesticide use and exposure, while others did not explicitly state the pesticide class or chemical(s) evaluated. All of the studies relied on self-reported pesticide use and/or blood measures of cholinesterase and all but one[227] utilized self-reported measures of sleep. One study comparing female pickers of Bt cotton, a genetically modified crop that may require less pesticide use, and non-Bt cotton reported higher frequency of insomnia among non-Bt pickers (12% to 39%)[228]; however, in reporting frequencies rather than adjusted model results for insomnia, factors such as socioeconomic status were not evaluated in this association. In another study, male pesticide applicators in the Agricultural Lung Health Study with self-reported exposure to carbamates, but not OPs, had higher self-reported doctor-diagnosed sleep apnea, with or without adjustment for BMI[229]. In a cross-sectional study of 1,336 greenhouse farmers exposed to pesticides, farmers with both medium and high cumulative exposure to pesticides had worse sleep quality and more trouble falling asleep compared to famers with low cumulative exposure; the highly exposed farmers also had 56% higher odds of short (≤6 hours) sleep duration compared to those with low exposure[230]; however, results were not differentiated by pesticide class. An investigation by Rubin et al of households in Ohio that had been illegally treated with an OP compound reported 28% of participants in the contaminated households experienced night waking, and in one case a 4 month old infant with disrupted sleep was brought to the emergency room after their sleeping area was sprayed[231]. In another case, an adult male developed sleep apnea after their home was improperly treated with Carbaryl, a carbamate compound[232]. None of the included studies specifically evaluated a pediatric population. Most epidemiologic studies of pesticide exposure and insomnia reported an association[228,233–235], whereas associations with sleep duration, quality, and SDB were inconclusive. In summary, the overall epidemiologic evidence between pesticide exposure and sleep disruption was of low quality (**Figure 6**), in contrast with the overall evidence in case studies, which was good (**Table 11, Supplemental Figure 1**). Insomnia was the sleep outcome most consistently associated with pesticide exposure.

**Figure 6.**
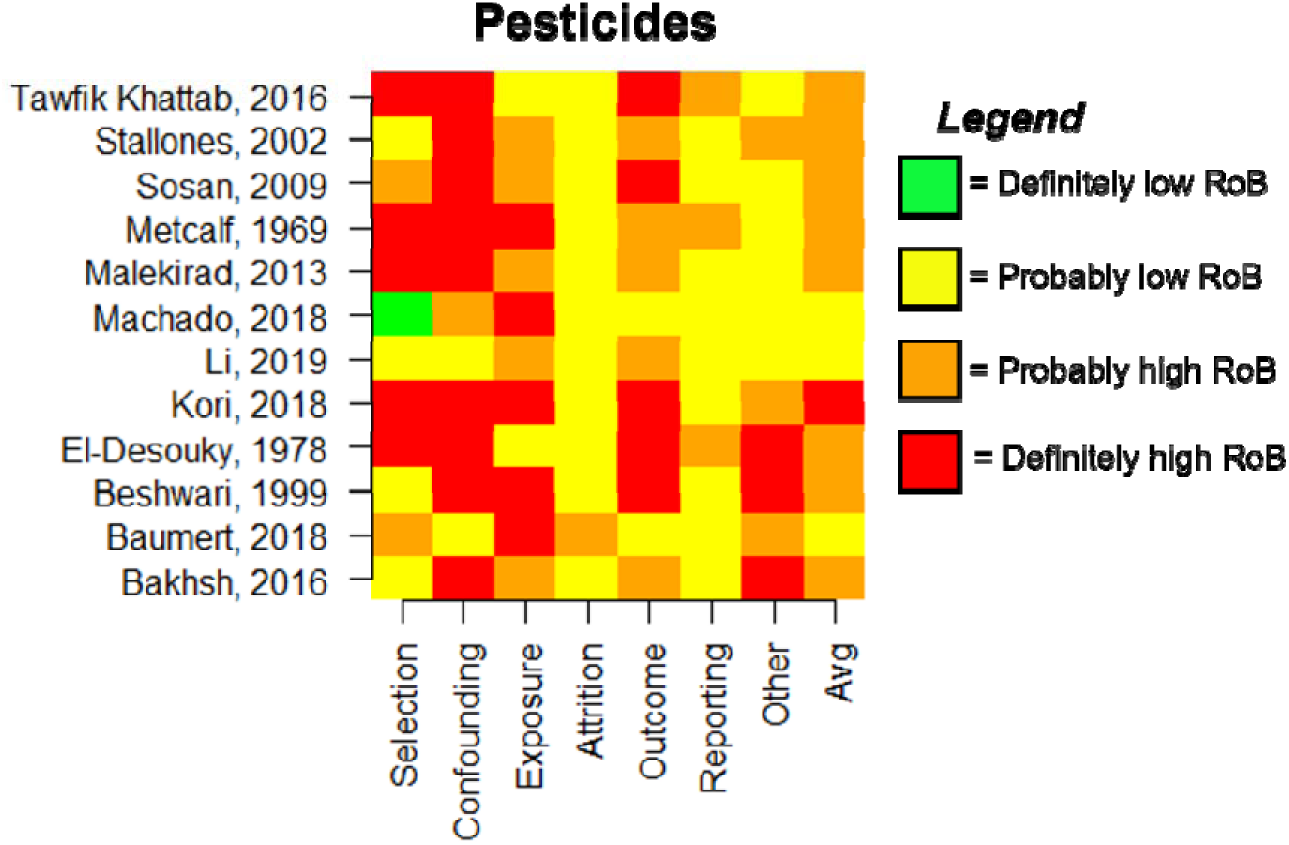
Chart depicting RoB ratings across evaluation domains for included epidemiologic studies on pesticides.

**Table 10.**
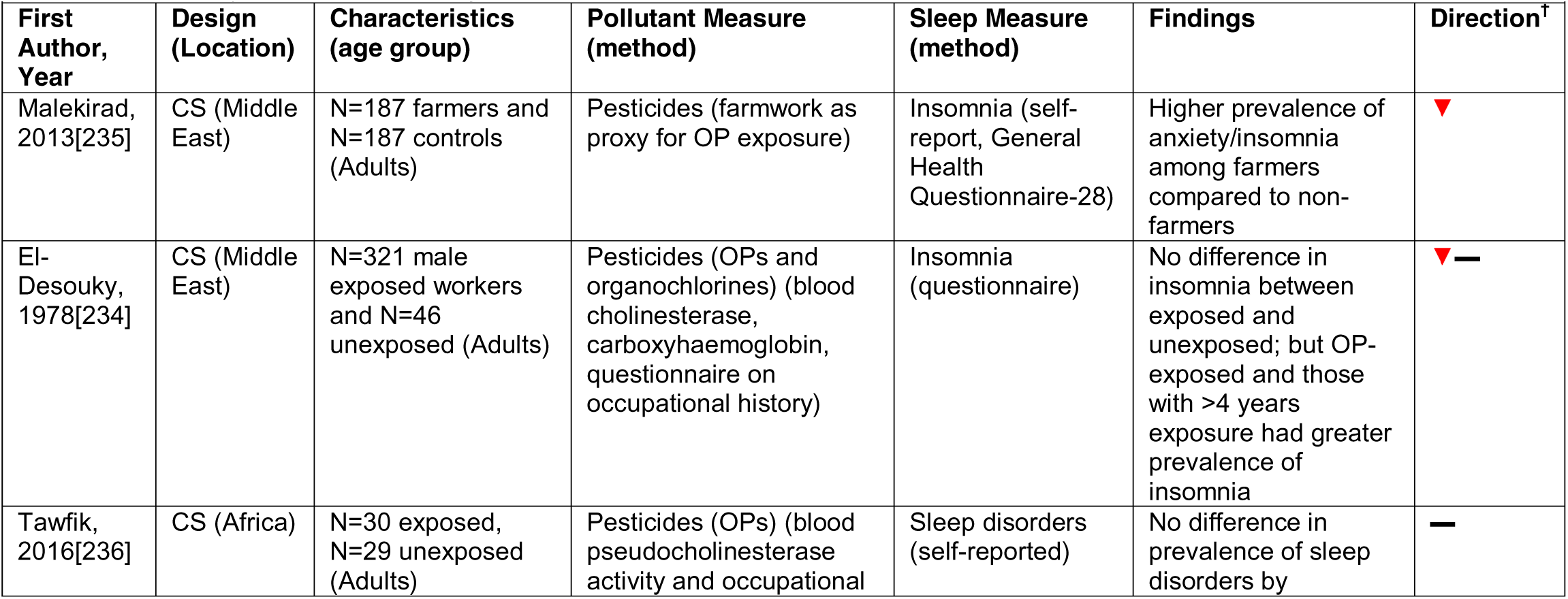
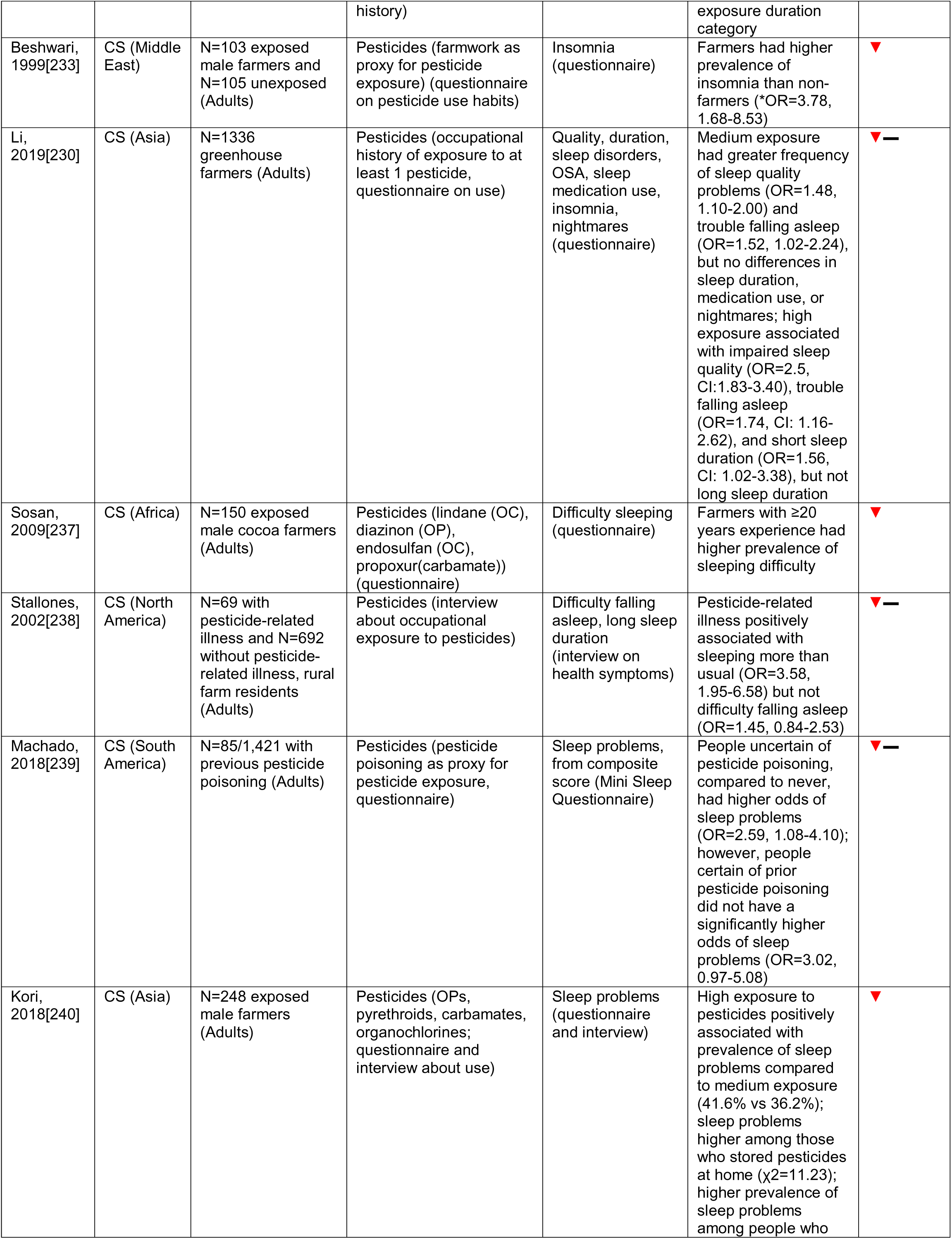
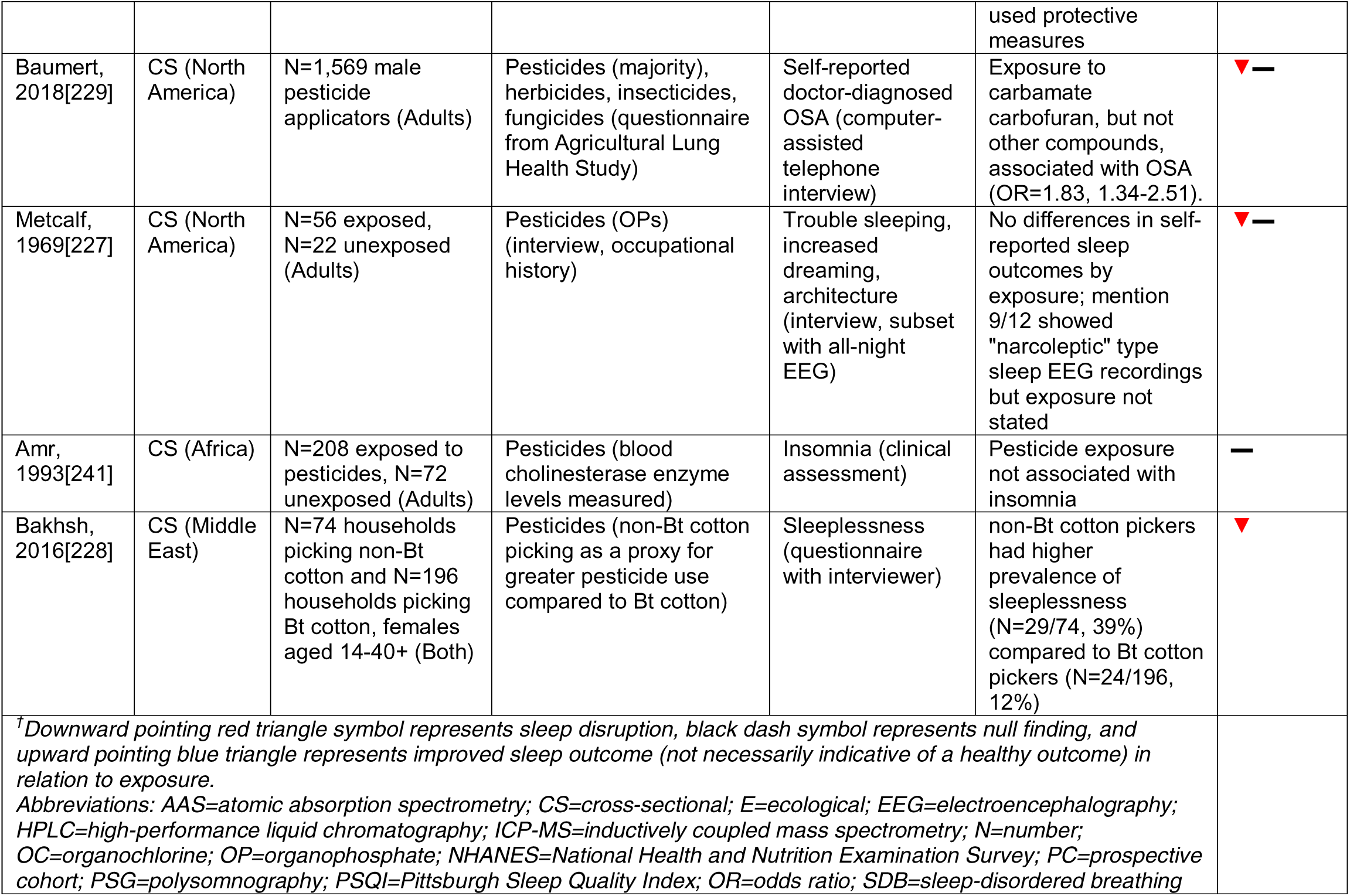
Summary table of epidemiologic studies of pesticides and sleep outcomes.

**Table 11.** Summary table of case studies / case series of pesticides and sleep outcomes.

|  |  |  |  |  |  |
| --- | --- | --- | --- | --- | --- |
| First Author, Year | Location | Characteristics (age group) | Pollutant Measure (method) | Sleep Measure (method)* | Findings |
| Bradwell, 1994[242] | Europe | N=1 male aged 21 working on farm (Adult) | Pesticides and herbicides (OPs, demeton-S-methyl [duratox], and aphid spray) (blood tests, known occupational exposure) | Duration (hypersomnia) and insomnia (self-report to clinician and family report) | Patient followed over time and clear occupational exposure; family history of mental health episodes; overall evidence is fair |
| Branch, 1986[232] | North America | N=1-3 family in household, focus on 1 male aged 75 (Adult) | Carbamate pesticide (carbaryl [Sevin]) (known regular treatment of home, blood cholinesterase levels reduced, contamination confirmed by Georgia Department of Agriculture inspector) | SDB (self-report to clinician) | All family members experienced symptoms that resolved after moving; temporality in OSA development as late clinical effect; overall evidence is good |
| Wood, 1971[243] | North America | N=1 male aged 34, duster pilot (Adult) | Pesticides (OPs, phosdrin, trichlorofon, demeton) (blood cholinesterase and known occupational history) | Sleep disturbance (self-reported), architecture (REM sleep study) | Following exposure after OP spraying and regular occupational exposure, disturbed sleep up to 17 months later, decreased REM, greater stage 2-3 sleep and 4 NREM sleep; overall evidence is good |
| Rubin, 2002[231] | North America | N=626 people from 254 homes with complete information (Both) | Pesticides (OP: methyl parathion) (urinary levels of p-nitrophenol measured with GC-MS) | Night waking (interview) | 28% of participants reported night waking, overall evidence is good |
| Tabershaw, 1966[244] | North America | N=114 people aged 14-60+ (Both) | Pesticides (OPs) (blood levels measured, interview, and/or self-report) | Insomnia (interview) | 2 cases reported insomnia, overall evidence is fair |

*\*Note: Method of self-report for sleep outcomes were not always explicitly stated in case studies, symptoms assumed reported to physician unless stated otherwise.* Abbreviations: GC-MS= gas chromatography mass spectrometry; N=number; NREM=non-rapid eye movement sleep; OP=organophosphate pesticide; OSA=obstructive sleep apnea; REM=rapid eye movement sleep; SDB=sleep-disordered breathing

### Solvents

There were 31 epidemiologic and 11 case studies of solvent exposure among the included results. Studies in this category evaluated occupational or workplace exposures to solvents, including benzene, toluene, and xylenes, among others (**Table 12**). Numerous studies investigated solvent exposures in painters. Only one study included children, but it did not specifically evaluate exposure in a pediatric population. Solvent exposure was most consistently associated with insomnia symptoms. Of the 7 studies that evaluated solvent exposure and SDB or OSA, the majority relied on PSG or oximetry for outcome measurement and supported a positive association[247,249–252]. One study by Monstad et al. repeatedly measured a subset of solvent-exposed workers with OSA, reporting overall decreases in AHI following exposure cessation and increased AHI shortly after re-exposure within participants[249]. Studies of sleep quality were mixed but suggestive of an inverse association; a study by Lundberg et al reported a worse sleep quality and greater fragmentation but no difference in other insomnia-related symptoms or OSA as exposure increased[248]. Most findings of solvent exposure and sleep duration were null, but a case study reported increased sleep requirement among 26% of 19 adult males following occupational exposure to benzene and toluene[253]. Most self-reported data was collected using tools such as the NSC-60 and the Euroquest questionnaire, which have not been validated for use in epidemiologic studies of sleep measurement. Overall, the quality of epidemiologic evidence for solvents was low (**Figure 7**), but the more rigorous studies suggested associations with sleep architecture, duration, and SDB among more highly exposed; the quality of evidence from case studies was fair (**Table 13, Supplemental Figure 1**).

**Figure 7.**
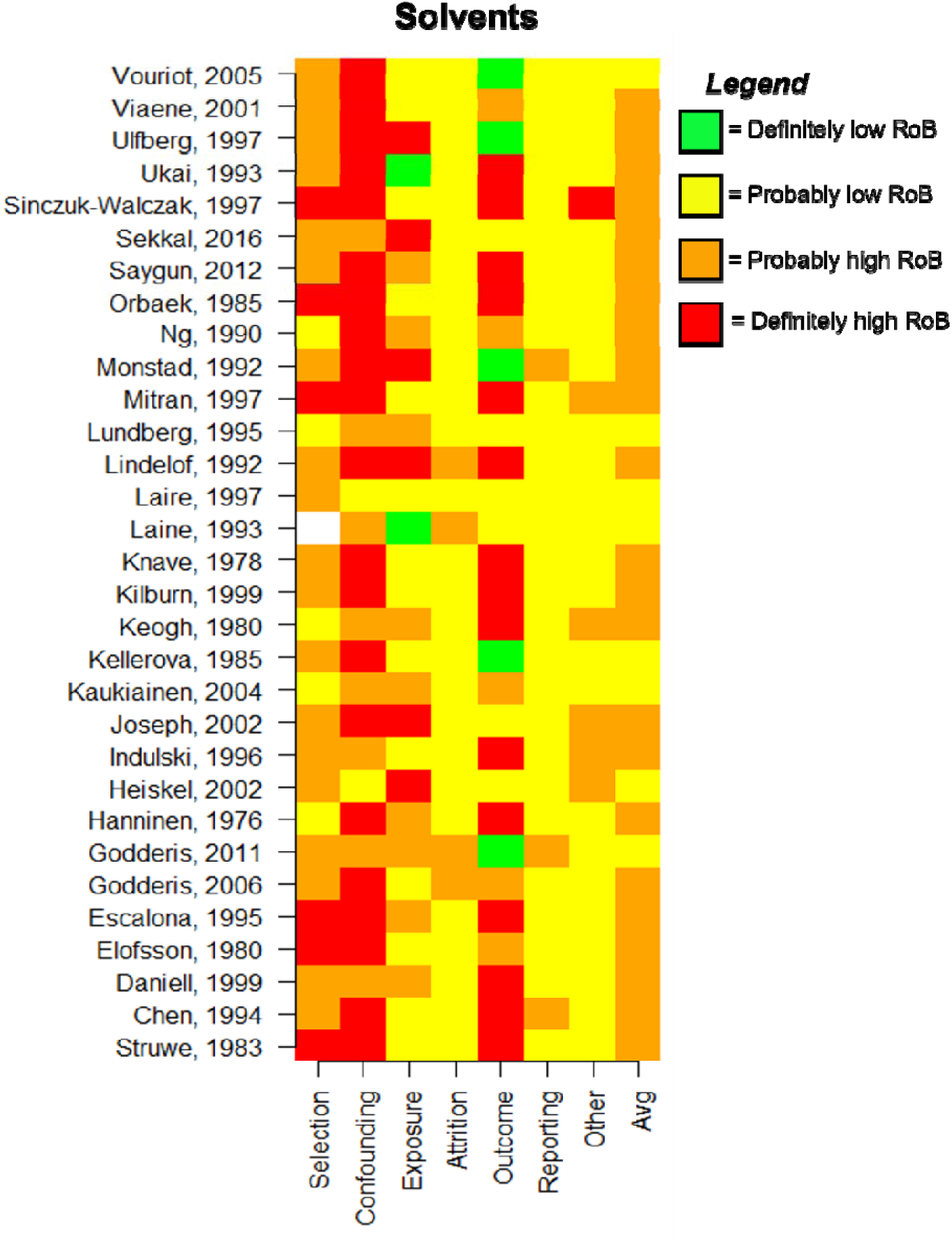
Chart depicting RoB ratings across evaluation domains for included epidemiologic studies on solvents.

**Table 12.**
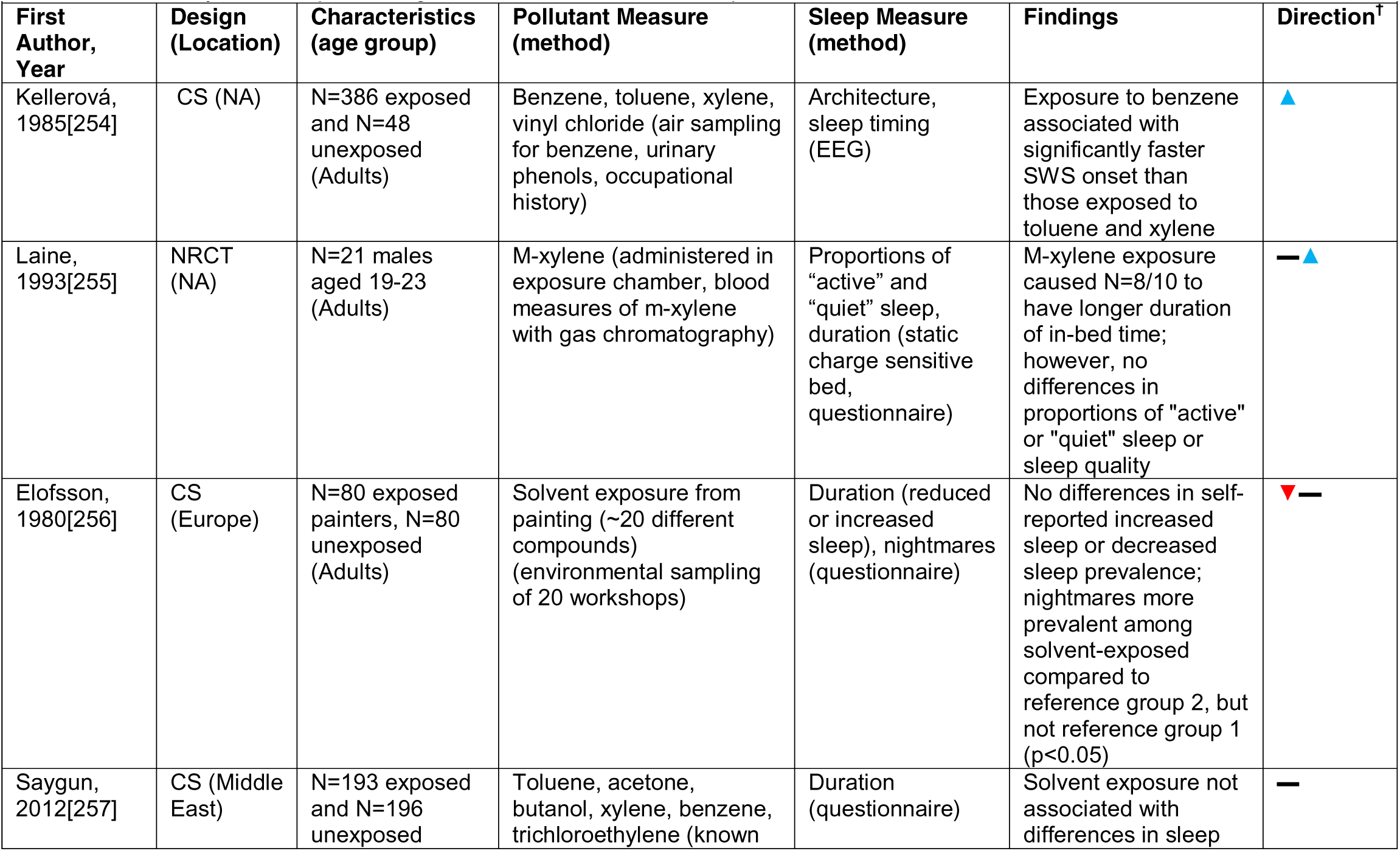
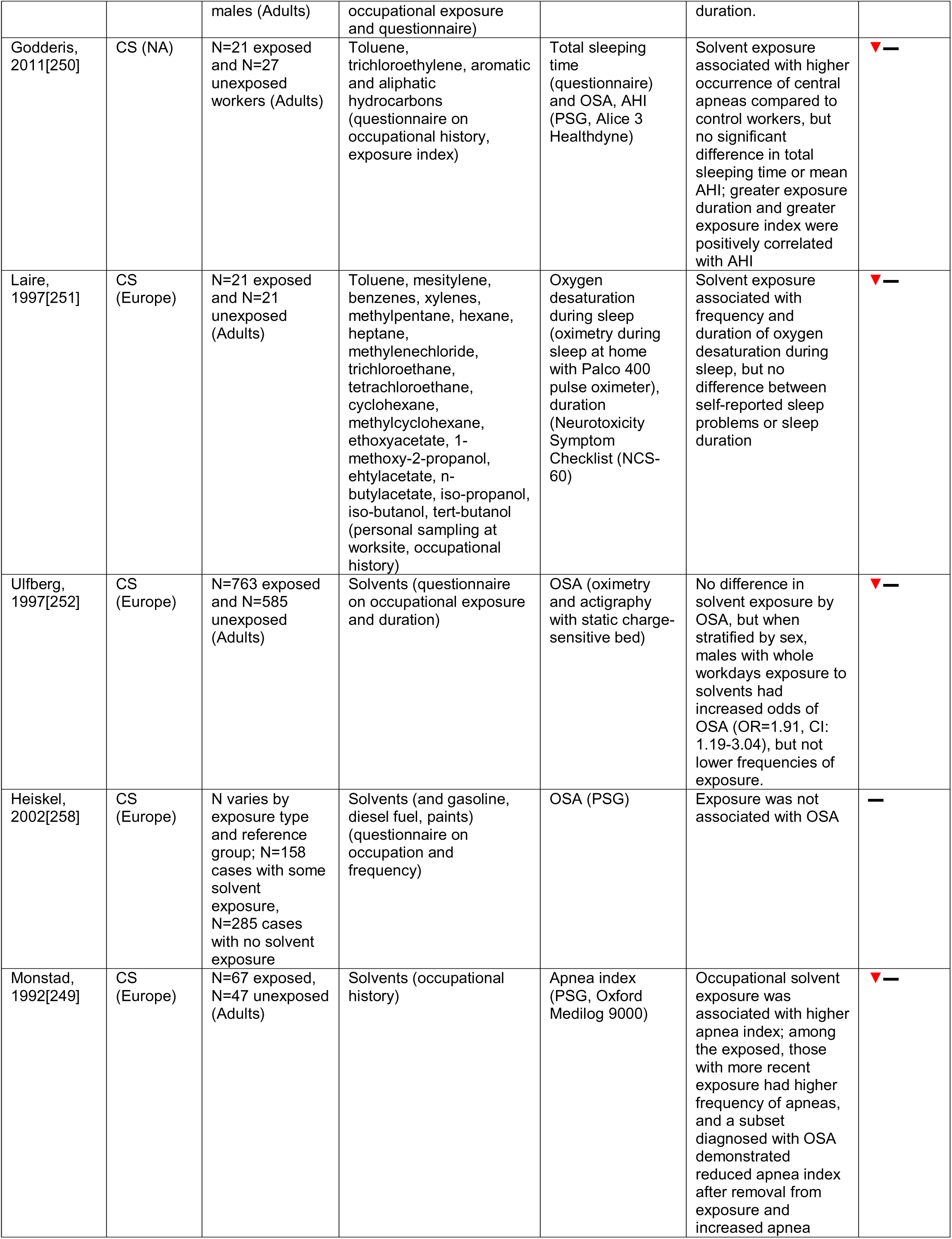
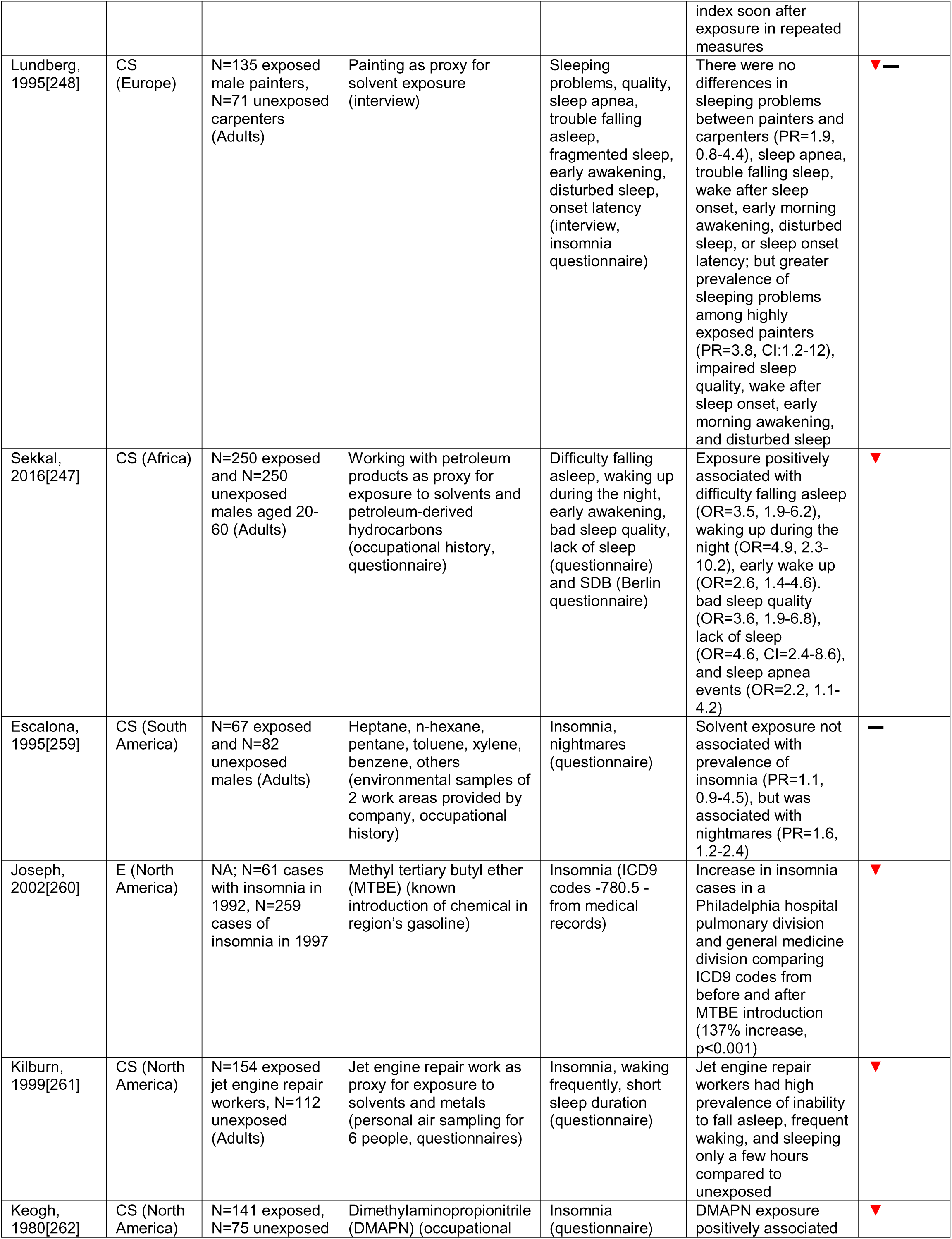
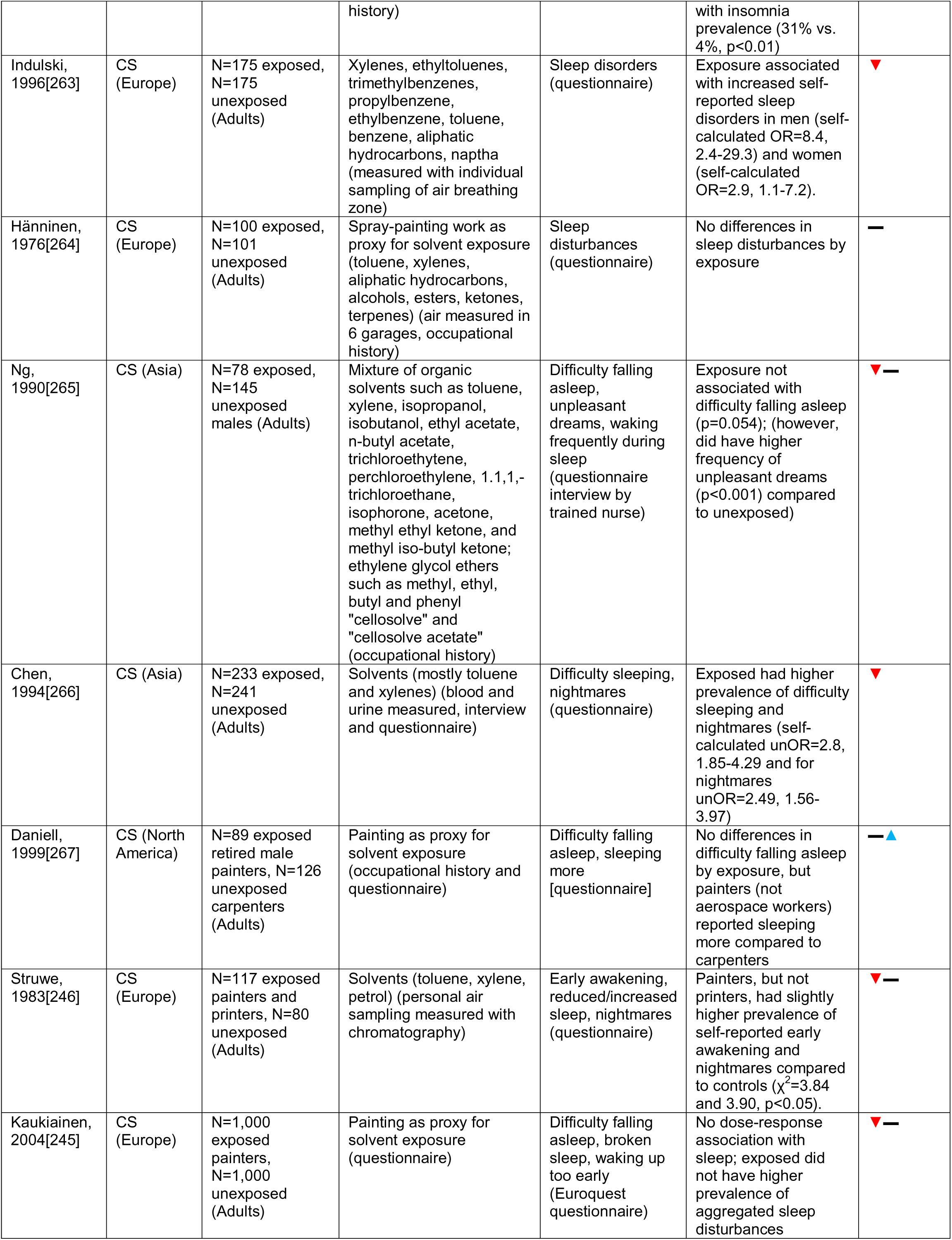
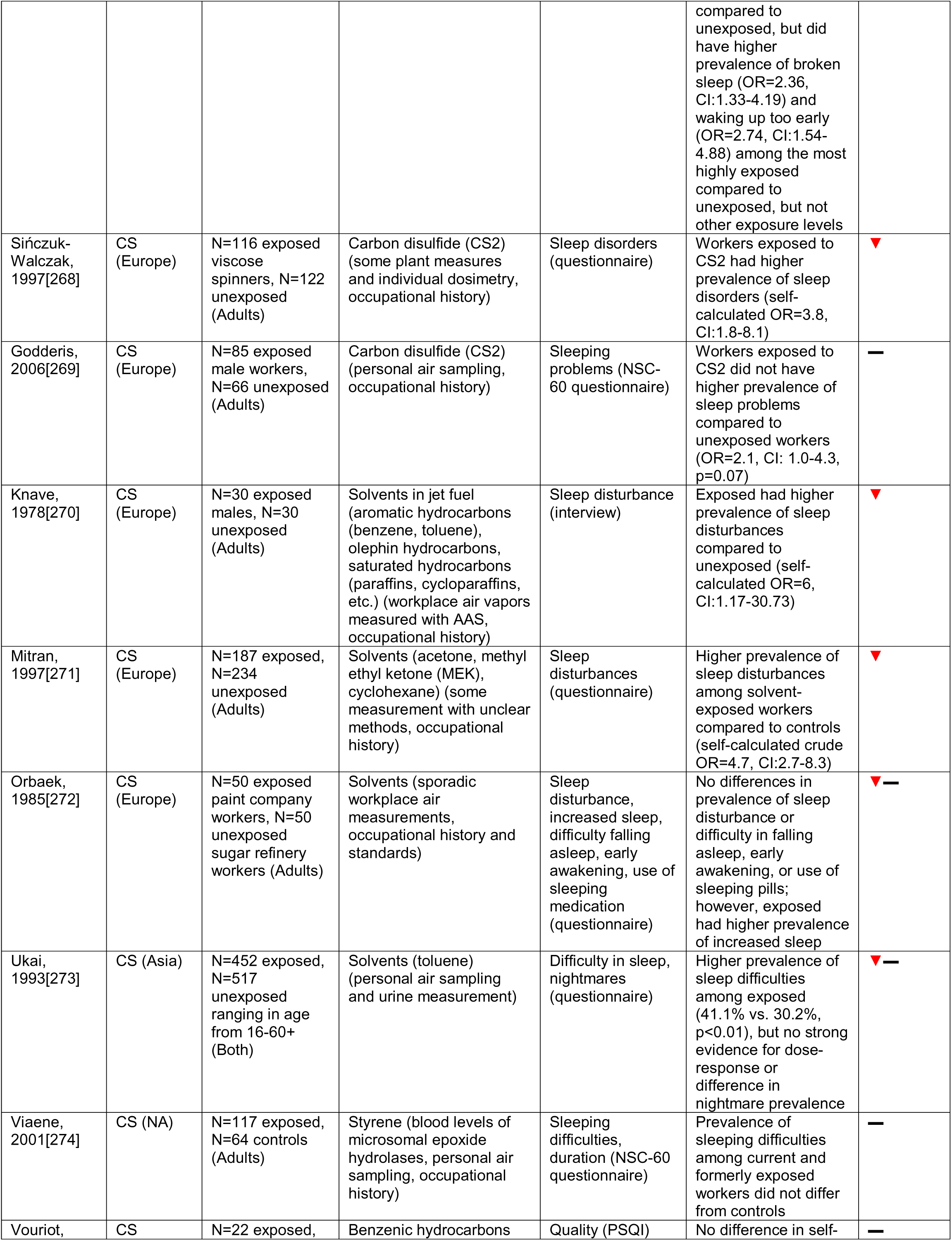
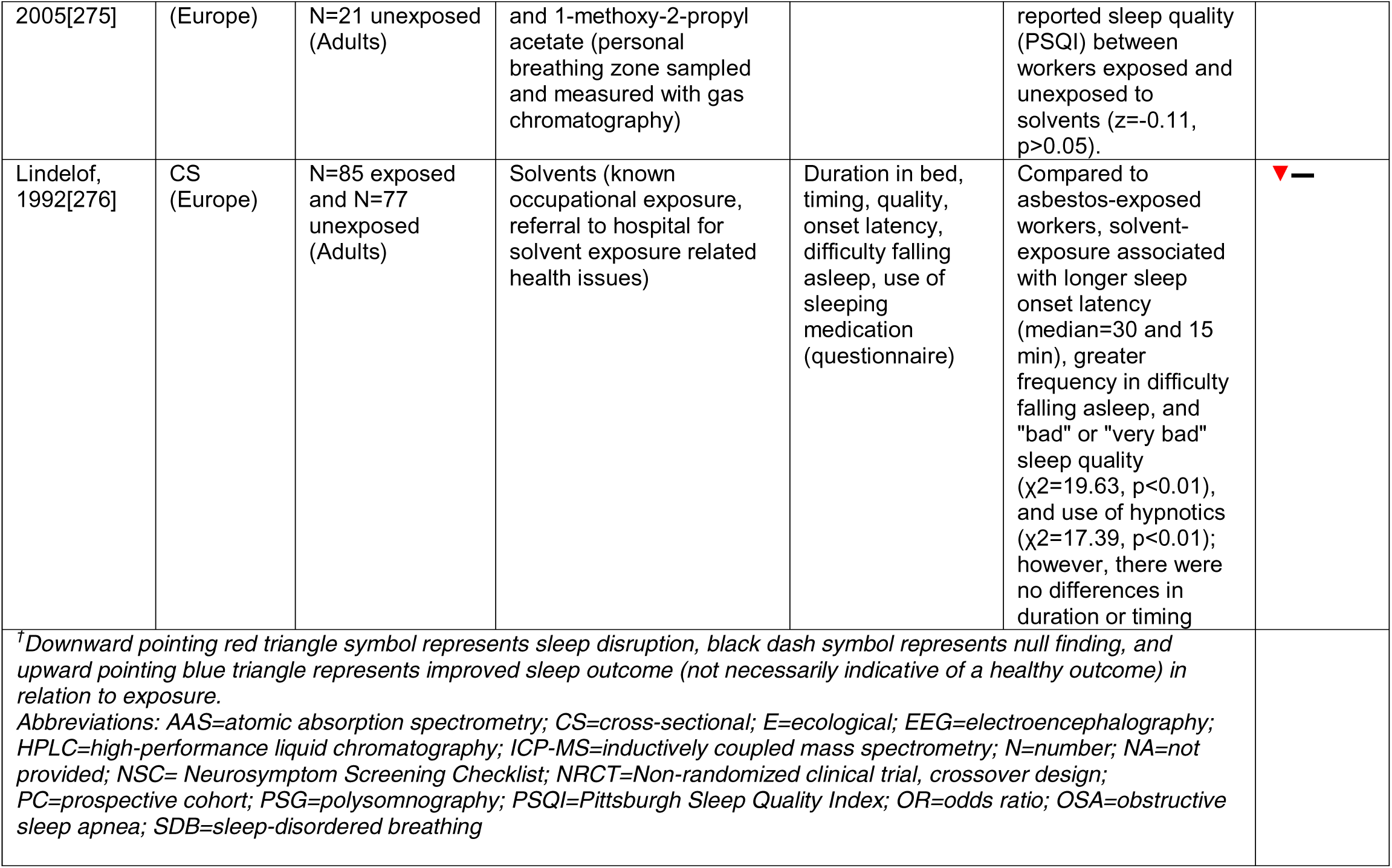
Summary table of epidemiologic studies of solvents and sleep outcomes.

**Table 13.**
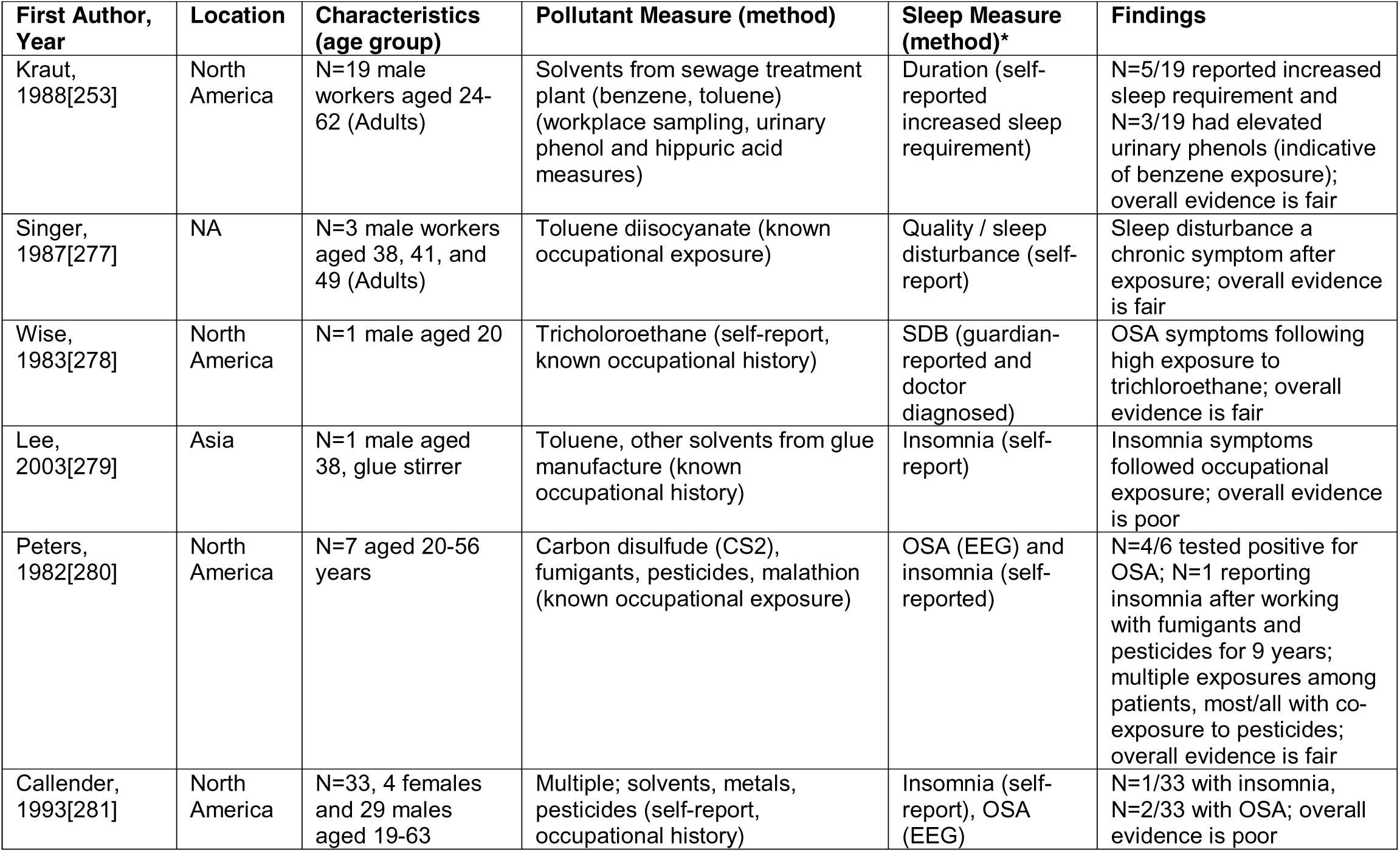
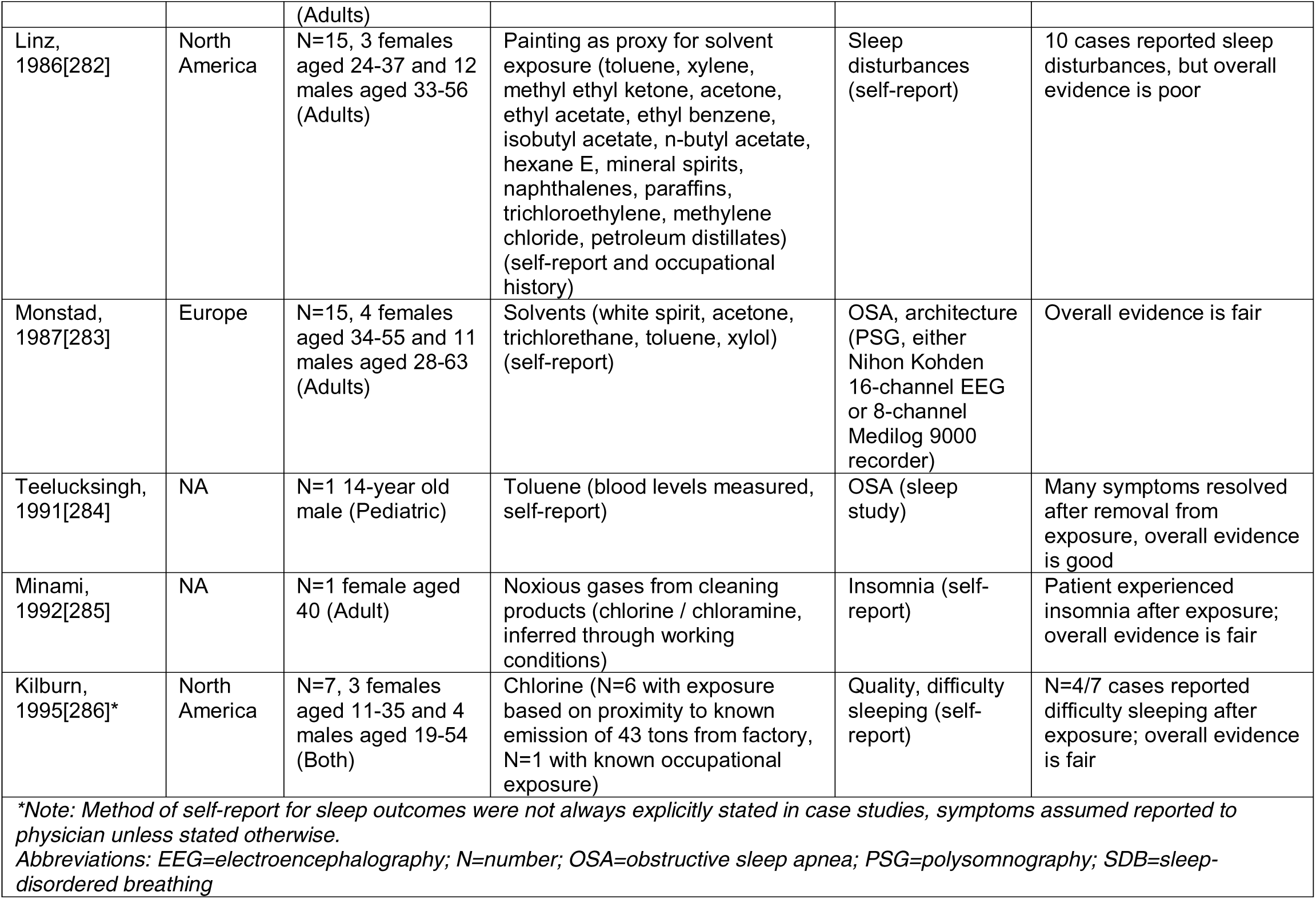
Summary table of case studies / case series of solvents and sleep outcomes.

## DISCUSSION

This review adds new information to the existing literature by systematically identifying and evaluating 204 studies of exposure to air pollution, other pollutants/endocrine disrupting compounds, metals, pesticides, solvents, and conflict-related exposures in relation to sleep health and sleep disorders and highlights possible biological mechanisms underlying these relationships. Overall, particulate matter and nitrogen dioxide were linked with poor sleep quality, and secondhand smoke in pediatric populations associated with SDB; dioxins, PCBs, and PBDEs were tied to sleep disruption and insomnia, and exposure to lead, mercury, and pesticides were also associated with insomnia. Solvents such as toluene were linked to disrupted sleep timing and SDB. Studies of conflict-related exposures relevant to GWI supported associations with poor sleep quality, insomnia, SDB, and altered sleep architecture. Pesticide exposure, particularly in pediatric populations, was not well explored. In general, the evidence presented in this review indicates environmental pollutants are detrimental to sleep health and disorders among adult and pediatric populations.

### Synthesis and Potential Mechanisms

Numerous mechanistic pathways, some shared by multiple exposures, may link environmental chemical pollutant exposure to impaired sleep health and sleep disorders. Broadly, common characteristics of exposures with consistent associations with sleep outcomes include: acting on the cholinergic system, inducing oxidative stress or inflammation, altering neurotransmission, and endocrine disruption (**Figure 8**); other possible mechanisms that weren’t explored in the literature include acting on components of the circadian clock and affecting melatonin and other relevant hormones.

**Figure 8.**
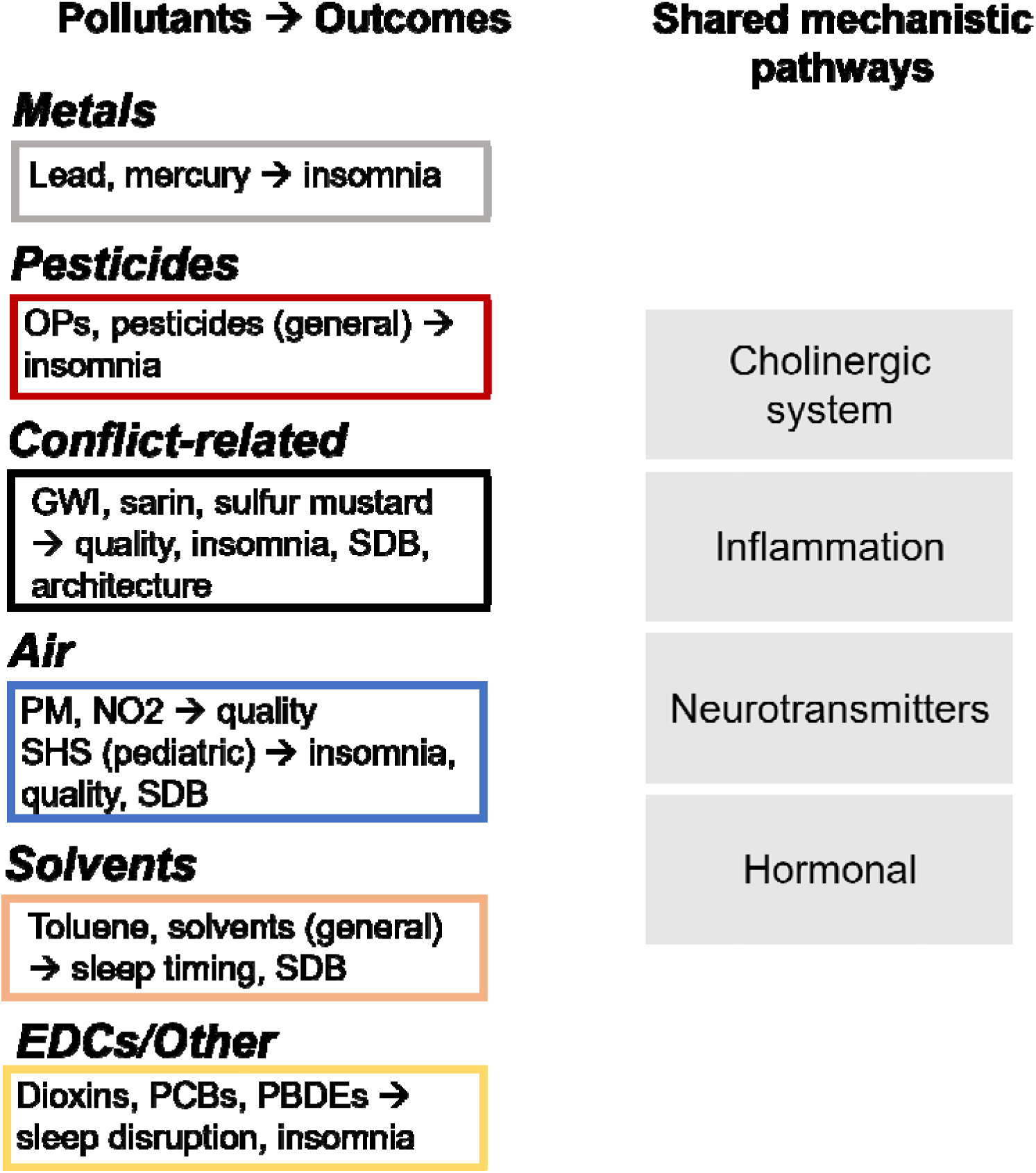
Summary of pollutant and outcome associations supported by the included literature and possible biological mechanisms underlying the relationships. (GWI=Gulf War Illness; NO_2_=nitrogen dioxide; OPs=organophosphates; PBDEs=poly-brominated diphenyl ethers; PCBs=polychlorinated biphenyls; PM=particulate matter; SDB=sleep disordered breathing; SHS=secondhand smoke)

#### Air pollution

Air pollution comprises a heterogenous mix of particulates and gases. PM and NO_2_ were most consistently associated with poor sleep quality, and, in pediatric populations, SHS with insomnia, worse quality, and SDB. PM may be composed of many different types of compounds, and is categorized by size, with PM_2.5_ the benchmark at which particles are small enough to pass directly from the lungs into the capillary blood supply and across the blood-brain barrier[54]. PM can also co-transport other chemicals, such as pesticides[287]. PM_2.5_ can induce inflammation[53] and neuroinflammatory responses[55,288–290], leading to damage and cell death. PM may disrupt sleep via systemic and/or pulmonary inflammation, but the evidence from included studies was mixed. In contrast to PM, gaseous components of air pollution may influence sleep by affecting neurotransmission. One high-quality study measured actigraphy over a 6-week period and evaluated within-person associations between air pollutants (O_3_, PM_2.5_, SO_2_, NO_2_, CO) and wake after sleep onset, sleep efficiency, sleep duration, and self-reported sleep quality, and reported an association between daily maximum ozone levels and longer sleep duration[87]. Experimental evidence suggests that O_3_ exposure increases slow wave sleep (SWS)[291–293] and alters serotonin levels[294, 295], possibly due to altered acetylcholine signaling[292]. While the other 3 included studies with O_3_ measures did not report associations, sleep outcomes were ascertained using questionnaires, ICD-10 codes, or text-mining rather than actigraphy or PSG. Findings of the included literature were similar to those presented in Liu et al, which reported overall adverse, though heterogenous, associations between ambient and indoor air pollution on sleep health[46], mostly in adult populations.

Air pollution may also influence immune system sensitization to allergens, promoting inflammation[296] and asthma incidence and exacerbation[297]. For example, NO_2_ has been linked both to pediatric asthma[298] and SDB[90], and studies previously reviewed in Liu et al[46] also supported a positive relationship between NO_2_ and greater odds of OSA[299]. Given the overlap between asthma and SDB in children[300], future studies should investigate co-exposures of air pollution and allergens as possible environmental drivers of pediatric OSA.

Furthermore, SHS exposure constitutes an additional form of environmental and household air pollution. Nicotine has well-established effects on sleep disruption, impairing initiation, maintenance, and duration[301], and affecting sleep architecture[301, 302]. However, the impact of SHS on sleep outcomes is less conclusive[51, 303]. SHS exposure is linked to higher blood inflammatory markers[304], and may be of particular concern for infants and children due to immature detoxification mechanisms and greater biological vulnerability to the effects of SHS. Heavy metals in tobacco smoke, such as cadmium, may act as co-exposures. Additionally, while vaping has grown in popularity in the last decade, no included studies specifically mentioned exposure to vaping. Therefore, the current evidence is limited, and future studies of tobacco smoke exposure should aim to capture cotinine measures in addition to device-based sleep outcomes, such as with actigraphy and PSG.

#### Conflict-related Exposures (Gulf War, WTC, Other)

Conflict-related exposures during the Persian Gulf War, the 9/11 attacks on the World Trade Center, and nerve agents were among the included studies, which were predominantly conducted among male samples. Multiple studies reported associations between exposures related to military service during the 1990-1991 Persian Gulf War and insomnia and sleep quality issues associated with Gulf War Illness (GWI). During the Gulf War, military personnel may have been exposed to OP compounds (sarin) in addition to pyridostigmine bromide (a carbamate compound), pesticides, mustard gas, air pollution from oil well fires, and uranium. Numerous reports have concluded that psychological stress and/or post-traumatic stress disorder are unlikely causes of GWI[305], suggesting an environmental etiology. Many of the symptoms of GWI overlap with those of OP and nerve agent poisoning, and while the cause and mechanism of GWI is unknown, individuals with GWI have chronic inflammation, and it is suspected that exposure to OP compounds, pyridostigmine bromide, and oil well fires may be the most likely contributors to symptoms[305]. Findings from a rat model of Gulf War exposures (permethrin, DEET, and pyridostigmine bromide over 4 weeks) supports a causal relationship, with exposure causing neurological deficits and inflammation[306]. Similar to mechanistic studies supporting a link between pesticide exposure and sleep impairment, sarin and cyclosarin are OP nerve agents that act on AChE, and experimental evidence supports a link between sarin and altered EEG patterns during sleep[307].

Four of the included studies reported associations between environmental exposures of first responders to the 2001 World Trade Center (WTC) attack with OSA. The toxic dust cloud and smoke produced by the ensuing building collapse and fire contained fine particulate matter (PM2.5), concrete, gypsum, and, at lower concentrations, asbestos, heavy metals, dioxins, pesticides, phthalates, PBDEs, and polyaromatic hydrocarbons[308, 309]. Chronic inflammation of the airways caused by this exposure could have increased the risk for development of OSA. However, the highly observed nature of these cohorts may have increased case detection.

#### EDCs/Other

EDCs such as BPA, phthalates, PCBs, PBDEs, and dioxins, can bind to nuclear hormone receptors[310], such as estrogen receptors and the aryl hydrocarbon receptor (AhR). EDCs may influence sleep via hormone signaling. For example, sleep traits exhibit sex differences[311, 312], and research in rats support a role for gonadal hormones in mediating sleep behavior in adulthood and during critical developmental windows to promote long-lasting differences in sleep behavior[313]; disruption of these processes, such as through EDCs, could potentially elicit changes in sleep behavior. For example, some phthalates, used as plasticizers, have been shown to exert estrogenic activity[314]. However, compared to an NHANES study that reported an association with shorter sleep duration[133], phthalates were not associated with sleep outcomes in premenopausal and perimenopausal participants in a prospective cohort study[137]. In contrast, dioxins, PCBs, and PBDEs were linked to insomnia and disrupted sleep. PBDEs are structurally similar to thyroid hormones[315] and could potentially influence sleep by disrupting thyroid hormone signaling[316]. Additionally, AhR activation may affect AChE and cholinergic signaling[317] as well as circadian clock function[318], additional pathways by which dioxins, PCBs, and PBDEs may be acting upon via AhR. Two studies reported positive associations between BPA levels and short sleep duration[128] and OSA[129], and additional studies reported positive associations between PCBs, PBDEs, or dioxins and sleep impairment. As lipophilic pollutants, these compounds build up in body fat over time[319] and their accumulation, or body burden, may differ by age and body fat; however, adiposity may also increase OSA risk and is associated with short sleep duration[320, 321], potentially confounding associations between lipophilic pollutants and sleep outcomes. Possible bidirectionality between sleep and fat storage may also muddy associations, as these lipophilic pollutants may promote fat development and obesity[322], but can be mobilized from fat stores to the systemic blood supply during weight loss[323]. Additionally, because dietary sources and food packaging may be main sources of phthalate and BPA body burden[324], these pollutants may be chemical proxies for diet quality and processed food consumption. Given the existing links between BMI, diet, and sleep disruption, future studies of EDCs and sleep should be specially designed to prospectively evaluate these relationships.

#### Metals

Metals were the most frequent exposure among included studies, yet no prior systematic reviews have evaluated metal pollutant exposure and sleep. Metals may impact sleep through interactions with components of the circadian clock[325], by promoting inflammation, by influencing neurotransmission, and/or by affecting the cholinergic system. There were multiple studies of copper and/or selenium (in non-occupational settings), essential trace elements that may be toxic in large amounts, with inconclusive findings; however, experimental studies in rats have shown that selenium compounds such as sodium selenite reduce sleep[326], possibly by inhibiting the enzyme that catalyzes prostaglandin D2[327], a sleep-promoting factor[328]. Lead, a toxic substance to which children are more vulnerable than adults, was most consistently associated with insomnia and quality. Lead may affect sleep by influencing neurotransmission[329], and/or by promoting inflammation as an immunotoxin[330, 331]. A few studies evaluated exposure to manganese, a micronutrient as well as an occupational hazard for welders and metal workers. During welding, manganese may become airborne, inhaled, and taken up by the olfactory or trigeminal nerves, or ingested following mucociliary clearance from the lungs[332]. Notably, high manganese exposure can cause manganism, a neurodegenerative disease with symptoms similar to Parkinson’s Disease, including sleep irregularities. In the brain, high manganese levels may affect dopaminergic[332] and cholinergic neurotransmission, such as through AChE inhibition[333].

Rodent models of chronic manganese exposure also exhibit sleep disruption[334–336]. Epidemiologic studies of occupational manganese exposure with sleep duration and insomnia were null or inconclusive, but one study of air pollution manganese exposure reported shorter sleep duration and greater use of hypnotic medication among residents in areas with higher air manganese exposure[190]. Case studies were mostly supportive of metal exposure with sleep disturbance and insomnia. Mercury was the most commonly evaluated pollutant and may cause insomnia and sleep disruption via cholinergic signaling. Mercury has high attraction for sulfhydryl groups[337] and can inhibit numerous enzymes, including cholineacetyl transferase[338–340], an enzyme responsible for synthesizing acetylcholine, and muscarinic acetylcholine receptors[341]; this may lead to increased expression of muscarinic acetylcholine receptors in the brain[342]. Experimental animal studies support a causal link between methylmercury and altered sleep-wake rhythms and sleep architecture [343, 344]. Like other metals, mercury may also affect sleep through ROS and inflammation[337]. The overall evidence between metals and sleep outcomes were mixed, but results supported a link between lead or mercury exposure and insomnia; findings were more consistent in highly exposed groups and/or case studies of overt heavy metal poisoning.

#### Pesticides

The included studies evaluated exposure to pesticides in the organophosphate (OP), pyrethroid[345], and/or carbamate class. OP compounds and carbamates inhibit AChE. AChE is an enzyme that breaks down acetylcholine, a neuromodulator and neurotransmitter[346] that plays an important role in sleep[347, 348], to prevent further neurotransmission. However, if AChE is inhibited, acetylcholine remains bound to post-synaptic receptors, leading to excessive cholinergic signaling. As the cholinergic system is an important regulator of sleep, with acetylcholine playing a role in wakefulness and REM sleep[349], acute or chronic inhibition of AChE by pesticides may affect sleep through cholinergic signaling[350]. Experimental studies in humans support a relationship between exposure to AChE agents and insomnia[351], and animal studies also support a causal relationship between pesticide exposure and sleep impairment. Interestingly, AChE inhibitor drugs are also used in treating Alzheimer’s disease symptoms; among the documented side effects of these drugs are insomnia[352] and altered sleep architecture[353, 354]. Rats exposed to chlorpyrifos, an OP, exhibited decreased sleep and sleep spindles[355] and increased sleep apnea index score[65]. OPs may also affect sleep via secondary mechanisms, such as inhibiting fatty acid amide hydrolase[356], the enzyme responsible for breaking down anandamide and oleamide[357], sleep-promoting lipids[358, 359]. In addition to inhibiting AChE, carbamates may affect sleep by binding to melatonin receptors, phase-shifting circadian rhythms, and altering pineal melatonin synthesis[360–363]; an *in silico* binding study demonstrated the ability of carbaryl and carbofuran to bind the MT1 and MT2 receptors[361]. While not evaluated in the reviewed articles, neonicotinoids, a class of pesticides that act similarly to nicotine by binding to nicotinic acetylcholine receptors, have also been shown to disrupt sleep in honeybees[364, 365]. Similar to metals, the overall epidemiologic evidence for pesticides were mixed, whereas case study findings tended to support an association. This may be due to reporting bias, in that case studies may be more likely to report the presence, rather than absence, of sleeping problems. Additionally, case studies tended to evaluate acute, high exposure or outright poisoning, whereas epidemiologic studies relied on farm work as proxy or self-reported data on prior pesticide poisoning symptoms, for example. Despite possible mechanistic pathways, the overall epidemiologic evidence for pesticides and sleep disruption was weaker than the evidence for air pollution or metals due to lack of device-based exposure and/or outcome measurements.

#### Solvents

Solvents, such as toluene, benzene, and xylenes, are well-established occupational hazards; chronic or acute exposure can lead to solvent-induced encephalopathy. Solvents are mucosal irritants which can cross the blood-brain barrier and act as central depressants[366], inhibiting glutamatergic transmission and promoting GABAergic and glycinergic signaling[367]. Experimental evidence suggests toluene may stimulate dopamine release[368], alter GABA_A_ receptor function[369], and/or inhibit nicotinic acetylcholine signaling[370]. Animal studies have also shown that toluene exposure alters monoamines and SWS in rats [371–375]. Solvents may act through mechanisms similar to that of anesthetics[367], which have been shown to decrease muscle tone and increase upper airway collapsibility[376], a primary risk factor for OSA. Therefore, it is possible that solvents may influence OSA development by promoting inflammation and airway collapse. However, the overall evidence between SDB/OSA and solvent exposure was low, with most studies lacking device-based exposure and/or outcome measures with small sample sizes and/or unadjusted for potential confounders. Therefore, the association between occupational solvent exposure and OSA is unclear within the realm of current workplace standards, and large prospective cohort studies of solvent exposure should consider including validated sleep and SDB measures as health outcomes.

### Pediatric Considerations

Most of the included studies were conducted among adult populations, with few comprehensive assessments of pediatric sleep health. Compared to the adult literature, there were no pediatric studies with environmental exposure to conflict-related exposures and few studies of pesticides; there was one pediatric case study of solvent exposure. Additionally, 86% of pediatric studies included device-based measures of exposure, whereas only 19% included device-based sleep outcome measures.

Among pediatric populations, the most common reported exposures to metals were lead (n=8) and mercury (n=7). A cross-sectional study reported that blood lead levels ≥10 μg/dL were associated with decreased sleep duration in Mexican children aged 6–8 years[156]. Follow-up longitudinal studies in China[153] and Mexico City[155] similarly reported that elevated blood lead levels in early childhood were associated with an increased risk of insomnia and excessive daytime sleepiness[153] and decreased sleep duration[155] in later childhood. Moreover, in a prospective birth cohort, lead levels were measured in maternal blood in early and late pregnancies, in cord blood at birth, and in 2, 3, and 5-year-old children’s blood.

Interestingly, postnatal blood lead levels at 2 years old were associated with sleep problems in females only, not in males. The authors suggested that differing sexual brain development between sexes may partially explain these findings. A cross-sectional study of children ages 9 to 11 reported that higher blood mercury levels were associated with shorter sleep duration[163]. However, a more recent cross-sectional study of Mexican children did not support an association between urinary mercury and sleep duration[164], although toxicokinetics and measurement in different biomatrices may explain discordant findings. Overall, there is good evidence for negative consequences of early lead exposure and poorer sleep outcomes.

There’s mixed evidence for SHS and sleep symptoms, however, this mostly depended on the study population being evaluated. In healthy children, self-reported SHS exposure was associated with increased Sleep-Related Breathing Disorder (SRBD) scores in school-aged children in Chile[105] and Turkey[91]. In Japanese adolescents, self-reported SHS exposure was associated with self-reported symptoms of difficulty with initiating sleep, maintaining sleep, early morning awakening, insufficient sleep, and short sleep duration[108]. Additionally, in a cohort of healthy children ages 2-5 years with SHS measured by hair nicotine, there was an association with self-reported sleep-related breathing problems. In children with asthma from Cincinnati ages 5-13 years, serum cotinine levels were associated with elevated SRBD scores and self-reported sleep problems including longer sleep onset delay, parasomnias, daytime sleepiness, and overall sleep disturbance[108]. However, in children ages 7-11 years with overweight or obesity, there was no association between SRBD scores and passive smoke exposure when measured by plasma cotinine assay[104]. Also, in a small cohort of 31 neonates referred for PSG with suspected gastroesophageal reflux disease (GERD), there were no differences in sleep outcomes between exposed vs unexposed (as measured by urine cotinine assay), but there was a 2-fold greater number of reflux events in infants with exposure to passive smoke[98]. Overall, there’s good evidence that SHS exposure is associated with poor sleep outcomes in healthy children and with asthma, however, a limitation of the evidence is that all the studies have a cross-sectional design; therefore, a causal association for adverse sleep outcomes could not be ascertained.

Only one pediatric study evaluated the association between air pollution and device-based measures of SDB. SDB was characterized by AHI >1 event per hour (often used as a pediatric cutoff for mild SDB) among children admitted to a hospital in Italy who had a sleep study to evaluate for SDB due to sleep-related breathing symptoms. The authors then used geographical monitoring data to determine the mean NO_2_ levels and reported a positive correlation between mean AHI and NO_2_ levels, with strongest association among children living in an area characterized by a high density of traffic-related pollutants[90]. However, in addition to the ecological study design, a limitation of this study is the low AHI threshold used to define the outcome; the use of a higher AHI threshold may have provided more clinically meaningful information. Overall, there is weak evidence that elevated air pollution influences the prevalence of SDB in children.

### Limitations of this Review

The findings presented in this review are not without limitations. Due to the heterogeneity of pollutants and how sleep outcomes were measured, we did not perform a meta-analysis of the results or compare effect estimates. In some cases, multiple exposures or sleep outcomes were grouped together as a single item, limiting our ability to tease apart specific health associations and making interpretation difficult; for example, a study with an outcome of “sleep disorders” which comprised ICD-10 codes for insomnias, hypersomnias, sleep-wake disorders, cataplexy/narcolepsy, and sleep apnea[92]. Additionally, while we present results on SHS/ETS and conflict-related exposures, the search strategy was not designed with these exposures in mind; in particular, the findings presented on SHS/ETS are not a comprehensive presentation of the literature, and prior systematic reviews on this topic should be sought for further coverage on the topic[50, 51].

### Current Gaps and Future Directions

Sleep is multi-dimensional, and due to the heterogeneity of how sleep health was evaluated, comparison across studies was limited. The majority of studies relied on questionnaire data, although such measures of sleep are prone to measurement error and reporting bias. For example, self-reported sleep duration is an overestimate of actigraphy-measured sleep[377], and the amount of overestimation may differ by race and ethnicity[378]. Many studies relied on one or limited measures of sleep health, thus preventing the evaluation of multiple dimensions of sleep health. Questionnaires such as the PSQI or STOP-BANG have been evaluated for use in studies of sleep, but a significant portion of included studies did not employ validated instruments and/or relied upon a single sleep-related question as an outcome. Additionally, insomnia is a self-reported condition that is diagnosed clinically by interview and patient response, but most studies of questionnaire-assessed insomnia did not consider chronicity or associated daytime impairment – features needed to diagnose insomnia as a clinical disorder [10]. In summary, self-reported sleep outcomes may be capturing different elements of sleep than device-based measures that are nevertheless important for health, and future studies with self-reported sleep measures should aim to assess sleep in a comprehensive way.

Among device-based measures, PSG is considered the gold standard for sleep measurement and captures sleep architecture in addition to OSA diagnosis and sleep continuity. PSG generally occurs overnight in a sleep laboratory, which can be expensive, time-consuming, and affected by the “first night effect” if measured over a single night. However, home-based PSG systems or PSG-like systems measure participants in their home environment and may represent a viable study design option for community-based studies. Actigraphy, which is based on detecting rest and activity patterns, can also be employed to derive measures of sleep-wake behavior and may be less onerous, more cost-effective, and allow for longer measurement than PSG. While the myriad specifications of these devices should be thoroughly considered in study design[379], in-home PSG and actigraphy both have the potential to advance our understanding of environmental contributors to sleep health, particularly among low-income populations that may face barriers to undergoing overnight in-lab sleep studies.

Another gap in the current research is investigation of the extent to which sleeping and circadian phase influence pollutant exposure and vulnerability. Vulnerability to toxicants may differ by time of the day[380] due to rhythmic fluctuation in detoxification systems; certain exposures may be more harmful at night, for example. Rhythmicity in excretion or biomarkers, such as daily rhythms in urine production, further complicates these relationships when measuring exposure[381]. Unfortunately, most studies did not provide details on the timing of sample collection or measurement or take circadian phase into account. Surprisingly, only one study included a measure of melatonin[124], and few studies investigated sleep timing. Furthermore, people spend approximately a third of their lifetimes sleeping, but the effect of indoor sleep environments on pollutant exposure has received limited attention. For example, numerous studies have examined outdoor air pollution and sleep, but fewer have focused on HAP or indoor air pollution with device-based measures of exposure[89, 97]. In summary, future research would benefit from integrating measures of circadian phase in the exposure/outcome measurement or as an interaction in the exposure-outcome relationship and timing of measurement and sample collection in study design. Shift work or other exposures which may disrupt circadian rhythms should also be considered. Additionally, future studies should evaluate the contribution of the sleep environment to pollutant exposure.

Statistical modeling is another important aspect of current and future research. Studies of sleep quality and OSA commonly dichotomize outcomes based on thresholds for the PSQI or AHI, for example, which may bias results depending on the placement of the cutoff value. Additionally, few studies have considered bidirectionality in exposures and outcomes, but people with impaired sleep and/or circadian misalignment may be more vulnerable to the effects of pollution or have altered detoxification/metabolic pathways that influence biological exposure measurement[382–384]. For example, there may be a bidirectional relationship between sleep and oxidative stress, where sleep helps protect against negative consequences of reactive oxygen species (ROS) and impaired sleep increases vulnerability to ROS[385], although further research in humans is necessary. Co-exposures were rarely considered in the included literature, which is another limitation of the current research; for example, areas with air pollution due to vehicle emissions may consequently also have higher burden of light at night and noise pollution. Future studies would benefit from considering continuous outcomes, bidirectionality, and co-exposures in study design and model building.

Environmental pollutant exposures tend to be patterned by socioeconomic status and race/ethnicity. Due to systematic racism and redlining policies that resulted in segregated, under-resourced neighborhoods, marginalized communities in the U.S. are more likely to be exposed to environmental chemical pollutants and adverse characteristics of the built environment[71], such as older housing and lead exposure[68] and pesticide use[386]. Data demonstrates that racial minorities are more likely to have exposure to air pollution[69]; yet few of the U.S.-based studies reviewed were conducted among these populations. Racial/ethnic minorities and lower socioeconomic individuals have a high prevalence of adverse sleep health and OSA[387, 388], yet few studies examine environmental pollutants as contributors to sleep disparities. Future studies in the U.S. and globally should aim to expand study samples to enroll sufficient sample sizes of minoritized individuals, in addition to including measures of environmental pollutant exposure in sleep health disparities research studies.

This review of current evidence of environmental exposures and sleep outcomes in adults and children reveals the need for future research to consider a life course approach. Few of the included pediatric sleep studies considered school start times, bedtime routines, and puberty-related hormonal shifts in chronotype in the study design and analysis. Additionally, children may be more vulnerable to pollutants’ effects on brain development and sleep health; harmful environmental exposures potentially differ by age, and pediatric populations are of special interest because infants and children have no or underdeveloped detoxification systems. While the adult literature between pesticide use and sleep outcomes was also somewhat sparse, the lack of studies investigating pediatric pesticide exposure and sleep outcomes should be considered as a gap in knowledge, given the known detrimental effects of pesticides on children’s health[389]. Future directions should include evaluating pediatric exposures to pesticides, metals, EDCs, and air pollution in relation to sleep health.

## Conclusions

There is documented evidence that air pollutants, conflict-related exposures, EDCs, metals, pesticides, and solvents are classes of environmental pollutants associated with sleep health. Possible biological pathways underlying pollutant-sleep relationships included cholinergic signaling, inflammation, neurotransmission, and hormonal signaling. Main limitations of the current research include lack of device-based sleep measurement, lack of prospective investigation, and little to no mechanistic evaluation. Based on the limitations, future studies should aim to include measures of circadian phase and timing of sample collection, evaluate bidirectionality and co-exposures, capture device-based measures of both exposure and sleep outcomes, include pediatric populations and historically minoritized individuals, and consider a life course approach in study design and interpretation to develop a more intersectional and holistic understanding of how our environment influences sleep health. To improve collaboration and scientific discovery, future and existing studies should strive to make their data accessible and well-annotated; researchers can contribute to the sleep research community by sharing data in repositories such as the National Sleep Research Resource (NSRR, https://sleepdata.org/), supported by the National Heart, Lung, and Blood Institute of the National Institutes of Health. There is a large gap in current knowledge around environmental pollutant exposure and pediatric sleep health, especially regarding pesticides. Overall, future studies should be robustly designed to evaluate environmental exposures and sleep health, with device-based measures of exposures and outcomes. Longitudinal study design incorporating measures of actigraphy and wearable devices, including representation of diverse backgrounds, as well as clear reporting and data availability will advance our understanding of the environmental contributions to sleep health across the life course.

## Supporting information

Supplemental File 1_Search terms

Supplemental File 2_Reviews

Supplemental File 3_Data extraction questions

Supplemental File 4_Risk of bias questions

Supplemental File 5_Epidemiological study consensus

Supplemental File 6_Case study consensus

## Data Availability

Search terms and results of the systematic review are provided in the main tables and figures as well as the Supplemental Files.

## ACKNOWLEDGEMENTS

Supported by funding from the National Institutes of Health (NICHD F31-HD097918 and NIH-NHLBI T32HL007901 [to DW and SGN], R35 HL135818 [to SR], and K01HL138211 [to DJ]). DW proposed and designed the study, performed screening and data extraction, created summary tables and figures, and wrote and edited the manuscript. JG performed screening and data extraction. SP developed the search strategy, performed the literature search, wrote the search methods, created the PRISMA chart, and edited the manuscript. SG-N contributed to writing and synthesis of the pediatric literature. KF contributed to search strategy and methods. SR contributed to results interpretation, synthesis, and edited the manuscript. All authors reviewed and edited the manuscript. DAJ contributed to study design, classification of exposures and outcomes, results interpretation, synthesis, and edited the manuscript.

## Data Availability

The search terms and extracted data used for plotting the RoB figures is provided in Supplemental Files.

---

**Supplemental Figure 1.**
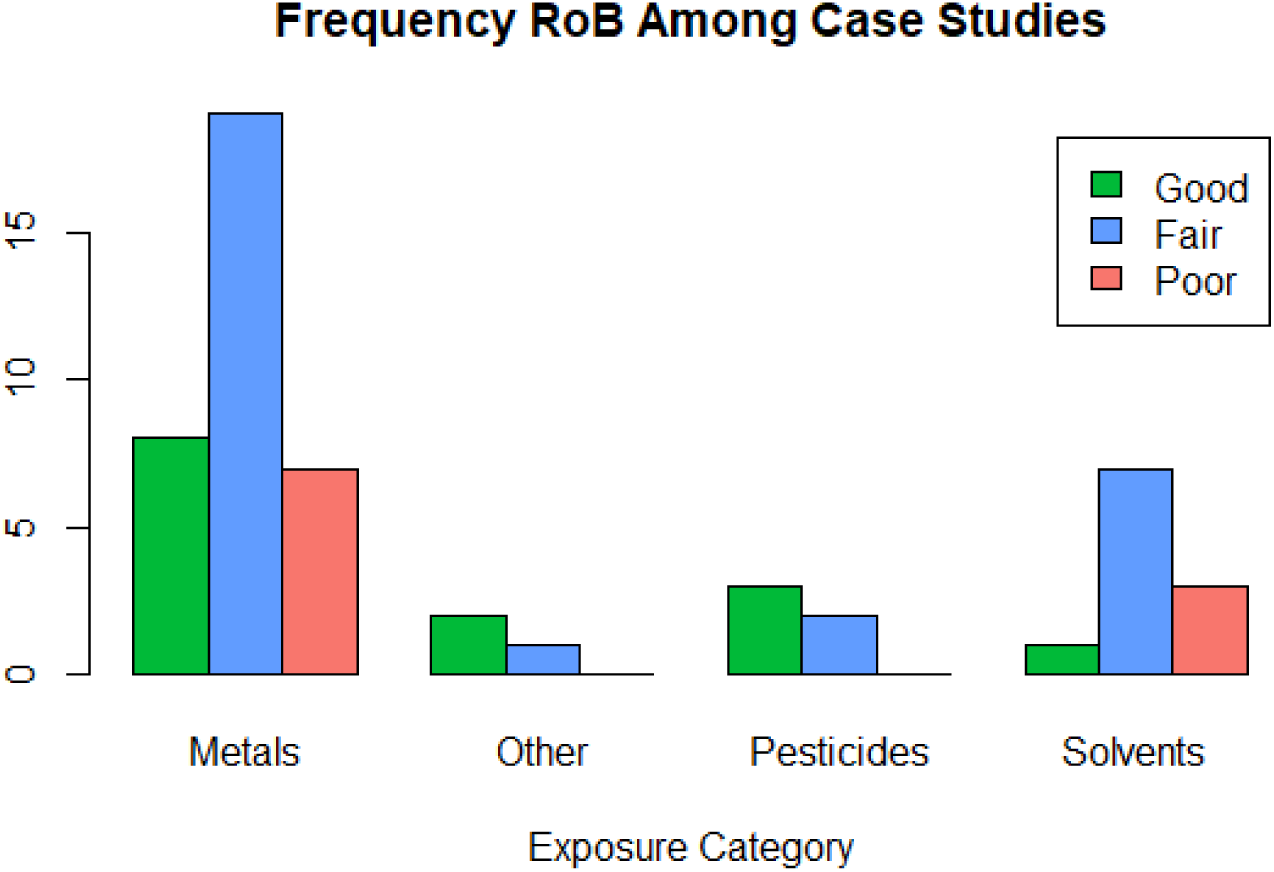
Plot of Risk of Bias determination for case studies by exposure pollutant group.

