## Supplemental File 1_Search terms for "Is exposure to chemical pollutants associated with sleep outcomes? A systematic review"

**Supplemental File 1.** Search terms and search details for the 9,198 articles identified through database searches.

**PubMed = 2595**

**April 24, 2021**

| **#1** | (pollut*[tiab] OR "environmental exposure"[tiab] OR hydrocarbon*[tiab] OR pesticide*[tiab] OR insecticide*[tiab] OR Organophosphate*[tiab] OR Naphthalene*[tiab] OR "heavy metal"[tiab] OR "heavy metals"[tiab] OR mercury[tiab] OR arsenic[tiab] OR cadmium[tiab] OR "lead poisoning"[tiab] OR selenium[tiab] OR manganese[tiab] OR antimony[tiab] OR chromium[tiab] OR copper[tiab] OR zinc[tiab] OR aluminum[tiab] OR benzene[tiab] OR Bisphenol[tiab] OR BPA[tiab] OR "polyvinyl chloride"[tiab] OR Phthalate*[tiab] OR "environmental pollutants"[MeSH Terms] OR "environmental exposure"[MeSH Terms] OR "Pesticides"[Mesh] OR "Pesticides"[Pharmacological Action] OR "Organophosphates"[Mesh] OR "Metals, Heavy"[Mesh] OR "Transition Elements"[Mesh] OR "Heavy Metal Poisoning"[Mesh]) |
| --- | --- |
| **#2** | (sleep*[tiab] OR Insomnia[tiab] OR Polysomnograph*[tiab] OR Actigraph*[tiab] OR "Restless leg syndrome"[tiab] OR hypersomnia[tiab] OR "Sleep Wake Disorders"[Mesh] OR "Actigraphy"[Mesh]) |
| **#3** | #1 AND #2 |
| **#4** | #3 AND English[lang] AND ("Journal Article" [Publication Type] OR "Review" [Publication Type] OR "Systematic Review" [Publication Type]) NOT ("animals"[MeSH Terms] NOT "humans"[MeSH Terms]) |

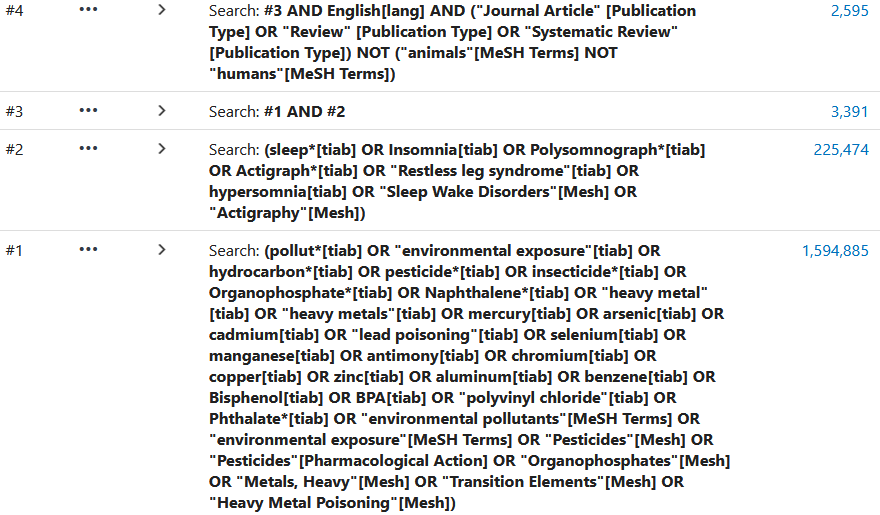

**Embase (Elsevier) = 1298**

**4/24/2021**

| #1 | (pollut*:ti,ab OR 'environmental exposure':ti,ab OR hydrocarbon*:ti,ab OR pesticide*:ti,ab OR insecticide*:ti,ab OR organophosphate*:ti,ab OR naphthalene*:ti,ab OR 'heavy metal':ti,ab OR 'heavy metals':ti,ab OR mercury:ti,ab OR arsenic:ti,ab OR cadmium:ti,ab OR 'lead poisoning':ti,ab OR selenium:ti,ab OR manganese:ti,ab OR antimony:ti,ab OR chromium:ti,ab OR copper:ti,ab OR zinc:ti,ab OR aluminum:ti,ab OR benzene:ti,ab OR bisphenol:ti,ab OR bpa:ti,ab OR 'polyvinyl chloride':ti,ab OR phthalate*:ti,ab) |
| --- | --- |
| #2 | (sleep*:ti,ab OR insomnia:ti,ab OR polysomnograph*:ti,ab OR actigraph*:ti,ab OR 'restless leg syndrome':ti,ab OR hypersomnia:ti,ab OR 'sleep'/mj OR 'sleep disorder'/mj OR 'actigraph'/exp OR 'restless legs syndrome'/exp OR 'sleep parameters'/exp OR 'sleep debt'/exp OR 'sleep induction'/exp OR 'sleep pattern'/exp OR 'wake after sleep onset'/exp) |
| #3 | #1 AND #2 |
| **#4** | #3 AND [english]/lim AND ([article]/lim OR [article in press]/lim OR [review]/lim) NOT ('animals'/exp NOT 'humans'/exp) |

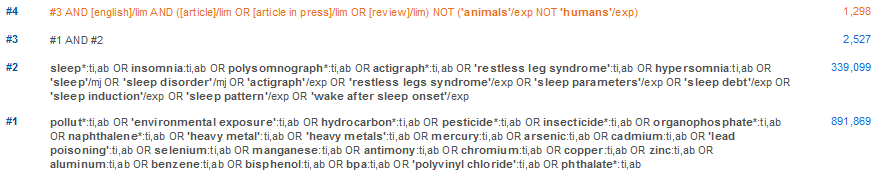

**Environment Complete (EBSCO) = 282**

**5/15/2021**

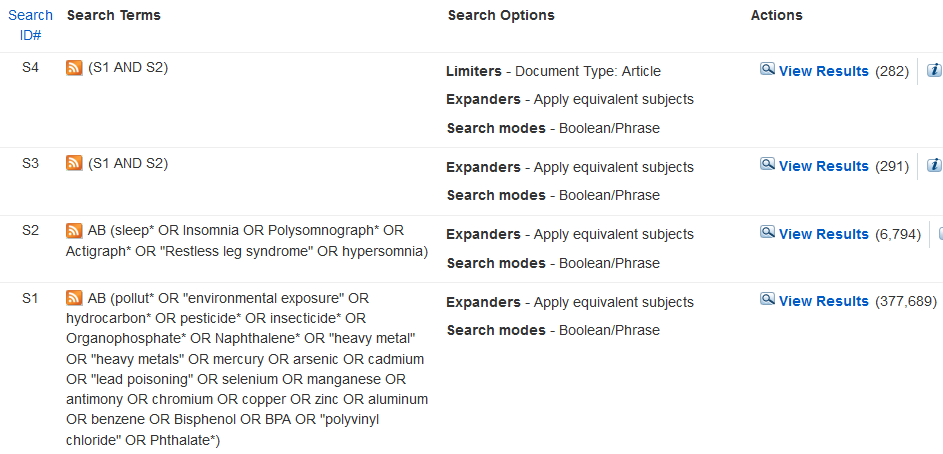

**Web of Science Core Collection = 2217**

[*Science Citation Index Expanded, Social Sciences Citation Index, Arts & Humanities Citation Index, Emerging Sources Citation Index, Conference Proceedings Citation Index, Book Citation Index, Current Chemical Reactions, Index Chemicus*]

**5/15/2021**

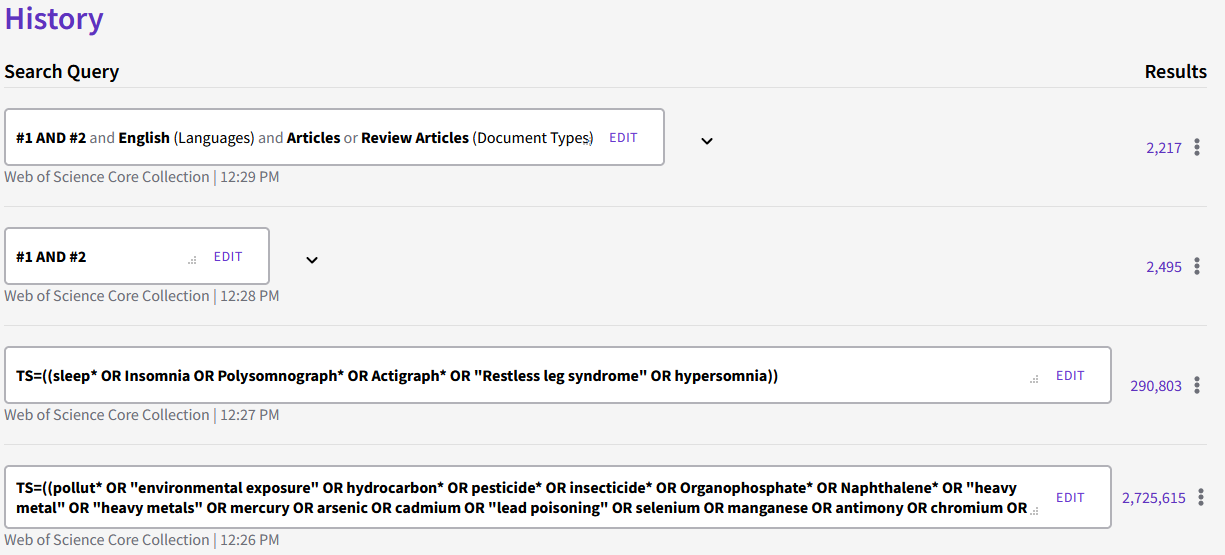

**Scopus (Elsevier) = 1771**

**5/15/2021**

| **#1** | ABS ( ( pollut* OR {environmental exposure} OR hydrocarbon* OR pesticide* OR insecticide* OR organophosphate* OR naphthalene* OR {heavy metal} OR {heavy metals} OR mercury OR arsenic OR cadmium OR {lead poisoning} OR selenium OR manganese OR antimony OR chromium OR copper OR zinc OR aluminum OR benzene OR bisphenol OR bpa OR {polyvinyl chloride} OR phthalate* ) ) |
| --- | --- |
| **#2** | ABS ( ( sleep* OR insomnia OR polysomnograph* OR actigraph* OR {restless leg syndrome} OR hypersomnia ) ) |
| **#3** | #1 AND #2 |
| **#4** | #3 AND ( LIMIT-TO ( DOCTYPE , "ar" ) OR LIMIT-TO ( DOCTYPE , "re" ) ) AND ( LIMIT-TO ( LANGUAGE , "english" ) ) |

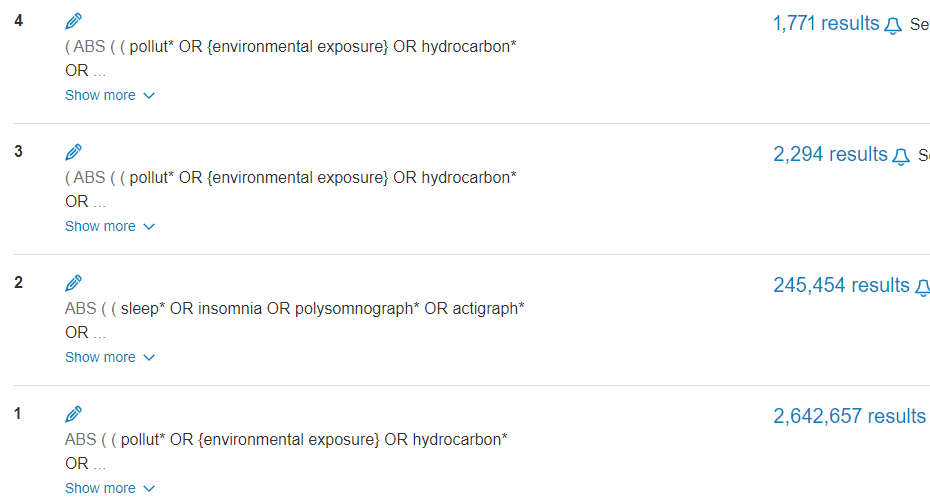

**Agricultural & Environmental Science Collection (ProQuest) = 1755**

**5/17/2021**

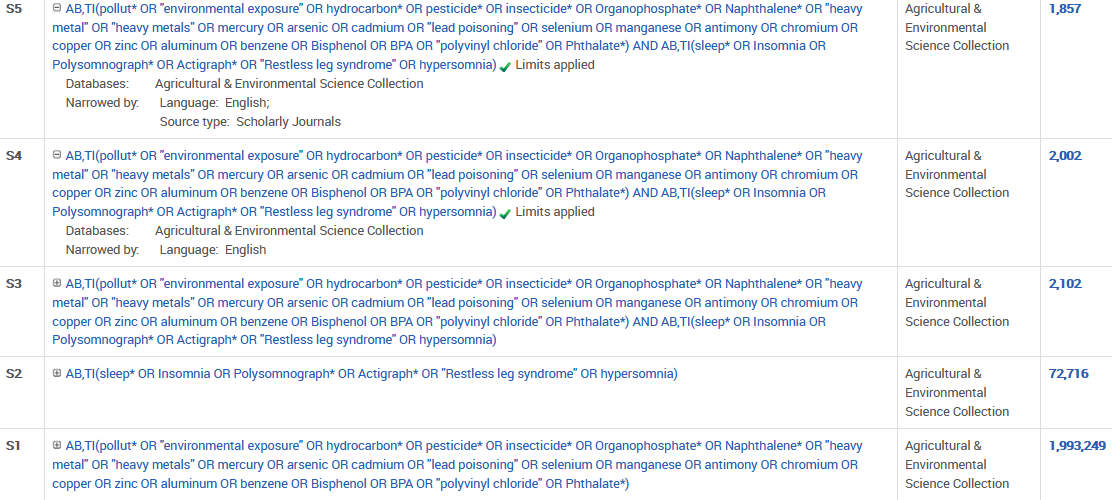
