## Supplemental File 2_Reviews for "Is exposure to chemical pollutants associated with sleep outcomes? A systematic review"

| ***Supplemental File 2.*** *Summary table of identified review studies with mention of pollutants and sleep outcomes.* | | | |
| --- | --- | --- | --- |
| **Study** | **Type of review** | **Relevant pollutant(s)** | **Sleep outcome(s) mentioned in relation to exposure** |
| [(Akinseye et al. 2015)](https://sciwheel.com/work/citation?ids=12918140&pre=&suf=&sa=0&dbf=0) | Narrative | Air pollution (PM10, black carbon) | SDB, sleep efficiency, sleep duration |
| [(An et al. 2019)](https://sciwheel.com/work/citation?ids=12918144&pre=&suf=&sa=0&dbf=0) | Systematic | Air pollution (PM 10, PM 2.5, O3, NOx, air quality) | Duration |
| [(Cao et al. 2021)](https://sciwheel.com/work/citation?ids=12918153&pre=&suf=&sa=0&dbf=0) | Narrative | Air pollution (PM 1/2.5/10, NO2, SO2, O3, CO, black carbon, NOx, AQI) | Quality, duration, SDB |
| [(Clark et al. 2020)](https://sciwheel.com/work/citation?ids=12918164&pre=&suf=&sa=0&dbf=0) | Systematic | Air pollution (PM1, PM2.5, PM10, black carbon, NOx, SOx, O3) | OSA/SDB |
| [(Hunter & Hayden 2018)](https://sciwheel.com/work/citation?ids=5632907&pre=&suf=&sa=0&dbf=0) | Narrative | Air pollution (ozone, PM2.5, PM10, black carbon) | Sleep disturbances |
| [(Johnson et al. 2018)](https://sciwheel.com/work/citation?ids=11339211&pre=&suf=&sa=0&dbf=0) | Narrative | Air pollution, environmental pollution | OSA/SDB |
| [(Mayne et al. 2021)](https://sciwheel.com/work/citation?ids=12130957&pre=&suf=&sa=0&dbf=0) | Systematic | Air pollution | Duration, quality, variability, disturbances, OSA/SDB |
| [(Tenero et al. 2017)](https://sciwheel.com/work/citation?ids=10671548&pre=&suf=&sa=0&dbf=0) | Systematic | Air pollution (indoor and outdoor) | OSA/SDB |
| [(Mantua et al. 2019)](https://sciwheel.com/work/citation?ids=12918223&pre=&suf=&sa=0&dbf=0) | Narrative | Air pollution related to military | Sleep disorders |
| [(Urbanik et al. 2020)](https://sciwheel.com/work/citation?ids=13868148&pre=&suf=&sa=0&dbf=0) | Narrative | Air pollution (SHS, PM) | OSA |
| [(Edwards & Tchounwou 2005)](https://sciwheel.com/work/citation?ids=7285100&pre=&suf=&sa=0&dbf=0) | Narrative | Pesticides (methyl parathion, organophosphates) | Insomnia |
| [(Stallones & Beseler 2016)](https://sciwheel.com/work/citation?ids=12918290&pre=&suf=&sa=0&dbf=0) | Narrative | Pesticides, pesticide poisoning | Sleep disorders, sleep disturbances |
| [(Ji et al. 2017)](https://sciwheel.com/work/citation?ids=8259441&pre=&suf=&sa=0&dbf=0) | Systematic | Micronutrients (selenium, copper, manganese) | Duration, onset latency, awakenings, architecture, timing |
| [(Parmalee & Aschner 2017)](https://sciwheel.com/work/citation?ids=7843342&pre=&suf=&sa=0&dbf=0) | Narrative | Metals (manganese, copper, zinc, lead, mercury, aluminum) | Duration, sleep problems |
| [(Liu et al. 2021)](https://sciwheel.com/work/citation?ids=11356039&pre=&suf=&sa=0&dbf=0) | Narrative | Multiple (heavy metals, secondhand smoke, air pollutants) | Quality, duration, OSA/SDB, others |
| [(Schwartz et al. 2017)](https://sciwheel.com/work/citation?ids=9681374&pre=&suf=&sa=0&dbf=0) | Systematic with meta-analysis | Solvents, particulate dust exposure (other OSA-related occupational exposures) | OSA/SDB |
| [(Viaene et al. 2009)](https://sciwheel.com/work/citation?ids=12918306&pre=&suf=&sa=0&dbf=0) | Narrative | Solvents (occupational exposure) | Sleep disturbances, SDB/OSA |
| [(Tchounwou et al. 2003)](https://sciwheel.com/work/citation?ids=12918293&pre=&suf=&sa=0&dbf=0) | Narrative | Dinitrotoluene | Insomnia |
| [(Kerr 2015)](https://sciwheel.com/work/citation?ids=2407920&pre=&suf=&sa=0&dbf=0) | Narrative | Gulf War exposures (pesticides, nerve agents) | Sleep problems, abnormalities |
| [(Worek et al. 2007)](https://sciwheel.com/work/citation?ids=12918273&pre=&suf=&sa=0&dbf=0) | Narrative | War-related exposures (nerve agents, OPs, sarin) | Insomnia |

[Akinseye OA, Williams SK, Seixas A, Pandi-Perumal SR, Vallon J, et al. 2015. Sleep as a mediator in the pathway linking environmental factors to hypertension: a review of the literature. *Int. J. Hypertens.* 2015:926414](https://sciwheel.com/work/bibliography/12918140)

[An R, Shen J, Ying B, Tainio M, Andersen ZJ, de Nazelle A. 2019. Impact of ambient air pollution on physical activity and sedentary behavior in China: A systematic review. *Environ. Res.* 176:108545](https://sciwheel.com/work/bibliography/12918144)

[Cao B, Chen Y, McIntyre RS. 2021. Comprehensive review of the current literature on impact of ambient air pollution and sleep quality. *Sleep Med.* 79:211–19](https://sciwheel.com/work/bibliography/12918153)

[Clark DPQ, Son DB, Bowatte G, Senaratna CV, Lodge C, et al. 2020. The association between traffic-related air pollution and obstructive sleep apnea: A systematic review. *Sleep Med. Rev.* 54:101360](https://sciwheel.com/work/bibliography/12918164)

[Edwards FL, Tchounwou PB. 2005. Environmental toxicology and health effects associated with methyl parathion exposure--a scientific review. *Int. J. Environ. Res. Public Health*. 2(3–4):430–41](https://sciwheel.com/work/bibliography/7285100)

[Hunter JC, Hayden KM. 2018. The association of sleep with neighborhood physical and social environment. *Public Health*. 162:126–34](https://sciwheel.com/work/bibliography/5632907)

[Ji X, Grandner MA, Liu J. 2017. The relationship between micronutrient status and sleep patterns: a systematic review. *Public Health Nutr.* 20(4):687–701](https://sciwheel.com/work/bibliography/8259441)

[Johnson DA, Billings ME, Hale L. 2018. Environmental determinants of insufficient sleep and sleep disorders: implications for population health. *Curr. Epidemiol. Rep.* 5(2):61–69](https://sciwheel.com/work/bibliography/11339211)

[Kerr KJ. 2015. Gulf War illness: an overview of events, most prevalent health outcomes, exposures, and clues as to pathogenesis. *Rev. Environ. Health*. 30(4):273–86](https://sciwheel.com/work/bibliography/2407920)

[Liu J, Ghastine L, Um P, Rovit E, Wu T. 2021. Environmental exposures and sleep outcomes: A review of evidence, potential mechanisms, and implications. *Environ. Res.* 196:110406](https://sciwheel.com/work/bibliography/11356039)

[Mantua J, Bessey A, Sowden WJ, Chabuz R, Brager AJ, et al. 2019. A review of environmental barriers to obtaining adequate sleep in the military operational context. *Mil. Med.* 184(7–8):e259–66](https://sciwheel.com/work/bibliography/12918223)

[Mayne SL, Mitchell JA, Virudachalam S, Fiks AG, Williamson AA. 2021. Neighborhood environments and sleep among children and adolescents: A systematic review. *Sleep Med. Rev.* 57:101465](https://sciwheel.com/work/bibliography/12130957)

[Parmalee NL, Aschner M. 2017. Metals and circadian rhythms. *Adv. Neurotoxicol.* 1:119–30](https://sciwheel.com/work/bibliography/7843342)

[Schwartz DA, Vinnikov D, Blanc PD. 2017. Occupation and Obstructive Sleep Apnea: A Meta-Analysis. *J. Occup. Environ. Med.* 59(6):502–8](https://sciwheel.com/work/bibliography/9681374)

[Stallones L, Beseler CL. 2016. Assessing the connection between organophosphate pesticide poisoning and mental health: A comparison of neuropsychological symptoms from clinical observations, animal models and epidemiological studies. *Cortex*. 74:405–16](https://sciwheel.com/work/bibliography/12918290)

[Tchounwou PB, Newsome C, Glass K, Centeno JA, Leszczynski J, et al. 2003. Environmental toxicology and health effects associated with dinitrotoluene exposure. *Rev. Environ. Health*. 18(3):203–29](https://sciwheel.com/work/bibliography/12918293)

[Tenero L, Piacentini G, Nosetti L, Gasperi E, Piazza M, Zaffanello M. 2017. Indoor/outdoor not-voluptuary-habit pollution and sleep-disordered breathing in children: a systematic review. *Transl. Pediatr.* 6(2):104–10](https://sciwheel.com/work/bibliography/10671548)

[Urbanik D, Martynowicz H, Mazur G, Poręba R, Gać P. 2020. Environmental Factors as Modulators of the Relationship between Obstructive Sleep Apnea and Lesions in the Circulatory System. *J. Clin. Med.* 9(3):](https://sciwheel.com/work/bibliography/13868148)

[Viaene M, Vermeir G, Godderis L. 2009. Sleep disturbances and occupational exposure to solvents. *Sleep Med. Rev.* 13(3):235–43](https://sciwheel.com/work/bibliography/12918306)

[Worek F, Eyer P, Szinicz L, Thiermann H. 2007. Simulation of cholinesterase status at different scenarios of nerve agent exposure. *Toxicology*. 233(1–3):155–65](https://sciwheel.com/work/bibliography/12918273)
