## Supplemental File 3_Data extraction questions for "Is exposure to chemical pollutants associated with sleep outcomes? A systematic review"

**Supplemental File 3**. Examples of the data extraction templates used for epidemiologic studies, case studies, and reviews. Cohort study data was extracted using Covidence, which case study and review data was extracted using Google forms.

Epidemiologic Studies:

1. Last name of first author
2. Year published
3. Date(s) of study
4. Geographical location
5. Study design
6. Source / location of exposure
7. Sample number exposed / unexposed
8. Sample number cases / controls
9. Sample number exposed / unexposed by salient characteristics
10. What is the exposure / pollutant?
11. How was the exposure reported?
12. What was used to measure the exposure?
13. What is the sleep-related outcome?
14. How was the sleep-related outcome reported?
15. What was used to measure the sleep-related outcome?
16. Did they adjust for covariates in the exposure-outcome relationship? If so, which covariates?
17. Strengths
18. Limitations
19. General summary of findings and direction of association
20. Any other notes about the study, such as potential sources of bias
21. Conflict of interest?

Case studies:

1. Last name of first author
2. Year published
3. Date(s) of study
4. Geographical location
5. Exposure setting
6. Case evaluation setting
7. How many patients are included in the case study?
8. Sample number by salient characteristics?
9. What is the exposure / pollutant?
10. How was the exposure measured?
11. What is the sleep-related outcome?
12. How was the sleep outcome measured?
13. General notes about the study (strengths, limitations, general summary)

Reviews:

1. Last name of first author
2. Year published
3. What date(s) range for published material did the review include?
4. What is the last search date of the review?
5. What exposure(s) does the review cover?
6. What sleep-related outcome(s) does the review cover?
7. Please provide DOIs or citation information for each of the potentially relevant studies included in the review.
8. Notes on review, methods, and review quality.
