## Supplemental File 4_Risk of bias questions for "Is exposure to chemical pollutants associated with sleep outcomes? A systematic review"

**Supplemental File 4**. Examples of the quality assessment templates for risk of bias (RoB) with example criteria used for epidemiologic and case studies.

Epidemiologic studies:

For each of the RoB quality assessment questions, one of the following categories was chosen:

a. Definitely low risk of bias
b. Probably low risk of bias
c. Probably high risk of bias
d. Definitely high risk of bias
e. No information on which to base judgement

The following questions pertain only to experimental/interventional studies:

1. (HCT only) Was administered dose or exposure level adequately randomized?
2. (HCT only) Was allocation to study groups adequately concealed?
3. (HCT only) Bias due to deviations from intended intervention?

The following questions pertain to all epidemiologic studies, except question 1 (only for observational studies):

1. Bias in selection of participants into the study: did selection result in appropriate comparison groups (exclude for experimental)?
2. Bias due to confounding: did the study design or analysis account for important confounding and modifying variables?
3. Bias in classification / measurement of exposure: can we be confident in the exposure classification?
4. Bias due to missing data: were outcome data complete without attrition or exclusion from analysis?
5. Bias in measurement of outcomes: can we be confident in the outcome assessment?
6. Bias in selection of reported result: were all measured outcomes reported?
7. Were there no other potential threats to internal validity (e.g., statistical methods were appropriate and researchers adhered to the study protocol)?

Case studies:

For quality assessment questions 1-8, one of the following RoB categories was chosen:

a. Yes
b. No
c. NA
1. Does the patient(s) represent(s) the whole experience of the investigator (center) or is the selection method unclear to the extent that other patients with similar presentation may not have been reported?
2. Was the exposure adequately ascertained?
3. Was the outcome adequately ascertained?
4. Were other alternative causes that may explain the observation ruled out?
5. Was there a challenge/rechallenge phenomenon?
6. Was there a dose–response effect?
7. Was follow-up long enough for outcomes to occur?
8. Is the case(s) described with sufficient details to allow other investigators to replicate the inferences related to their own practice?
9. Overall judgement about methodological quality (based on questions deemed most critical in specific scenario)
   a. Good
   b. Fair
   c. Poor
10. Notes on quality assessment
